# Surgery for Esotropia: The “Legend” of the Dose-Response Curve Re-visited and the Optimal Surgical Strategy

**DOI:** 10.1101/2025.11.07.25339775

**Authors:** Christopher T. Leffler, Emilia Varrone, Satya Siri Paruchuri, Catherine Phan

## Abstract

**Objective:** To determine clinical predictors of surgical failures following horizontal strabismus surgery for esotropia, in order to estimate the optimal surgical strategy.

**Design:** Retrospective pooled observational case series of published cases.

**Subjects:** Patients having horizontal strabismus surgery for esotropia, published between 1940 and 2025, with known preoperative deviation, surgical approach, and outcome.

**Methods:** Clinical data from individual patients having strabismus surgery for esotropia was recorded from published case series, and analyzed using multivariable logistic regression to predict over- and under-correction.

**Main Outcome Measure:** Surgical failure, as determined by reoperation, suture adjustment, or postoperative strabismus of 10 prism diopters or more.

**Results:** We abstracted individual patient data for 3518 surgeries from 163 publications. Binocular (as compared with monocular) surgery was associated with fewer under-corrections (odds ratio [OR] 0.75, 95% CI 0.61 to 0.92, p=0.005) and more over-corrections (OR 1.87, 95% CI 1.26 to 2.79, p=0.002, n=3266). Increasing preoperative deviation was associated with more under-corrections (OR 1.06/°, 95% CI 1.05/° to 1.07/°, p<0.0001) and fewer overcorrections (OR 0.97/°, 95% CI 0.95/° to 0.99/°, p=0.001, n=3266). Increasing surgical dose was associated with fewer under-corrections (OR 0.95/mm, 95% CI 0.91/mm to 0.99/mm, p=0.01), and more over-corrections (OR 1.08/mm, 95% CI 1.01/mm to 1.16/mm, p=0.02, n=3266). The failure rate was minimized with a large per-muscle surgical dose. As the preoperative deviation increases, one progresses from unilateral recessions, to unilateral recession-resections, and then bi-medial recessions. Under a range of assumptions, bi-medial recessions of 6 mm are optimal for preoperative deviations of 45 to 50 prism diopters.

**Conclusions:** Larger doses for esotropia surgery do produce a larger response. Most models predicted the lowest failure rates with large recessions or resections, with additional muscles operated for larger preoperative deviations. Thus, the analysis supports the approach of Scobee (1951) over that of Parks (1975). The preferred surgical strategy depends on multiple factors.

## Introduction

For decades, strabismus surgeons have debated the preferred surgical strategy and dose for esotropia. Some, such as Richard Scobee in 1951, have recommended reasonably large surgeries in all cases (e.g. 6 mm medial rectus recessions), with the only variation being the number of muscles operated (Scobee 1951). Others, such as Marshall Parks’ text of 1975, recommended two-muscle surgeries in all cases, but with graded surgical dosing, with smaller doses used for smaller angles of deviation (Parks 1975, pp. 108-121).

One conventional approach to predict the likely outcome of strabismus surgery has been to look at the change in alignment as a function of the surgical dose measured in mm. However, it has been pointed out that the major predictor of the change in alignment is the preoperative alignment itself (Archer 2018). Moreover, it is conventional for surgeons to deliver a larger surgical dose when the preoperative deviation is greater (Parks 1975, pp. 108-121). Therefore, both change in alignment and surgical dose will be correlated with preoperative alignment, and, hence, with each other, even if the surgical dose does not change the size of the response in a causative manner. In other words, spurious dose-response relations can be generated, even with small datasets (Archer 2018).

Recent approaches to understand strabismus surgery failures have looked at registries or insurance datasets with thousands of patients (Christensen 2018, Leffler 2015, Leffler 2016, Oke 2022, Leffler 2024, Oke 2024). Unfortunately, these large datasets have not included preoperative or postoperative deviations, or the surgical dose in mm (Christensen 2018, Leffler 2015, Leffler 2016, Oke 2022, Leffler 2024, Oke 2024). This fact has limited their ability to offer insight into the preferred surgical approach.

Therefore, we abstracted and analyzed individual patient data from case series published from 1940 to 2025 to determine predictors of over- and under-correction following surgery for esotropia, and to better understand the preferred surgical strategy and dose.

## Methods

Studies of patients having medial rectus recessions and/or lateral rectus resections for esotropia were identified in Google Scholar by searching for terms such as “esotropia”, “medial rectus recession”, “lateral rectus resection”, combined with “mm”, in October 2025, as tabulated (Supplemental Table 1). Study inclusion criteria included: individual patient data (preoperative deviation, mm of surgery performed, postoperative status as success, over- or under-correction), relevant information provided in the English language, and at least two patients with strabismus. Only surgeries on previously unoperated muscles were included. We did not use surgeries with transpositions, posterior fixation sutures, pulley fixation, slanting recessions, Y-splitting of the medial rectus, or union surgery. We did not include case series which were nonrepresentative because the authors selected them on the basis that they were known to have succeeded or failed. We did not study surgeries for exotropia. We did not include retracted studies.

**Supplemental Table 1.**
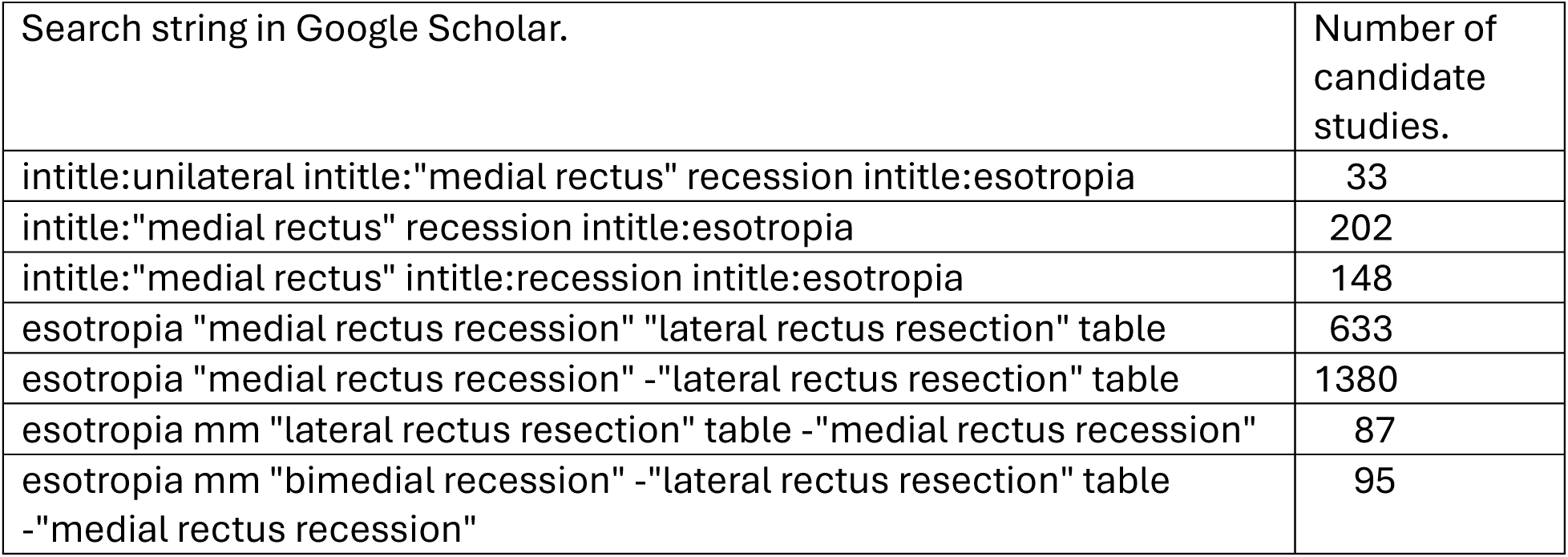
Search strategy to identify candidate studies, in October 2025.

We identified qualifying studies (Appendix 1) from over 700 candidate studies for review from 1940 to 2025. The studies rejected, primarily for lacking individual patient information with adequate specificity, such as the mm of surgery, are listed in Appendix 2. Steven Archer provided us with raw data on strabismus outcomes without any individually identifiable patient information (Archer 2009).

Under-corrections and over-corrections were defined as deviations of 10 prism diopters or more of esotropia or exotropia postoperatively, respectively, or a reoperation for horizontal strabismus. In addition, when the preoperative deviation was 10 prism diopters or less, the case was considered a failure if the postoperative deviation was not improved. The follow-up date in the study is the same as the outcome date used for the data analysis. For studies which reported recessions of the medial rectus muscle as distance from the limbus, the original insertion distance was assumed to be 5.5 mm.

Multivariable logistic regression models to predict under- and over-corrections were prepared with the following variables: 1. preoperative deviation in degrees (°). When both far and near deviations were provided, the larger deviation was used. When deviations with and without refractive correction were provided for accommodative esotropia, the deviation with refractive correction was used. 2. The total number of horizontal rectus muscles operated. 3. binocular vs. monocular surgery. Variables tested in the multivariable models included those significant in univariate analysis, as well as the total surgical dose (mm). The surgical dose was defined as the sum of recessions and resections in mm.

We explored the effect on the predicted optimal surgical strategy if over-corrections are more undesirable than under-corrections, perhaps because under-corrections are less likely than over-corrections to produce more severe adverse outcomes, such as a failure of ductions. We also determined for maximal surgery the preoperative deviation for which at least 20 and at least 40 cases had been performed. The study was approved by the VCU Office of Research Subjects Protection.

## Results

### Patient and surgery characteristics

We identified 163 qualifying reports containing 3518 surgeries in 3504 patients (Appendix 1).

Most analyses focused on 3266 cases without thyroid disease, and for which, if adjustable sutures were used, the initial alignment (before adjustment) was known (Table 1, Supplemental Tables 2-3). Only 62 of the surgeries in these baseline analyses used adjustable sutures.

**Table 1.** Multivariable Logistic Predictors of Under-corrections and Over-corrections after Strabismus Surgery for Esotropia.

| Variable | Under-corrections |  | Over-corrections |  |
| --- | --- | --- | --- | --- |
|  | Odds Ratio | p value | Odds Ratio | p value |
| Preop. Deviation (°) | 1.06 (1.05, 1.07) | <0.0001 | 0.97 (0.95, 0.99) | 0.001 |
| surgical dose (mm) | 0.95 (0.91, 0.99) | 0.01 | 1.08 (1.01, 1.16) | 0.02 |
| No. of muscles operated | 0.98 (0.75, 1.26) | 0.85 | 1.61 (1.07, 2.41) | 0.02 |
| Binocular surgery | 0.75 (0.61, 0.92) | 0.005 | 1.87 (1.26, 2.79) | 0.002 |

Probability of under-correction = $1 / (1 + \exp(-(-1.447552 + 0.055703 * (\text{preoperative deviation in degrees}) - 0.053285 * (\text{total surgery in mm}) - 0.291698 * (\text{binocular surgery}) - 0.024721 * (\text{number of muscles operated}))))$ .
Probability of over-correction = $1 / (1 + \exp(-(-4.195557 - 0.033682 * (\text{preoperative deviation in degrees}) + 0.079761 * (\text{total surgery in mm}) + 0.626323 * (\text{binocular surgery}) + 0.474277 * (\text{number of muscles operated}))))$ .
The variable binocular surgery equals 1 for binocular surgery, and 0 for monocular surgery. n=3266 cases, including 62 with adjustable sutures. No cases of thyroid eye disease were included.

**Supplemental Table 2.** Univariate Predictors of Under-corrections from Strabismus Surgery for Esotropia.

| Factor. | Factor Present | Factor Absent | P |
| --- | --- | --- | --- |
| Child (age<18 years) | 26.7% (673/2516) | 35.9% (132/368) | <0.001 |
| Child <6 years old | 22.5% (356/1579) | 30.2% (294/975) | <0.001 |
| Age ≥ 45 years | 21.7% (25/115) | 27.4% (803/2931) | 0.20 |
| Binocular Surgery | 26.7% (561/2098) | 28.5% (333/1168) | 0.29 |
| One muscle surgery | 21.3% (138/649) | 28.9% (756/2617) | <0.001 |
| 3- or 4-muscle surgery | 18.5% (50/271) | 28.2% (844/2995) | <0.001 |
| Four muscle surgery | 30.2% (16/53) | 27.3% (878/3213) | 0.64 |
| Total surgery ≥ 11 mm | 29.3% (491/1678) | 25.4% (403/1588) | 0.06 |
| Preop. Deviation ≥ 38 pd | 33.1% (537/1623) | 21.7% (357/1643) | <0.001 |
| Follow-up ≥ 6 months | 26.3% (542/2063) | 31.3% (324/1036) | 0.004 |
| Duane syndrome | 18.5% (23/124) | 27.8% (853/3064) | 0.02 |

Totals vary by row due to missing or ambiguous data. N=3266 cases. Includes 62 adjustable cases. Includes no thyroid eye disease cases.

**Supplemental Table 3.** Univariate Predictors of Over-corrections from Strabismus Surgery for Esotropia.

| Factor. | Factor Present | Factor Absent | p value |
| --- | --- | --- | --- |
| Child (age<18 years) | 7.4% (186 / 2516) | 4.6% (17 / 368) | 0.06 |
| Child < 6 years old | 8.0% (126/1579) | 6.5% (63/975) | 0.16 |
| Age ≥45 years | 8.7% (10 / 115) | 6.7% (196 / 2931) | 0.35 |
| Binocular Surgery | 8.5% (179/2098) | 3.3% (39/1168) | <0.001 |
| One muscle surgery | 2.5% (16/649) | 7.7% (202/2617) | <0.001 |
| 3- or 4-muscle surgery | 15.1% (41/271) | 5.9% (177/2995) | <0.001 |
| Four muscle surgery | 24.5% (13/53) | 6.4% (205/3213) | <0.001 |
| Total surgery ≥11 mm | 8.0% (134/1678) | 5.3% (84/1588) | 0.002 |
| Preop. Deviation ≥ 38 pd | 7.7% (125/1623) | 5.7% (93/1643) | 0.02 |
| Follow-up ≥ 6 months or more | 7.0% (145/2063) | 5.5% (57/3099) | 0.11 |
| Duane syndrome | 2.4% (3/124) | 6.9% (210/3064) | 0.06 |

Totals vary by row due to missing or ambiguous data. N=3266 cases. Includes 62 adjustable cases. Includes no thyroid eye disease cases.

The mean preoperative deviation was 22.3° (SD 9.7°) (which corresponds with 40.9 PD), with a range of 0° to 52° (0 to 128 PD, n=3266). (Some strabismus surgeries are performed on patients with orthophoria in primary gaze but esotropia in lateral gaze due to incomitance, or for accommodative esotropia seen only without correction (Phanphruk 2023; Gunduz 2019; Hornbrook 1984)).

The surgical approach was bi-medial rectus (n=1616 surgeries), unilateral medial rectus (n=572), both horizontal muscles in one eye (n=517), bilateral lateral rectus (n=224), bi-medial rectus with one lateral rectus (n=205), unilateral lateral rectus (n=79), all four horizontal muscles (n=50), or bilateral lateral rectus muscle with one medial (n=3). Of 2963 surgeries which involved at least one medial rectus recession, 2361 (80%) were between 3 mm and 6 mm, inclusive.

### Associations with Over- and Under-correction

Binocular (as compared with monocular) surgery was associated with fewer under-corrections (odds ratio [OR] 0.75, 95% CI 0.61 to 0.92, p=0.005) and more over-corrections (OR 1.87, 95% CI 1.26 to 2.79, p=0.002, n=3266, Table 1). Increasing preoperative deviation was associated with more under-corrections (OR 1.06/°, 95% CI 1.05/° to 1.07/°, p<0.0001) and fewer over-corrections (OR 0.97/°, 95% CI 0.95/° to 0.99/°, p=0.001). Increasing surgical dose was associated with fewer under-corrections (OR 0.95/mm, 95% CI 0.91/mm to 0.99/mm, p=0.01), and more over-corrections (OR 1.08/mm, 95% CI 1.01/mm to 1.16/mm, p=0.02). For a given surgical dose in mm, operating on more muscles increased the likelihood of overcorrections (odds ratio 1.61/muscle, 95% CI 1.07/muscle to 2.41/muscle, p=0.02, Table 1). This model included no cases of thyroid eye disease (n=3266, Table 1).

Most surgeries (87.2%, 2516 of 2884 surgeries with data available) were performed in children under age 18 years. In univariate analysis, children were less likely to be under-corrected (p<0.001, Supplemental Table 1), and tended to have more overcorrections, though not significantly so (p=0.06, Supplemental Table 2). In multivariable logistic regression, children had a lower probability of under-correction (odds ratio 0.59, 95% CI 0.46 to 0.76, p<0.001, n=2884).

Postoperative follow-up was 6 months or more in 2063 of 3099 surgeries with data available (66.6%). In univariate analysis, follow-up of 6 months or more was associated with fewer under-corrections (26.3% vs. 31.3%, p=0.004, Supplemental Table 1), but only a nonsignificant trend towards more overcorrections (7.0% vs. 5.5%, p=0.11, Supplemental Table 2). With multivariable logistic regression (n=3099), follow-up over 6 months was associated with fewer under-corrections (odds ratio 0.78, 95% CI 0.66 to 0.92, p=0.004). These observations are consistent with an exotropic drift after surgery for esotropia.

Duane syndrome was present in 124 surgeries (3.8%). In univariate analysis, Duane syndrome was associated with a lower risk of under-correction (18.5% vs. 27.8%, p=0.02, Supplemental Table 1). However, in a multivariable logistic regression model, the association of Duane syndrome with under-corrections was no longer significant (odds ratio 0.69, 95% CI 0.43 to 1.11, p=0.13, n=3188).

### Preferred Surgical Approach

From the equations for the probability of under- and over-corrections, the total failure rate can be determined. The optimal surgical strategy is that which minimizes the failure rate. For instance, for bi-medial recessions, if under- and over-corrections are considered equally undesirable, the lowest failure rate for 20 prism diopters of esotropia occurs with 5 mm recessions for both eyes (Figure 1). For monocular surgeries, maximal surgical doses are recommended for this deviation (Supplemental Figs. 1-2).

**Figure 1.**
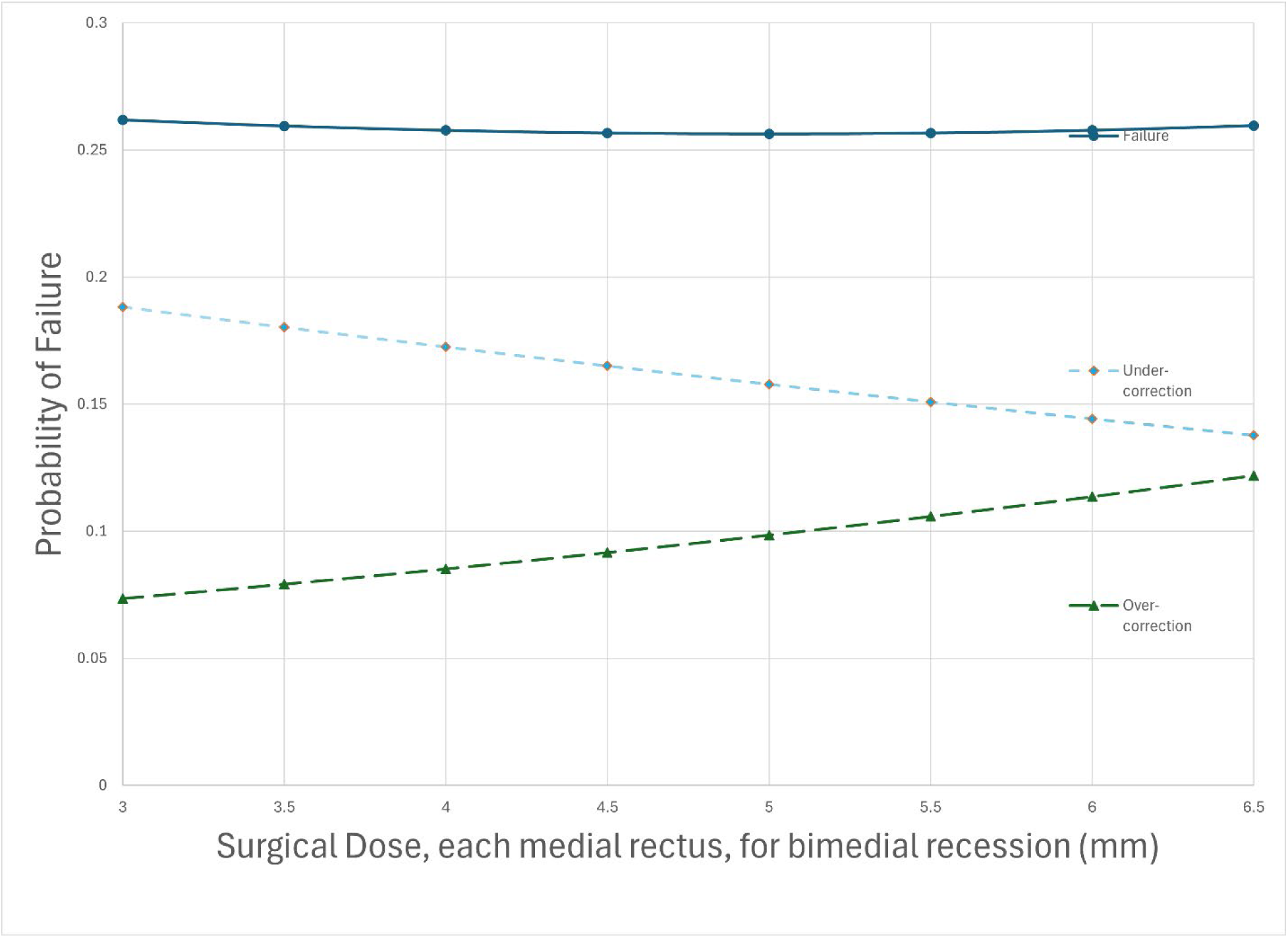
Probability of failures, under-corrections, and over-corrections for bi-medial recession with a preoperative deviation of 20 prism diopters, as a function of surgical dose in mm per muscle. This is the basic model in which under- and over-corrections are considered equally undesirable.

**Supplemental Figure 1.**
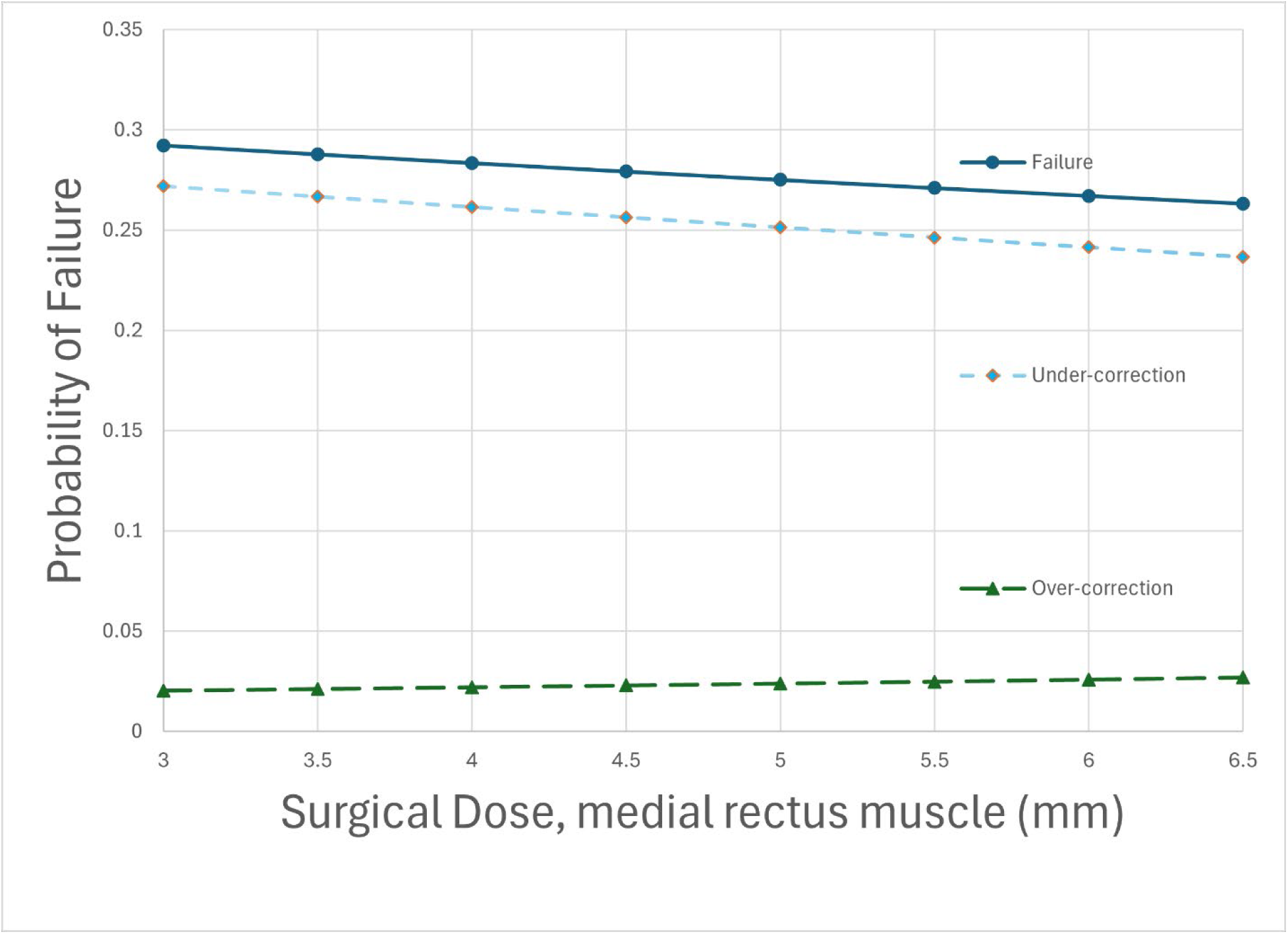
Probability of failure, under-correction, and over-correction for unilateral medial rectus recession for 20 PD preoperative deviations as a function of surgical dose in mm, for the basic model, in which under- and over-corrections are considered equally undesirable.

**Supplemental Figure 2.**
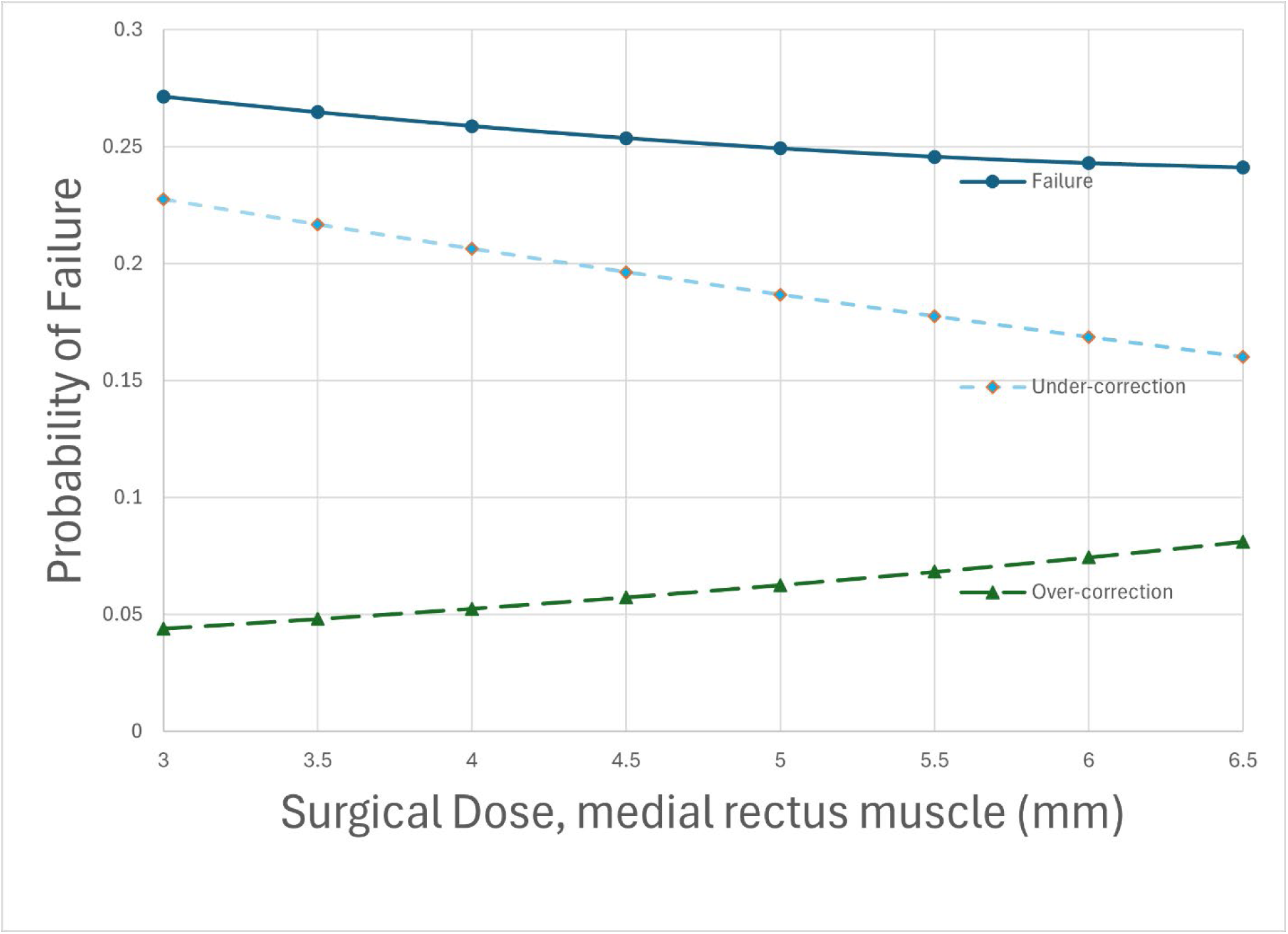
**Probability of failure, under-correction, and over-correction for unilateral 2-muscle strabismus surgery for 20 PD preoperative deviations as a function of surgical dose for medial rectus recession plus lateral rectus resection, for the model in which under- and over-corrections were considered equally undesirable.** The resections were one third larger than the recessions.

Although the precise optimal surgical approach depends on the specific assumptions, certain common themes can be identified. If one must perform two-muscle surgery, as in the tables of Parks (1975), then larger preoperative deviations do require larger surgical doses, measured in mm (Supplemental Tables 4,5). The relative value which one places on over-corrections and under-corrections influences the surgical dose determined to be optimal in the models. For instance, assuming that under-corrections only result in significant adverse findings 64.5% as often as over-corrections results in tables which approximate the bi-medial recession tables of Parks (1975), with optimal doses of 4 mm OU for 25 pd and 4.5 mm OU for 30 pd (Supplemental Table 4).

**Supplemental Table 4.** Bi-medial recession tables.

| Preoperative Deviation (PD) | Medial rectus recession, both eyes, mm (each muscle). |  |  |
| --- | --- | --- | --- |
|  | Minimize Failure Rate | Under-corrections weighted 64.5% | Under-corrections weighted 40% |
| 0 to 10 | 3.0 | 3.0 | 3.0 |
| 15 | 4.0 | 3.0 | 3.0 |
| 20 | 5.0 | 3.0 | 3.0 |
| 25 | 6.0 | 4.0 | 3.0 |
| 30 | 6.0 | 4.5 | 3.0 |
| 35 | 6.0 | 5.5 | 3.0 |
| 40 | 6.0 | 6.0 | 4.0 |
| 45 | 6.0 | 6.0 | 5.0 |
| 50 | 6.0 | 6.0 | 6.0 |

The “minimize failure rate” model weights under-corrections and over-corrections as equally undesirable. The subsequent models considered under-corrections to be 64.5% and 40% as likely as over-corrections to result in severe adverse outcomes. With a weight of 64.5%, the tables approximate the bi-medial recession tables of Parks (1975). Values from 3.0 to 6.0 mm were tested in the model.

Moreover, assuming that under-corrections only result in significant adverse findings 40% as often as over-corrections approximates the unilateral recess / resect tables of Parks (1975), with an optimal dose of 4 mm recession / 6 mm resection for 25 pd (Supplemental Table 5).

**Supplemental Table 5.** Table for unilateral medial rectus recession plus lateral rectus resection.

| Preoperative Deviation (PD) | Medial rectus recession (mm) / Lateral rectus resection (mm) |  |  |
| --- | --- | --- | --- |
|  | Minimize Failure Rate | Under-corrections weighted 64.5% | Under-corrections weighted 40% |
| 0 | 4.0/5.3 | 3.0/4.0 | 3.0/4.0 |
| 5 | 5.0/6.7 | 3.0/4.0 | 3.0/4.0 |
| 10 | 5.5/7.3 | 4.0/5.3 | 3.0/4.0 |
| 15 | 6.0/8.0 | 4.5/6.0 | 3.0/4.0 |
| 20 | 6.0/8.0 | 5.5/7.3 | 3.5/4.7 |
| 25 | 6.0/8.0 | 6.0/8.0 | 4.0/5.3 |
| 30 | 6.0/8.0 | 6.0/8.0 | 5.0/6.7 |
| 35 | 6.0/8.0 | 6.0/8.0 | 6.0/8.0 |

The “minimize failure rate” model weights under-corrections and over-corrections as equally undesirable. The subsequent models considered under-corrections to be 64.5% and 40% as likely as over-corrections to result in severe adverse outcomes. With a weight of 40%, the table approximates the unilateral recess/resect table of Parks (1975). Medial rectus recessions from 3.0 to 6.0 mm were tested in the model, with the lateral rectus resections one third larger than the recessions.

If the restriction that surgery must involve two muscles is removed, then, all the models recommend maximal (or near-maximal) surgery. One does not perform small recessions or resections. Rather, if one decides to operate on a muscle, one performs a reasonably large surgery, in accord with the approach of Scobee (1951). If the deviation is small one operates on few muscles, and if large, one operates on more muscles. Moreover, there is a consistent progression as one moves from smaller to larger preoperative deviations: 1) unilateral recession; 2) unilateral recess-resect; 3) bi-medial recession; 4) 3- muscle surgery; 5) 4-muscle surgery (Table 2, Figure 2).

**Table 2.** Optimal Surgical Approach for Esotropia.

| Number of muscles | Surgical Approach | Preoperative Deviation (prism diopters): |  |  |  |  |  |
| --- | --- | --- | --- | --- | --- | --- | --- |
| | | Basic model. | Under-corrections weighted 64% | Under-corrections weighted 64% + 2% threshold | Under-corrections weighted 40% | With $\geq 20$ cases | With $\geq 40$ cases |
| 1 | MR recess 4 mm | | | | | $\leq 7$ | $\leq 12$ |
| | MR recess 5 mm | | | | 0 | $\leq 8$ | $\leq 12$ |
| | MR recess 6 mm. | 0-5 | 0-20 | 0-30 | 5-30 | $\leq 14$ | $\leq 16$ |
| 2 | MR recess 5 mm + LR resect 6.7 mm | | | | | $\leq 35$ | $\leq 36$ |
| | MR recess 5.5 mm + LR resect 7.3 mm | 10 | | | | $\leq 55$ | $\leq 60$ |
| | MR recess 6 mm + LR resect 8 mm | 15-30 | 25-40 | 35-40 | 35-40 | $\leq 55$ | $\leq 62$ |
| | MR recess 6 mm OU | 35-50 | 45-70 | 45-120 | 45-120 | $\leq 25$ | $\leq 35$ |
| 3 | MR recess 6 mm OU + LR resect 8 mm | 55-80 | 75-120 |  |  |  |  |
| 4 | MR recess 6 mm OU + LR resect 8 mm OU | 85-120 |  |  |  |  |  |

The basic model minimizes the raw failure rate, and considers under- and over-corrections equally undesirable. The weighted model assumes under-corrections are only 64.5% or 40% as likely to produce severe adverse outcomes as overcorrections. These assumptions produce optimal bi-medial recession and unilateral recess/resect tables, respectively, which approximate the Parks tables. The weighted plus threshold model also requires that operation on each additional muscle must decrease the failure rate by 2% to justify the added surgical risk. The final 2 columns indicate the preoperative deviation for which there are at least 20 (or 40) cases in the database using that surgical strategy (or one more aggressive). For instance, there are at least 40 cases of unilateral medial rectus recession of 6 mm for preoperative deviation of $\leq 16$ prism diopters.

**Figure 2.**
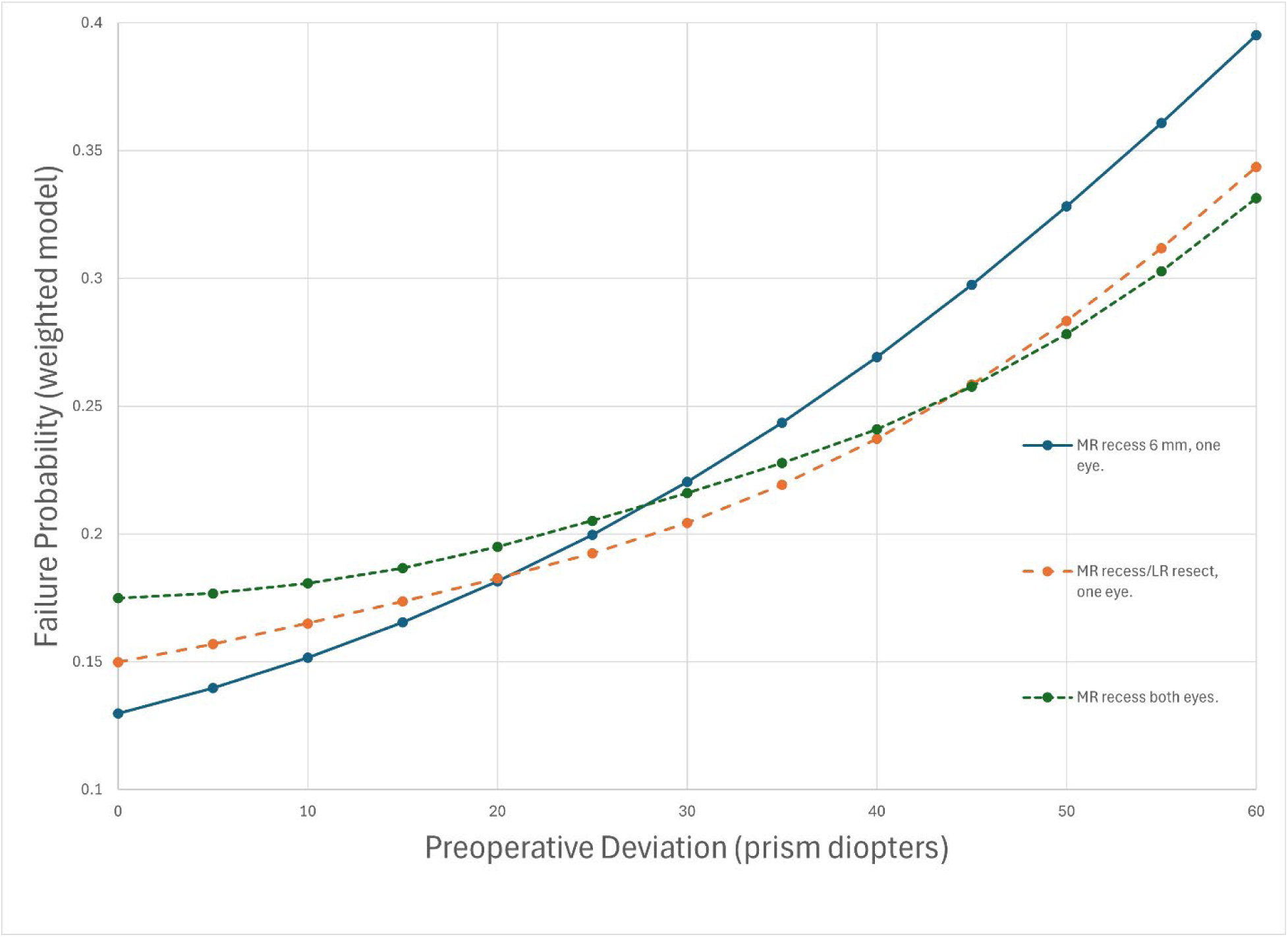
Failure probability as a function of preoperative deviation, for each surgical strategy (unilateral medial rectus recession, unilateral medial rectus recession with lateral rectus resection, and bi-medial recession). This is the model with weighting, so that under-corrections are considered only 64.5% as likely as over-corrections to result in significant adverse effects, in order to reproduce the bi-medial recession strabismus table of Parks (1975). The mm of surgery for each strategy was selected to minimize the weighted failure rate: medial rectus recession one eye of 6 mm; medial rectus recession/lateral rectus and bi-medial recession optimal dosages as in Supplemental Tables 4 and 5.

Finally, certain preoperative deviations have the same approach regardless of the weighting assumptions used. For instance, preoperative deviations of 45 to 50 prism diopters are optimally treated with 6-mm bi-medial recessions in all models tested.

According to the regression equations, small preoperative deviations are optimally treated with a large unilateral medial rectus recession. One caution is that there are limited data with this approach for the smallest deviations. For instance, there were at least 40 cases for which a medial rectus recession of at least 6 mm was performed for a preoperative deviation of 16 prism diopters or less. There were at least 40 cases for which a unilateral recession of 5 mm was performed for a preoperative deviation of 12 prism diopters or less (Table 2). If one performs a large recession for smaller deviations, one is venturing into territory with limited data.

### Thyroid Eye Disease

In a multivariable model which included 103 cases of thyroid eye disease, the rates of under-correction (p=0.44) and overcorrection (p=0.35) were not significantly associated with the surgical dose (Supplemental Table 6). This lack of association was likely due to the small number of thyroid cases.

**Supplemental Table 6.** Multivariable Logistic Predictors of Under-corrections and Overcorrections after Strabismus Surgery for Esotropia (includes cases of thyroid disease).

| Variable | Under-corrections |  | Over-corrections |  |
| --- | --- | --- | --- | --- |
|  | Odds Ratio | p value | Odds Ratio | p value |
| Preop. Deviation (°) | 1.06 (1.05, 1.07) | <0.0001 | 0.97 (0.95, 0.99) | 0.001 |
| surgical dose (mm) |  |  |  |  |
| ...if thyroid disease | 0.97 (0.91, 1.04) | 0.44 | 1.06 (0.94, 1.21) | 0.35 |
| ...if non-thyroid | 0.95 (0.91, 0.99) | 0.01 | 1.08 (1.01, 1.16) | 0.02 |
| Thyroid eye disease | 0.69 (0.24, 2.01) | 0.50 | 0.37 (0.03, 4.98) | 0.45 |
| No. of muscles operated | 0.94 (0.73, 1.20) | 0.61 | 1.59 (1.007, 2.36) | 0.02 |
| Binocular surgery | 0.76 (0.62, 0.93) | 0.009 | 1.82 (1.23, 2.70) | 0.003 |

The variable “thyroid eye disease” equals 1 if thyroid eye disease present, and 0 otherwise. The variable “binocular surgery” equals 1 for binocular surgery, and 0 for monocular surgery. n=3369 cases, including 67 with adjustable sutures, and 103 with thyroid eye disease.
Probability of Under-correction = $1 / (1 + \exp(-(-1.428055 + 0.056611 * (\text{preoperative deviation in degrees}) - 0.270096 * (\text{binocular surgery}) - 0.064695 * (\text{number of muscles operated}) - 0.367631 * (\text{thyroid disease}) - 0.026136 * (\text{mm of surgery if thyroid disease}) - 0.051293 * (\text{mm of surgery if not thyroid disease})))$
Probability of over-correction = $1 / (1 + \exp(-(-4.183356 - 0.031800 * (\text{preoperative deviation in degrees}) + 0.597680 * (\text{binocular surgery}) + 0.463199 * (\text{number of muscles operated}) - 1.002918 * (\text{thyroid disease}) + 0.061185 * (\text{mm surgery if thyroid disease}) + 0.079007 * (\text{mm surgery if not thyroid disease}))))$

### Adjustable Sutures

Adjustable sutures were used for 216 cases. For this analysis, the outcome was determined post-adjustment. Only 6 over-corrections (2.8%) and 28 under-corrections (13.0%) were observed. Patients who were under-corrected did not differ according to whether surgery was binocular (p=0.28), by total mm of surgery (p=0.38), the presence of thyroid eye disease (p=0.51), or number of muscles operated (p=0.81). Likewise, the preoperative deviation in under-corrected patients 18.8 degrees (SD 7.7 degrees) was not significantly different than that of other patients (15.8 degrees, SD 8.8 degrees, p=0.07).

## Discussion

This study identified predictors of under- and over-correction following horizontal muscle surgery for esotropia, and provided insight into the optimal surgical strategy.

As one might expect, binocular surgery (as compared with monocular surgery) is less likely to result in under-corrections, and more likely to result in over-corrections.

An age-old question in strabismus surgery has been the degree to which a change in the surgical dose (in mm) changes the final alignment. As early as 1951, Scobee noted that “The amount of surgery obtained in a patient is usually directly proportional to the deviation present before surgery and is not particularly related to the amount of surgery performed as measured in millimeters.” (Scobee 1951) This point as been emphasized more recently (Archer 2018).

Nonetheless, our study confirmed the common-sense notion that larger doses of esotropia surgery do produce a larger response. The difficulty of demonstrating the dose-response curve for strabismus surgery has led to this curve being called a “legend” (Archer 2018). Our study sheds light on why the relationship is difficult to demonstrate. In short, strabismus surgery typically “works”. Failures are rare. For instance, in our study, only 7.0% of surgeries resulted in over-corrections. Therefore, a study of 100 patients with strabismus might only be expected to produce 7 over-corrections. In order to learn when failures occur, one needs to analyze thousands of cases.

The regression equations permit determination of optimal surgical strategies. For a variety of model assumptions, large medial rectus recessions (about 6 mm in this dataset) were recommended. The idea that a muscle should be operated aggressively, or not at all, is not new. Prangen wrote in 1947 that he performed maximal resections, and that “The medial rectus muscle I recess 5 mm, no more and no less.” In 1951, Stine advocated recessing the muscle to the equator, and wrote “if one has decided to attack a muscle, one should either recess or resect it fully”. Also in 1951, Scobee published a series of patients having 6 mm medial rectus recessions, regardless of whether surgery was unilateral or bilateral. According to our results, this “all-or-nothing” approach to each muscle is favored over the tables outlined in Parks in 1975, with graded doses and two-muscle surgery for all cases.

The models predicted that as the preoperative deviation increases, one progresses from a unilateral medial rectus recession, to a unilateral two-muscle surgery, and finally to bi-medial recessions. Larger deviations typically require not a larger per-muscle dose, but rather that one operate on more muscles. The precise cutoffs where one transitions to the next surgical approach depend on how one values under-corrections relative to over-corrections. Nonetheless, some common features were seen. For instance, preoperative deviations of 45 to 50 prism diopters were optimally treated with bi-medial recessions of 6 mm, over a range of model assumptions.

Over-corrections were uncommon in this dataset, and the fact that they were not strongly dependent on surgical dose could indicate that overcorrections occur by a different mechanism, such as by slipped muscles (Chen 2005), by stretched scars (Ludwig 2000), or by variation in the brain’s response to the operation.

The results of this large dataset do not definitively settle the question of optimal surgical approach, because patients and doctors might vary in their preferences. For instance, some surgeons might find over-corrections more objectionable than under-corrections. Moreover, some surgeons might require a certain absolute benefit before being willing to operate on an additional muscle. In addition, when one eye has much better vision than the other, a unilateral surgery might be preferred.

This study has multiple limitations. Most of the patients were children. Larger datasets, particularly with more adult data, could be valuable. Also, larger studies with more thyroid eye disease, and/or adjustable sutures, could help to better characterize the dose-response relations and optimal surgical strategy for these types of patients or surgical approaches. In addition, we defined over- and under-corrections based on alignment in primary gaze at near and far, but did not assess incomitance in lateral gaze.

## Data Availability

All data produced in the present study are available upon reasonable request to the authors

## Appendix 1. Included Studies

Adamopoulou C, Rao RC. Surgical correction of consecutive esotropia with unilateral medial rectus recession. Journal of Pediatric Ophthalmology & Strabismus. 2015 Nov 1;52(6):343-7.

Agrawal S, Singh V, Gupta SK, Agrawal S. Evaluating a new surgical dosage calculation method for esotropia. Oman Journal of Ophthalmology. 2013 Sep;6(3):165.

Alattar S, Saad MS, Rashed GE, Anwar M. Botulinum toxin augmented surgery versus conventional surgery in the management of large-angle concomitant esotropia: A randomized clinical trial. Oman Journal of Ophthalmology. 2024 Jan 1;17(1):84-90.

Ali MH, Berry S, Qureshi A, Rattanalert N, Demer JL. Decompensated esophoria as a benign cause of acquired esotropia. American journal of ophthalmology. 2018 Oct 1;194:95-100.

Altintas AG, Arifoglu HB, Arikan M, Simsek S. Clinical findings and surgical results of Duane retraction syndrome. Journal of Pediatric Ophthalmology & Strabismus. 2010 Jul 1;47(4):220-6.

Archer SM. The effect of medial versus lateral rectus muscle surgery on distance-near incomitance. Journal of American Association for Pediatric Ophthalmology and Strabismus. 2009 Feb 1;13(1):20-6.

Arnoldi KA, Tychsen L. Surgery for esotropia with a high accommodative convergence/accommodation ratio: effects on accommodative vergence and binocularity. Ophthalmic Surgery, Lasers and Imaging Retina. 1996 May 1;27(5):342-8.

Bagheri A, Adhami F, Repka MX. Bilateral recession-resection surgery for convergent strabismus fixus associated with high myopia. Strabismus. 2001 Jan 1;9(4):225-30.

Balgos JD, Amesty MA, Rodriguez AE, Al-Shymali O, Abumustafa S, Alio JL. Keratopigmentation combined with strabismus surgery to restore cosmesis in eyes with disabling corneal scarring and squint. British Journal of Ophthalmology. 2020 Jun 1;104(6):785-9.

Barbe ME, Scott WE, Kutschke PJ. A simplified approach to the treatment of Duane’s syndrome. British journal of ophthalmology. 2004 Jan 1;88(1):131-8.

Bayramlar H, Karadag R, Yildirim A, Öçal A, Sari Ü, Dag Y. Medium-term outcomes of three horizontal muscle surgery in large-angle infantile esotropia. Journal of Pediatric Ophthalmology & Strabismus. 2014 May 1;51(3):160-4.

Biedner B, Yassur Y. Effect of resection of lateral rectus muscle in undercorrected esotropia. Ophthalmologica. 1987 Mar 31;195(1):45-8.

Camuglia JE, Walsh MJ, Gole GA. Three horizontal muscle surgery for large-angle infantile esotropia: validation of a table of amounts of surgery. Eye. 2011 Nov;25(11):1435-41.

Carter D, Pujara P, Bolton K, Nicholson R. Simultaneous Development of Acute Acquired Concomitant Esotropia in Two Siblings during the COVID-19 Pandemic: A Case Report. The British and Irish Orthoptic Journal. 2023;19(1):1.

Çelik S, Inal A, Aygit ED, Ocak OB, Gökyigit B. Effect of lateral rectus muscle resection on abduction in Duane retraction syndrome type 1. International Ophthalmology. 2021 Mar;41:797-803.

Chen M, Tang S, Yan J. Assessment of surgical strategies for management of complicated strabismus reoperation in Graves’ ophthalmopathy. International Ophthalmology. 2024 Jun 25;44(1):278.

Chiu TY, Cheng MC, Wei YH, Liao SL. The role of lateral rectus muscle resection for severe esotropia after medial rectus muscle myectomy in Graves’ ophthalmopathy. European Journal of Ophthalmology. 2025 Jan;35(1):239-44.

Cogen MS, Roberts BW. Graded unilateral supramaximal medial rectus recession for moderate angle esotropia. Binocular vision & strabismus quarterly. 2006 Jan 1;21(3):147-53.

Dal Canto AJ, Crowe S, Perry JD, Traboulsi EI. Intraoperative relaxed muscle positioning technique for strabismus repair in thyroid eye disease. Ophthalmology. 2006 Dec 1;113(12):2324-30.

Damanakis AG, Arvanitis PG, Kalitsis A, Ladas ID. Bilateral medial rectus recession in convergence excess esotropia, with and without distance orthophoria. European journal of ophthalmology. 1999 Oct;9(4):297-301.

Damanakis AG, Arvanitis PG, Ladas ID, Theodossiadis GP. 8 mm bimedial rectus recession in infantile esotropia of 80-90 prism dioptres. The British Journal of Ophthalmology. 1994 Nov;78(11):842.

Dotan G, Nelson LB, Mezad-Koursh D, Stolovitch C, Cohen Y, Morad Y. Surgical outcome of strabismus surgery in patients with unilateral vision loss and horizontal strabismus. Journal of Pediatric Ophthalmology & Strabismus. 2014 Sep 1;51(5):294-8.

Du Bruyn M. Simultaneous three or four horizontal rectus muscle surgery versus two-staged surgery for large angle congenital esotropia in children: a randomized controlled trial (Doctoral dissertation).

Ela-Dalman N, Velez FG, Rosenbaum AL. Incomitant esotropia following pterygium excision surgery. Archives of Ophthalmology. 2007 Mar 1;125(3):369-73.

Erkan D, Oge I, Aritürk N, Süllü Y. Unilateral medial rectus recession for the treatment of esotropia. Annals of Ophthalmology-Glaucoma. 1997;29(6).

Eustis HS, Shah P. Accommodative esotropia treatment plan utilizing simultaneous strabismus surgery and photorefractive keratectomy. American Journal of Ophthalmology. 2018 Mar 1;187:125-9.

Farvardin H, Ahmadifar A, Farvardin H, Farvardin M. Long-term results of strabismus surgery for treatment of esotropia in patients with Möbius syndrome. Journal of American Association for Pediatric Ophthalmology and Strabismus. 2023 May 12.

Flanders M, Hastings M. Diagnosis and surgical management of strabismus associated with thyroid-related orbitopathy. Journal of Pediatric Ophthalmology & Strabismus. 1997 Nov 1;34(6):333-40.

Forrest MP, Finnigan S, Finnigan S, Gole GA. Three horizontal muscle squint surgery for large angle infantile esotropia. Clinical & Experimental Ophthalmology. 2003 Dec;31(6):509-16.

Fouad HM, Abdelhakim MA, Awadein A, Elhilali H. Comparison between medial rectus pulley fixation and augmented recession in children with convergence excess and variable-angle infantile esotropia. Journal of American Association for Pediatric Ophthalmology and Strabismus. 2016 Oct 1;20(5):405-9.

García-Basterra I, Rodríguez Del Valle JM, García-Ben A, Rodríguez Sánchez JM, García-Campos JM. Outcomes of medial rectus recession with adjustable suture in acute concomitant esotropia of adulthood. Journal of Pediatric Ophthalmology & Strabismus. 2019 Mar 19;56(2):101-6.

Garrity JA, Greninger DA, Ekdawi NS, Steele EA. The management of large-angle esotropia in Graves ophthalmopathy with combined medial rectus recession and lateral rectus resection. Journal of American Association for Pediatric Ophthalmology and Strabismus. 2019 Feb 1;23(1):15-e1.

Gigante E, Bicas HE. Monocular surgery for large-angle esotropias: a new paradigm. Arquivos Brasileiros de Oftalmologia. 2009;72:47-56.

Gigante E, Romão RA, Valério FD. Monocular surgery to correct large-angle esotropia: a 10-year follow-up study. Arquivos Brasileiros de Oftalmologia. 2018 May;81:232-8.

Goldstein JH, Sacks DB. Bilateral Duane’s syndrome. Journal of Pediatric Ophthalmology & Strabismus. 1977 Jan 1;14(1):12-7.

Greenberg MF, Pollard ZF. Poor results after recession of both medial rectus muscles in unilateral small-angle Duane’s syndrome, type I. Journal of American Association for Pediatric Ophthalmology and Strabismus. 2003 Apr 1;7(2):142-5.

Grin TR, Nelson LB. Large unilateral medial rectus recession for the treatment of esotropia. The British Journal of Ophthalmology. 1987 May;71(5):377.

Gunasekera LS, Simon JW, Zobal-Ratner J, Lininger LL. Bilateral lateral rectus resection for residual esotropia. Journal of American Association for Pediatric Ophthalmology and Strabismus. 2002 Feb 1;6(1):21-5.

Gunduz A, Ozsoy E, Ulucan PB. Duane retraction syndrome: Clinical features and a case group-specific surgical approach. In: Seminars in Ophthalmology 2019 Jan 2 (Vol. 34, No. 1, pp. 52-58). Taylor & Francis.

Habot-Wilner Z, Spierer A, Glovinsky J, Wygnanski-Jaffe T. Bilateral medial rectus muscle recession: results in children with developmental delay compared with normally developed children. Journal of American Association for Pediatric Ophthalmology and Strabismus. 2006 Apr 1;10(2):150-4.

Helveston EM, Ellis FD, Plager DA, Miller KK. Early surgery for essential infantile esotropia. Journal of Pediatric Ophthalmology & Strabismus. 1990 May 1;27(3):115-8.

Helveston EM, Neely DF, Stidham DB, Wallace DK, Plager DA, Sprunger DT. Results of early alignment of congenital esotropia. Ophthalmology. 1999 Sep 1;106(9):1716-26.

Hess JB, Calhoun JH. A new rationale for the management of large angle esotropia. Journal of Pediatric Ophthalmology & Strabismus. 1979 Nov 1;16(6):345-8.

Hilliard G, Pruett J, Donahue SP, Velez FG, Peragallo JH, Ditta L, Tavakoli M, Hoehn ME, Kuo AF, Indaram M, Kerr NC. Outcomes of Strabismus Surgery Following Teprotumumab Therapy. American Journal of Ophthalmology. 2024 Jan 6.

Hoover DL, Giangiacomo J. Results of a single lateral rectus resection for divergence and partial sixth nerve paralysis. Journal of Pediatric Ophthalmology & Strabismus. 1993 Mar 1;30(2):124-6.

Hornbrook J, Stephens A. Surgery in the management of accommodative esotropia. Australian Journal of Opthalmology. 1984 Aug;12(3):239-43.

Iida K, Goseki T, Fukaya K, Aoki T, Kuga S, Ariga C, Onouchi H, Nakano T. Efficacy of the double-dose medial rectus muscle recession technique for sagging eye syndrome with esotropia of 10-prism diopters or less at distance. Japanese Journal of Ophthalmology. 2025 Apr 25:1-8.

Infeld D, Prior C, Ryan H, O’Day J. The long-term results of surgical correction of childhood esotropia. Australian and New Zealand Journal of Ophthalmology. 1993 Feb;21(1):23-8.

Ing MR. Progressive increase in the angle of deviation in congenital esotropia. Transactions of the American Ophthalmological Society. 1994;92:117.

Jang GJ, Park MR, Park SC. Bilateral lateral rectus resection in patients with residual esotropia. Korean Journal of Ophthalmology. 2004 Dec 1;18(2):161-7.

Jo MS, Park SC. Outcomes of Bilateral Lateral Rectus Tucking in Patients with Divergence Insufficiency. Journal of the Korean Ophthalmological Society. 2023 Mar 15;64(3):245-51.

Jotterand VH, Isenberg SJ. Enhancing surgery for acquired esotropia. Ophthalmic Surgery, Lasers and Imaging Retina. 1988 Apr 1;19(4):263-6.

Kaban TJ, Smith K, Day C, Orton R, Kraft S, Cadera W. Single medial rectus recession in unilateral Duane syndrome type I. American Orthoptic Journal. 1995 Jan 1;45(1):108-14.

Kalevar A, Tone SO, Flanders M. Duane syndrome: Clinical features and surgical management. Canadian Journal of Ophthalmology. 2015 Aug 1;50(4):310-3.

Keenan JM, Willshaw HE. Outcome of strabismus surgery in congenital esotropia. British journal of ophthalmology. 1992 Jun 1;76(6):342-5.

Kim EY, Roper-Hall G, Cruz OA. Effectiveness of bilateral lateral rectus resection for residual esotropia in dysthyroid ophthalmopathy. American Journal of Ophthalmology. 2016 Nov 1;171:84-7.

Kim JA, Velez FG, Pineles SL. Strabismus surgery in patients with ocular neuromyotonia: potential unmasking of the condition and effective management tool. Journal of Neuro-Ophthalmology. 2016 Sep 1;36(3):259-63.

Kim N, Kim JH, Kim JS, Hwang JM. Möbius syndrome: clinico-radiologic correlation. Graefe’s Archive for Clinical and Experimental Ophthalmology. 2018 Nov;256:2219-23.

Köse S (Kose S), Pamukçu K, Akkin C, Cengiz H. Effect of bimedial rectus recession with a loop on the deviation in essential infantile esotropia. Strabismus. 1994 Jan 1;2(2):79-85.

Kothari M. Clinical characteristics of spontaneous late-onset comitant acute nonaccommodative esotropia in children. Indian Journal of Ophthalmology. 2007 Mar 1;55(2):117-20.

Krohel GB, Tobin DR, Harnett ME, Barrows NA. Divergence paralysis. Am J Ophthalmol 1982; 94: 506-510.

Kushner BJ, Fisher MR, Lucchese NJ, Morton GV. How far can a medial rectus safely be recessed?. Journal of Pediatric Ophthalmology & Strabismus. 1994 May 1;31(3):138-46.

Kushner BJ, Morton GV. A randomized comparison of surgical procedures for infantile esotropia. American journal of ophthalmology. 1984 Jul 1;98(1):50-61.

Lau YH, Tang EW, Lai TH, Li KK. Acute acquired esotropia during the COVID-19 pandemic: four case reports. Hong Kong Med J. 2023 Apr 1;29(2):165-7.

Lee BJ, Lim HT. High Accommodative Convergence/Accommodation Ratio Consecutive Esotropia Following Surgery for Intermittent Exotropia: Clinical Feature, Diagnosis, and Treatment. Journal of Clinical Medicine. 2021 May 15;10(10):2135.

Lee HJ, Kim SJ. Clinical characteristics and surgical outcomes of adults with acute acquired comitant esotropia. Japanese Journal of Ophthalmology. 2019 Nov;63:483-9.

Lee M, Han SH, Han J. Strabismus Surgical Outcomes in Patients with Tonic and Accommodative Convergence Excess Esotropia. Journal of the Korean Ophthalmological Society. 2018 May 1;59(5):465-70.

Lee DA, Dyer JA. Bilateral medial rectus muscle recession and lateral rectus muscle resection in the treatment of congenital esotropia. American Journal of Ophthalmology. 1983 Apr 1;95(4):528-35.

Legrand A, Bui-Quoc E, Bucci MP. Re-alignment of the eyes, with prisms and with eye surgery, affects postural stability differently in children with strabismus. Graefe’s archive for clinical and experimental ophthalmology. 2012 Jun;250:849-55.

Lekskul A, Wuthisiri W, Jarupanich N. A Prospective Study of One-Muscle Surgery in 15–25 Prism Diopters Horizontal Comitant Strabismus in Adults. Clinical Ophthalmology. 2021 Aug 31:3669-78.

Lewis JR, Malem AH, Buncic JR. Periodic alternating esotropia associated with disorders of neural tube closure. Journal of American Association for Pediatric Ophthalmology and Strabismus. 2025 May 21:104226.

Li B, Sharan S. Evaluation and surgical outcome of acquired nonaccommodative esotropia among older children. Canadian Journal of Ophthalmology. 2018 Feb 1;53(1):45-8.

Lockhart LB, Biglan AW, Hiles DA. A comparison of the effectiveness of bilateral resection to unilateral marginal myotomy and resection for correcting residual esodeviations. American Orthoptic Journal. 1986 Jan 1;36(1):49-57.

Lueder GT, Galli M. Long-term outcomes of strabismus surgery in Mobius sequence. Strabismus. 2019 Apr 3;27(2):43-6.

Lueder GT, Norman AA. Strabismus surgery for elimination of bifocals in accommodative esotropia. American journal of ophthalmology. 2006 Oct 1;142(4):632-5.

Masoomian B, Shields CL, Esfahani HR, Khalili A, Ghassemi F, Rishi P, Akbari MR, Khorrami-Nejad M. Strabismus management in retinoblastoma survivors. BMC ophthalmology. 2024 Mar 13;24(1):114.

Mavrikakis I, Pegado V, Lyons C, Rootman J. Congenital orbital fibrosis: a distinct clinical entity. Orbit. 2009 Jan 1;28(1):43-9.

McCracken MS, del Prado JD, Granet DB, Levi L, Kikkawa DO. Combined eyelid and strabismus surgery: examining conventional surgical wisdom. Journal of Pediatric Ophthalmology & Strabismus. 2008;45(4):220-4.

Merino P, Freire M, Yáñez-Merino J, de Liaño PG. Surgical outcomes of acquired acute comitant esotropia. Causes and classification. Archivos de la Sociedad Española de Oftalmología (English Edition). 2022 Oct 1;97(10):558-64.

Merino P, Merino M, De Liaño PG, Blanco N. Horizontal rectus surgery in Duane syndrome. European Journal of Ophthalmology. 2012 Mar;22(2):125-30.

Merrill K, Anderson J, Watson D, Areaux Jr RG. A cluster of cyclic esotropia: white matter changes on MRI and surgical outcomes. Journal of Pediatric Ophthalmology & Strabismus. 2019 May 22;56(3):178-82.

Michieletto P, Pensiero S, Diplotti L, Ronfani L, Giangreco M, Danieli A, Bonanni P. Strabismus surgery in Angelman syndrome: More than ocular alignment. PloS one. 2020 Nov 13;15(11):e0242366.

Millán T, Carvalho KM, Minguini N. Results of monocular surgery under peribulbar anesthesia for large-angle horizontal strabismus. Clinics. 2009;64:303-8.

Mims 3rd JL, Wood RC. A three dimensional surgical dose-response schedule for lateral rectus resections for residual congenital/infantile esotropia after large bilateral medial rectus recessions. Binocular vision & strabismus quarterly. 2000 Jan 1;15(1):20-8.

Mims III JL, Wood RC. The maximum motor fusion test: a parameter for surgery for acquired esotropia. Journal of American Association for Pediatric Ophthalmology and Strabismus. 2000 Aug 1;4(4):211-6.

Mims III JL, Wood RC. Verification and refinement of surgical guidelines for infantile esotropia: a prospective study of 40 cases. Binocular Vision. 1989;4:7-14.

Minkoff OV, Donahue SP. Three-muscle surgery for infantile esotropia in children younger than age 2 years. Journal of Pediatric Ophthalmology & Strabismus. 2005 May 1;42(3):144-8.

Mittelman D. Surgical management of adult onset age-related distance esotropia. Journal of Pediatric Ophthalmology & Strabismus. 2011 Jul 1;48(4):213-7.

Mittelman D. The surgical treatment of combined-mechanism accommodative esotropia. In: Reinecke RD (ed.). Strabismus II. New York. Grune and Stratton, 1982: pp 203-210.

Mohan A, Srivastava RM, Agrawal S. Horizontal recti-down displacement for unilateral dissociated vertical deviations–A case series. Journal of Clinical Ophthalmology and Research. 2023 Jan 1;11(1):45-7.

Molarte AB, Rosenbaum AL, von Noorden GK. Clinical Characteristics and Surgical Treatment of Intermittent Esotropia/Clinical Characteristics and Surgical Treatment of Intermittent Esotropia: Discussion. Journal of Pediatric Ophthalmology & Strabismus. 1991 May 1;28(3):137-42.

Molinari A, Plager D, Merino P, Galan MM, Swaminathan M, Ramasuramanian S, de Faber JT. Accessory extraocular muscle as a cause of restrictive strabismus. Strabismus. 2016 Oct 1;24(4):178-83.

Morad Y, Kraft SP, Mims III JL. Unilateral recession and resection in Duane syndrome. Journal of American Association for Pediatric Ophthalmology and Strabismus. 2001 Jun 1;5(3):158-63.

Muminovic I, Toffoli D, Lambert SR. Management of Restrictive Esotropia Following Pterygium Surgery. Investigative Ophthalmology & Visual Science. 2018 Jul 13;59(9):2928-.

Muralidhar R, Churawan L, Sekar M, Chidambaram AP, Mugdha P, Ramamurthy D. Outcome of delayed adjustable strabismus surgery in children using a bow-tie optional adjustable technique. Indian Journal of Ophthalmology. 2019 Feb;67(2):258.

Nabie R, Manouchehri V, Salehpour S, Ghadim BK, Bahramani E. Three horizontal muscle surgery for large-angle esotropia: success rate and dose-effect ratio. International Journal of Ophthalmology. 2020;13(4):632.

Nadeem S. Long-term outcomes of adjustable strabismus surgery at a Pakistani university hospital. International Ophthalmology. 2023 Mar;43(3):825-36.

Nelson LB. Severe adduction deficiency following a large medial rectus recession in Duane’s retraction syndrome. Archives of Ophthalmology. 1986 Jun 1;104(6):859-62.

Oatts JT, Salchow DJ. Age-related distance esotropia–fusional amplitudes and clinical course. Strabismus. 2014 Jun 1;22(2):52-7.

O’Brien CS. Surgical management of convergent strabismus. In: Allen JH (ed.). Strabismus Ophthalmic Symposium 1950. Mosby, St. Louis. Page 388-394.

O’Hara MA, Calhoun JH: Surgical correction of excess esotropia at near. J Pediatr Ophthalmol Strabismus 1990; 27:120-123.

Orton HP, Burke JP. Sensory adaptations in Duane’s retraction syndrome. Acta Ophthalmologica Scandinavica. 1995 Oct;73(5):417-20.

Peragallo JH, Velez FG, Demer JL, Pineles SL. Long-term follow-up of strabismus surgery for patients with ocular myasthenia gravis. Journal of Neuro-Ophthalmology. 2013 Mar 1;33(1):40-4.

Phillips PH, Fray KJ, Grigorian AP, Qureshi H, Spencer HJ, Rook BS. The effect of horizontal rectus muscle surgery on distance-near incomitance. American Journal of Ophthalmology. 2020 May 1;213:97-108.

Pollard ZF, Manley D. Unilateral medial rectus recession for small-angle esotropia. Archives of Ophthalmology. 1976 May 1;94(5):780-1.

Pressman SH, Scott WE. Surgical treatment of Duane’s syndrome. Ophthalmology. 1986 Jan 1;93(1):29-38.

Procianoy E, Justo DM. Results of unilateral medial rectus recession in high AC/A ratio esotropia. Journal of Pediatric Ophthalmology & Strabismus. 1991 Jul 1;28(4):212-4.

Rajavi Z, Ghadim HM, Nikkhoo M, Dehsarvi B. Comparison of hang-back and conventional recession surgery for horizontal strabismus. Journal of Pediatric Ophthalmology & Strabismus. 2001 Sep 1;38(5):273-7.

Rajavi Z, Ghadim HM, Ramezani A, Azemati M, Daneshvar F. Lateral rectus resection versus medial rectus re-recession for residual esotropia: early results of a randomized clinical trial. Clinical & Experimental Ophthalmology. 2007 Aug;35(6):520-6.

Rajavi Z, Sabbaghi H, Kheiri B, Sheibani K. Lateral rectus resection versus lateral rectus plication in patients with residual Esotropia. Strabismus. 2020 Oct 1;28(4):194-200.

Repka MX, Guyton DL. Comparison of hang-back medial rectus recession with conventional recession. Ophthalmology. 1988 Jun 1;95(6):782-7.

Roizen A, Ela-Dalman N, Velez FG, Coleman AL, Rosenbaum AL. Surgical treatment of strabismus secondary to glaucoma drainage device. Archives of Ophthalmology. 2008 Apr 1;126(4):480-6.

Roussat B, Kilani W, de Preobrajensky N, Berche M, Du Pasquier L, Paques M. Surgical management of dysthyroid diplopia with preservation of the anterior ciliary vascularization: Review of ten cases. Journal Français d’Ophtalmologie. 2015 Feb 1;38(2):118-25.

Sarici AM, Mergen B, Oguz V, Dogan C. Intraoperative relaxed muscle positioning technique results in a tertiary Center for Thyroid Orbitopathy Related Strabismus. BMC ophthalmology. 2018 Dec;18(1):1-7.

Savino G, Colucci D, Rebecchi MT, Dickmann A. Acute onset concomitant esotropia: sensorial evaluation, prism adaptation test, and surgery planning. Journal of Pediatric Ophthalmology & Strabismus. 2005 Nov 1;42(6):342-8.

Scaramuzzi M, Serafino M, Nucci P. Outcome of surgery for near angle in patient with incomitant distance/near esotropia and without manifest deviation at distance. European Journal of Ophthalmology. 2021 Nov;31(6):3394-8.

Schildwächter-von Langenthal A, Kommerell G, Klein U, Jan Simonsz H. Preoperative prism adaptation test in normosensoric strabismus. Graefe’s archive for clinical and experimental ophthalmology. 1989 May;227:206-8.

Scobee RG. Esotropia: incidence, etiology, and results of therapy. American Journal of Ophthalmology. 1951 Jun 1;34(6):817-33.

Scofield-Kaplan SM, Dunbar K, Stein G, Kazim M. Improvement in both primary and eccentric ocular alignment after thyroid eye disease-strabismus surgery with Tenon’s recession. Ophthalmic Plastic & Reconstructive Surgery. 2018 Jul 1;34(4S):S85-9.

Scott WE. Office use of prisms. In: Symposium on strabismus. Transactions of the New Orleanas Academy of Ophthalmology. St. Louis: Mosby, 1978: 91-103.

Sener EC, Mocan MC, Saraç ÖI, Gedik S, Sanaç AS. Management of strabismus in nanophthalmic patients: a long-term follow-up report. Ophthalmology. 2003 Jun 1;110(6):1230-6.

Shen T, Kang Y, Deng D, Wang Z, Qiu X, Yan J. Variant types of Duane retraction syndrome: synergistic divergences and convergences. Journal of American Association for Pediatric Ophthalmology and Strabismus. 2021 Feb 1;25(1):14-e1.

Sheppard RW, Panton CM, Smith DR. The single horizontal muscle recession operation. A survey. Canadian journal of ophthalmology. Journal canadien d’ophtalmologie. 1973 Jan 1;8(1):68-74.

Shippman S, Schudel S, Millman A, Weseley AC. Unusual Ocular Findings in Identical Twins. Journal of Pediatric Ophthalmology & Strabismus. 1988 Nov 1;25(6):298-300.

Shirabe H, Mori Y, Dogru M, Yamamoto M. Early surgery for infantile esotropia. The British Journal of Ophthalmology. 2000 May;84(5):536.

Smoot CN, Simon JW, Nelson LB. Binocularity following surgery for secondary esotropia in childhood. British journal of ophthalmology. 1990 Mar 1;74(3):155-7.

Somer D, Cinar FG, Kaderli A, Ornek F. Surgical planning and innervation in pontine gaze palsy with ipsilateral esotropia. Journal of American Association for Pediatric Ophthalmology and Strabismus. 2016 Oct 1;20(5):410-4.

Song J, Kim SK, Choi MY. Clinical characteristics and outcomes of smartphone overusers with acute acquired comitant esotropia. Journal of the Korean Ophthalmological Society. 2018 Feb 1;59(2):169-75.

Souza-Dias C, Kushner BJ, Rebouças de Carvalho LE. Long-term follow-up of cyclic esotropia. Journal of Binocular Vision and Ocular Motility. 2018 Oct 2;68(4):148-53.

Spierer A, Barak A. Strabismus surgery in children with Möbius syndrome. Journal of American Association for Pediatric Ophthalmology and Strabismus. 2000 Feb 1;4(1):58-9.

Spierer A, Barak A. Surgery for esotropia in myopic adults. Annals of Ophthalmology. 2003 Sep; 35:105-7.

Stack RR, Burley CD, Bedggood A, Elder MJ. Unilateral versus bilateral medial rectus recession. Journal of American Association for Pediatric Ophthalmology and Strabismus. 2003 Aug 1;7(4):263-7.

Sturm V, Hejcmanova M, Landau K. Effects of extraocular muscle surgery in children with monocular blindness and bilateral nystagmus. BMC ophthalmology. 2014 Dec;14:1-8.

Sturm V, Schöffler C. Long-term follow-up of children with benign abducens nerve palsy. Eye. 2010 Jan;24(1):74-8.

Sugar HS. Guides in the operative (cosmetic) treatment of nonaccommodative concomitant squint in adults. Archives of Ophthalmology. 1943 Nov 1;30(5):593-602.

Sugar HS. An evaluation of results in the use of measured recessions and resections in the correction of horizontal concomitant strabismus. American Journal of Ophthalmology. 1952 Jul 1;35(7):959-67.

Szmyd SM, Nelson LB, Calhoun JH, Spratt C. Large bimedial rectus recessions in congenital esotropia. British journal of ophthalmology. 1985 Apr 1;69(4):271-4.

Takihata Y, Mukaisho M, Kani K. Binocular function after surgical treatment of infantile esotropia. Neuro-Ophthalmology. 2001 Jan 1;26(4):235-46.

Thacker NM, Velez FG, Bhola R, Britt MT, Rosenbaum AL. Lateral rectus resections in divergence palsy: results of long-term follow-up. Journal of American Association for Pediatric Ophthalmology and Strabismus. 2005 Feb 1;9(1):7-11.

Thomas AH. Divergence insufficiency. Journal of American Association for Pediatric Ophthalmology and Strabismus. 2000 Dec 1;4(6):359-61.

Thomas S, Guha S. Large-angle strabismus: can a single surgical procedure achieve a successful outcome?. Strabismus. 2010 Dec 1;18(4):129-36.

Tjujitno AV, Prastyani R, Susanto J, Loebis R, Indriaswati L, Wulandari LR. Predictors of Success in Horizontal Strabismus Surgery: Insights from a Prospective Study. JUXTA: Jurnal Ilmiah Mahasiswa Kedokteran Universitas Airlangga. 2025 January, XVI (01): 57-62.

Tran HM, Mims III JL, Wood RC. A new dose-response curve for bilateral medial rectus recessions for infantile esotropia. Journal of American Association for Pediatric Ophthalmology and Strabismus. 2002 Apr 1;6(2):112-9.

Urist MJ. Divergence excess combined with convergence excess in the V-syndrome. American Journal of Ophthalmology. 1960 Nov 1;50(5):765-83.

Urist MJ. Recession and upward displacement of the medial rectus muscles in A-pattern esotropia. American Journal of Ophthalmology. 1968 May 1;65(5):769-73.

Urist MJ. The surgical treatment of esotropia with bilateral depression in adduction. AMA Archives of Ophthalmology. 1956 May 1;55(5):643-65.

Ventura LO, Cruz CB, Almeida HC, Millar M, Lira AF, Antunes DL. Möbius sequence: long-term strabismus surgical outcome. Arquivos Brasileiros de Oftalmologia. 2007;70:195-9.

Ventura LO, Travassos S, Ventura Filho MC, Marinho P, Lawrence L, Wilson ME, Carreiro N, Xavier V, Gois AL, Ventura CV. Congenital Zika syndrome: surgical and visual outcomes after surgery for infantile strabismus. Journal of Pediatric Ophthalmology & Strabismus. 2020 May 1;57(3):169-75.

Vodicková K, Autrata R, Šenková K, Rehurek J, Pellarová H. Results of surgery in Duane’s retraction syndrome: comparison of unilateral recession and resection versus bilateral medial rectus recessions. Scripta Medica (Brno). 2007 Feb;80(1-2):71-80.

Volk AE, Carter O, Fricke J, Herkenrath P, Poggenborg J, Borck G, Demant AW, Ivo R, Eysel P, Kubisch C, Neugebauer A. Horizontal gaze palsy with progressive scoliosis: three novel ROBO3 mutations and descriptions of the phenotypes of four patients. Molecular vision. 2011;17:1978.

Vroman DT, Hutchinson AK, Saunders RA, Wilson ME. Two-muscle surgery for congenital esotropia: rate of reoperation in patients with small versus large angles of deviation. Journal of American Association for Pediatric Ophthalmology and Strabismus. 2000 Oct 1;4(5):267-70.

Walsh HL, Aldhahwani B, Tibi C, Clauss KD, Wester ST, Cavuoto KM, Capo H. Strabismus patterns and surgical results in teprotumumab-treated thyroid eye disease: insights from a single-center study. Journal of American Association for Pediatric Ophthalmology and Strabismus. 2025 Jul 29:104275.

Wang Jia-lu., Dong Ling-yan, Cen Jie, & Kang Xiao-li. (2021). The Surgical Effects of Medial Rectus Recession on Divergence Insufficiency Esotropia. Journal of Sichuan University (Medical Sciences), 52(6), 1011-1015.

Warkad VU, Hunter DG, Dagi AF, et al. Impact of adding augmented superior rectus transpositions to medial rectus muscle recessions when treating esotropic Moebius syndrome. Am J Ophthalmol 2022;237:83-90.

Weakley DR Jr, Stager DR, Everett ME: Seven-millimeter bilateral medial rectus recessions in infantile esotropia. J Pediatr Ophthalmol Strabismus 28:113, 1991.

West CE, Repka MX. A comparison of surgical techniques for the treatment of acquired esotropia with increased accommodative convergence/accommodation ratio. Journal of Pediatric Ophthalmology & Strabismus. 1994 Jul 1;31(4):232-7.;

Wright KW, Edelman PM, McVey JH, Terry AP, Lin M. High-grade stereo acuity after early surgery for congenital esotropia. Archives of ophthalmology. 1994 Jul 1;112(7):913-9.

Yang HK, Woo SJ, Kim SJ, Hwang JM. Surgical outcomes of strabismus after iatrogenic ophthalmic artery occlusion caused by cosmetic filler injections. BMC ophthalmology. 2019 Dec;19(1):1-4.

Yoo SH, Pineles SL, Goldberg RA, Velez FG. Rectus muscle resection in Graves’ ophthalmopathy. Journal of American Association for Pediatric Ophthalmology and Strabismus. 2013 Feb 1;17(1):9-15.

## Appendix 2. Candidate studies reviewed but not included because of insufficient specification of surgical strategy, nonrepresentative case selection, surgeries other than recession or resection, surgeries for exotropia, nonsurgical therapies, or reviews or editorials

Abbasoglu OE, Sener EC, Sanac AS. Factors influencing the successful outcome and response in strabismus surgery. Eye. 1996 May;10(3):315-20.

Abdallah ME, Eltoukhi EM, Awadein AR, Zedan RH. Superior rectus transposition with medial rectus recession versus medial rectus recession in esotropic Duane retraction syndrome. Journal of Pediatric Ophthalmology & Strabismus. 2020 Sep 1;57(5):309-18.

Abdelhafez MS. A new approach to treat esotropia in patients with large V-pattern. Journal of American Association for Pediatric Ophthalmology and Strabismus {JAAPOS}. 2018 Aug 1;22(4):e46.

Abdul Qayyum MS. Post-operative Diplopia in Children with Horizontal Strabismus. Ophthalmology. 2014 Apr;12(2):121.

Acar Z, Altintas O, Ozkan B, Yılmaz Tuğan B, Turkseven Kumral E, Ercalik Y. Decrease of over-elevation in adduction after surgery of medial rectus muscles in partially refractive accommodative esotropia. European Journal of Ophthalmology. 2023 Nov 6:11206721231212766.

AGNELLO R. Adjustable sutures in strabismus surgery: A personal series of cases. Australian and New Zealand Journal of Ophthalmology. 1986 May;14(2):143-53.

Agrawal S, Singh V, Yadav A, Bangwal S, Katiyar V. Modified adjustable suture hang-back recession: description of technique and comparison with conventional adjustable hang-back recession. Indian journal of ophthalmology. 2017 Nov;65(11):1183.

Agrawal S, Srivastava RM, Yadav A. Comitant Horizontal Strabismus. Strabismus: For every Ophthalmologist. 2019:47-58.

Ahn SE, Ha SG, Kim SH. Esotropia surgery considering the angle under general anesthesia. InSeminars in Ophthalmology 2017 Nov 2 (Vol. 32, No. 6, pp. 787-792). Taylor & Francis.

Akar S, Gokyigit B, Sayın N, Demirok A, Yilmaz OF. Medial rectus Faden operations with or without recession for partially accommodative esotropia associated with a high accommodative convergence to accommodation ratio. British journal of ophthalmology. 2013 Jan 1;97(1):83-7.

Akar S. Surgical treatment for partially accommodative esotropia associated with a high accommodative convergence/accommodation ratio. OPHTHALMOLOGY. 2013 Aug.

Akbari M, Bayat R, Mirmohammadsadeghi A, Mahmoudzadeh R, Eshraghi B, Salabati M. Strabismus surgery in thyroid-associated ophthalmopathy; surgical outcomes and surgical dose responses. Journal of Binocular Vision and Ocular Motility. 2020 Oct 1;70(4):150-6.

Akbari MR, Alghurab A, Azizi E, Khorrami-Nejad M. Basic acquired nonaccommodative esotropia patients managed with surgery; a study of 2102 patients. Strabismus. 2023 Oct 2;31(4):281-9.

Akbari MR, Bahar MR, Mirmohammadsadeghi A, Bayat R, Masoumi A. Short prism adaptation test in patients with acquired nonaccommodative esotropia; clinical findings and surgical outcome. Journal of American Association for Pediatric Ophthalmology and Strabismus. 2018 Oct 1;22(5):352-5.

Akbari MR, Manouchehri V, Mirmohammadsadeghi A. Surgical treatment of Duane retraction syndrome. Journal of Current Ophthalmology. 2017 Dec 1;29(4):248-57.

Akbari MR, Mirmohammadsadeghi A, Mahmoudzadeh R, Veisi A. Management of thyroid eye disease-related strabismus. Journal of Current Ophthalmology. 2020 Jan;32(1):1.

Al Ashoor MA. Comparison of Rectus Muscle Resection and Rectus Muscle Plication for Ocular Horizontal Strabismus. Iraqi National Journal of Medicine. 2021;3(2).

Alajbegović-Halimić J. Binocular vision after concomitant alternating esotropia surgery. Medical Journal. 2011 Apr 1;17(2).

Albert DG, Lederman ME. Abnormal distance—near esotropia. Documenta Ophthalmologica. 1973 Feb;34:27-36.

Aletaha M, Bagheri A, Gholipour HM, Kheiri B. Effect of limited tenon capsule and intermuscular membranes dissection on the outcome of surgery in patients with horizontal strabismus. Strabismus. 2016 Jan 2;24(1):12-5.

Alghofaili RS, Sesma G, Khandekar R. Strabismus surgery outcomes and their determinants in patients with chronic sixth nerve palsy. Middle East African Journal of Ophthalmology. 2021 Apr;28(2):104.

Al-Hayouti H, Awadein A, Gawdat G, Elhilali H. Augmented medial rectus muscle recession versus medial rectus recession with posterior scleral fixation in partially accommodative esotropia: a randomized clinical trial. Journal of American Association for Pediatric Ophthalmology and Strabismus. 2020 Oct 1;24(5):274-e1.

Ali HM. Bimedial recti slanted recession versus bimedial recti Y-split recession for surgical management of infantile-onset esotropia. Journal of Medicine in Scientific Research. 2022 Apr 1;5(2):199.

Almahmoudi FH, Al Shamrani M, Khan AM. The use of one muscle recession for horizontal strabismus. Saudi Journal of Ophthalmology. 2018 Jul 1;32(3):200-3.

Almaradny MA, Kamel MA, Hassan ZS, Al-Badry AF. Comparative study between bimedial recti Y-split recession versus bimedial recti combined recess–resect versus bimedial recti slanted recession for surgical management of infantile-onset esotropia. The Scientific Journal of Al-Azhar Medical Faculty, Girls. 2021 Oct 1;5(4):923-8.

Alnajjar T, Sesma G, Alfreihi S. Conventional surgery versus botulinum toxin injection for the management of esotropia in children with Down syndrome. Journal of American Association for Pediatric Ophthalmology and Strabismus. 2022 Oct 1;26(5):251-e1.

AlShamlan FT, Alghazal F. Comparison of Dose Increments of Botulinum Toxin A with Surgery as Primary Treatment for Infantile Esotropia and Partially Accommodative Esotropia. Clinical Ophthalmology. 2022 Jan 1:2843-9.

AlShammari S, Alaam M, Alfreihi S. Conventional surgery versus botulinum toxin injections for partially accommodative esotropia. Journal of American Association for Pediatric Ophthalmology and Strabismus. 2022 Feb 1;26(1):16-e1.

Altınsoy HI, Gokce G, Ceylan OM, Mutlu FM. Long-term motor and sensory outcomes after surgery for infantile esotropia. Vojnosanitetski pregled. 2016;73(5):463-8.

Altintas AG, Arifoglu HB, Dal D, Simsek S. Are most sixth nerve palsies really paralytic?. Journal of Pediatric Ophthalmology & Strabismus. 2011 May 1;48(3):187-91.

Altintas AK, Yilmaz FG, Duman S. Results of classical and augmented bimedial rectus recession in infantile esotropia. Strabismus. 1999 Jan 1;7(4):227-36.

Altintas O, Acar Z, Ozkan B, Elibol O. Augmented medial rectus recession with non-absorbable suture loops is effective in the treatment of convergence excess esotropia. InSeminars in Ophthalmology 2022 Feb 17 (Vol. 37, No. 2, pp. 227-231). Taylor & Francis.

Ameri A, Akbari MR, Jaafari AR, Rajabi MT, Fard MA, Mirmohammadsadeghi A. Combining rectus muscle recessions with a central tenectomy to treat large-angle horizontal strabismus. Journal of American Association for Pediatric Ophthalmology and Strabismus. 2014 Dec 1;18(6):534-8.

Arnoldi K, Shainberg M. High AC/a ET: bifocals? Surgery? Or nothing at all?. American Orthoptic Journal. 2005 Jan 1;55(1):62-75.

Arnoult JB, Yeshurun O, Mazow ML. Comparative Study of the Surgical Management of Congenital Esotropia of 50 [PD] or Less. Journal of Pediatric Ophthalmology & Strabismus. 1976 May 1;13(3):129-31.

Arslan U, Atilla H, Erkam N. Dissociated vertical deviation and its relationship with time and type of surgery in infantile esotropia. British Journal of Ophthalmology. 2010 Jun 1;94(6):740-2.

Assaf AA. Original papers: Large bimedial rectus recession (6.5 mm or more) in the management of large-angle esotropia. Strabismus. 1997 Jan 1;5(2):59-66.

Astudillo PP, Cotesta M, Schofield J, Kraft S, Mireskandari K. The effect of achieving immediate target angle on success of strabismus surgery in children. American Journal of Ophthalmology. 2015 Nov 1;160(5):913-8.

Avellaneda-Chevrier VK, Kim HA, Parvataneni MD. Single lateral rectus resection in adult nonaccommodative esotropia. Journal of American Association for Pediatric Ophthalmology and Strabismus {JAAPOS}. 2019 Aug 1;23(4):e17-8.

Awadein A, Arfeen SA, Chougule P, Kekunnaya R. Duane—minus (Duane sine retraction and Duane sine limitation): possible incomplete forms of Duane retraction syndrome. Eye. 2021 Jun;35(6):1673-9.

Awadein A, Gouda J, Elhilali H, Arnoldi K. Convergence Excess Esotropia. Journal of Binocular Vision and Ocular Motility. 2023 Oct 2;73(4):131-59.

Awadein A, Marsh JD, Guyton DL. Nonabsorbable versus absorbable sutures in large, hang-back medial rectus muscle recessions. Journal of American Association for Pediatric Ophthalmology and Strabismus. 2016 Jun 1;20(3):206-9.

Awadein A, Sharma M, Bazemore MG, Saeed HA, Guyton DL. Adjustable suture strabismus surgery in infants and children. Journal of American Association for Pediatric Ophthalmology and Strabismus. 2008 Dec 1;12(6):585-90.

Bachar Zipori A, Spierer O, Sherwin JC, Kowal L. Why bilateral medial rectus recession fails? Factors associated with early repeated surgery. International Ophthalmology. 2020 Jan;40:59-66.

Badawi N, Hegazy K. Comparative study of Y-split recession versus bilateral medial rectus recession for surgical management of infantile esotropia. Clinical Ophthalmology. 2014 May 23:1039-45.

Badr YA, Nasef MH, Arafa ES, Ali AL. Supramaximal recession of medial recti for correction of infantile esotropia of more than 50 prism diopters. Tanta Medical Journal. 2021 Jan 1;49(1):37-41.

Bagheri A, Abbasnia E, Tavakoli M. Modified Y-split and recession of medial rectus muscles in convergence excess esotropia. European Journal of Ophthalmology. 2021 Nov;31(6):3386-93.

Bagheri A, Ale-Taha M, Abrishami M, Salour H. Effect of horizontal rectus surgery on clinical and paraclinical indices in congenital nystagmus. Journal of Ophthalmic & Vision Research. 2008 Jan;3(1):6.

Bagheri A, Babsharif B, Abrishami M, Salour H, Aletaha M. Outcomes of surgical and non-surgical treatment for sixth nerve palsy. Journal of ophthalmic & vision research. 2010 Jan;5(1):32.

Bagheri A, Veisi A, Tavakoli M. Medial rectus disinsertion for management of chronic complete sixth nerve palsy. European Journal of Ophthalmology. 2022 Sep;32(5):2622-9.

Bang GM, Martinez J, Mohney BG. Incidence and clinical characteristics of adult-onset distance esotropia. Journal of American Association for Pediatric Ophthalmology and Strabismus {JAAPOS}. 2013 Feb 1;17(1):e11.

Bartley GB, Dyer JA, Ilstrup DM. Characteristics of recession-resection and bimedial recession for childhood esotropia. Archives of Ophthalmology. 1985 Feb 1;103(2):190-5.

Basmak H, Sahin A, Yildirim N. Combined cataract and strabismus surgery with adjustable sutures. Ophthalmic Surgery, Lasers and Imaging Retina. 2006;37(3):198-203.

Bateman JB, Parks MM, Wheeler N. Discriminant analysis of congenital esotropia surgery: predictor variables for short-and long-term outcomes. Ophthalmology. 1983 Oct 1;90(10):1146-53.

Bateman JB, Parks MM. Clinical and computer-assisted analyses of preoperative and postoperative accommodative convergence and accommodation relationships. Ophthalmology. 1981 Oct 1;88(10):1024-30.

Baxter SL, Nguyen BJ, Kinori M, Kikkawa DO, Robbins SL, Granet DB. Identification and correction of restrictive strabismus after pterygium excision surgery. American journal of ophthalmology. 2019 Jun 1;202:6-14.

Bayramlar H, Ünlü C, Dag Y. Slanted medial rectus recession is effective in the treatment of convergence excess esotropia. Journal of Pediatric Ophthalmology & Strabismus. 2014 Nov 1;51(6):337-40.

Beğendi D, Duranoğlu Y. Comparison of the results of the modified and classical bi-medial hang-back recession in infantile esotropia. International Ophthalmology. 2023 Nov;43(11):4011-8.

Behrens A, Wang H, Lowery RS. Surgical outcome of unilateral medial rectus recession for congenital and acquire esotropia. Investigative Ophthalmology & Visual Science. 2018 Jul 13;59(9):2935-.

Beisse F, Koch M, Uhlmann L, Beisse C. Consideration of eyeball length and prismatic side-effects of spectacle lenses in strabismus surgery—a randomised, double-blind interventional study. Graefe’s Archive for Clinical and Experimental Ophthalmology. 2020 Jun;258:1319-26.

Bhojane VR, Bule SM, Moolchandani MM. Assessment of the surgical outcomes of esotropia in pediatric subjects with high accommodative convergence/accommodation ratio: A clinical assessment. International Journal of Health Sciences.(I):8049-56.

Bi Y, Lin S. Refractive Changes After Horizontal Strabismus Surgery. Current Eye Research. 2024 Jan 12:1-5.

Biedner B, Rothkoff L. Treatment for’A’or’V’pattern esotropia by slanting muscle insertion. British journal of ophthalmology. 1995 Sep 1;79(9):807-8.

Birch EE, Stager DR, Everett ME. Random dot stereoacuity following surgical correction of infantile esotropia. Journal of Pediatric Ophthalmology & Strabismus. 1995 Jul 1;32(4):231-5.

Bollinger KE, Kattouf V, Arthur B, Weiss AH, Kivlin J, Kerr N, West CE, Kipp M, Traboulsi EI. Hypermetropia and esotropia in myotonic dystrophy. Journal of American Association for Pediatric Ophthalmology and Strabismus. 2008 Feb 1;12(1):69-71.

Bond FM. Surgical treatment of convergent strabismus. California Medicine. 1959 Jun;90(6):433.

Bothun ED, Archer SM. Bilateral medial rectus muscle recession for divergence insufficiency pattern esotropia. Journal of American Association for Pediatric Ophthalmology and Strabismus. 2005 Feb 1;9(1):3-6.

Boulakh L, Toft-Petersen AP, Severinsen M, Toft PB, Ellervik C, Buch Hesgaard H, Heegaard S. Topical anaesthesia in strabismus surgery for Graves’ orbitopathy: a comparative study of 111 patients. Acta Ophthalmologica. 2022 Jun;100(4):447-53.

Breckenridge AL, Dickman DM, Nelson LB, Attia M, Ceyhan D. Long-term results of hang-back medial rectus recession. Journal of Pediatric Ophthalmology & Strabismus. 2003 Mar 1;40(2):81-4.

Breidenstein BG, Robbins SL, Granet DB, Acera EC. Comparison of the efficacy of medial rectus recession and lateral rectus resection for treatment of divergence insufficiency. Journal of Pediatric Ophthalmology & Strabismus. 2015 May 1;52(3):173-6.

Britt MT, Velez FG, Thacker N, Alcorn D, Foster RS, Rosenbaum AL. Surgical management of severe cocontraction, globe retraction, and pseudo-ptosis in Duane syndrome. Journal of American Association for Pediatric Ophthalmology and Strabismus. 2004 Aug 1;8(4):362-7.

Bryselbout S, Promelle V, Pracca F, Milazzo S. Clinical and surgical risk factors for consecutive exotropia. European Journal of Ophthalmology. 2019 Jan;29(1):33-7.

Bunyavee C, Archer SM, Wu CY, Del Monte MA. Comparison of unilateral and bilateral surgical approaches for the treatment of age-related divergence insufficiency esotropia. Journal of Binocular Vision and Ocular Motility. 2022 Oct 2;72(4):205-11.

Burian HM. Anomalies of the convergence and divergence functions and their treatment. New Orleans Academy of Ophthalmology Transactions. St. Louis, Mo: CV Mosby; 1971; 8: 223-232.

Burian HM. The principles of surgery on the extraocular muscles: Part I. Fundamental principles: choice of operation in concomitant strabismus: horizontal muscles. American Journal of Ophthalmology. 1950 Mar 1;33(3):380-7.

Burke JP. Distance–near disparity esotropia: can we shrink the gap?. Eye. 2015 Feb;29(2):208-13.

Cadet N, Huang PC, Superstein R, Koenekoop R, Hess RF. The effects of the age of onset of strabismus on monocular and binocular visual function in genetically identical twins. Canadian Journal of Ophthalmology. 2018 Dec 1;53(6):609-13.

Cai C, Dai H, Shen Y. Clinical characteristics and surgical outcomes of acute acquired Comitant Esotropia. BMC ophthalmology. 2019 Dec;19:1-6.

Cakmak H, Kocatürk T, Dündar SO. Comparison of surgically induced astigmatism in patients with horizontal rectus muscle recession. International Journal of Ophthalmology. 2014;7(4):709.

Calis F, Atilla H, Bingol Kiziltunc P, Alay C. Brain abnormalities in infantile esotropia as predictor for consecutive exotropia. Strabismus. 2019 Oct 2;27(4):199-204.

Can ÇÜ, Polat S, Yaşar M, İlhan B, Altıntaş AG. Ocular alignment and results of strabismus surgery in neurologically impaired children. International journal of ophthalmology. 2012;5(1):113.

Caputo AR, Guo S, Wagner RS, Picciano MV. Long term follow-up of extraocular muscle surgery for congenital esotropia. American Orthoptic Journal. 1991 Jan 1;41(1):67-71.

Caputo R, Tinelli F, Bancale A, Campa L, Frosini R, Guzzetta A, Mercuri E, Cioni G. Motor coordination in children with congenital strabismus: effects of late surgery. European Journal of Paediatric Neurology. 2007 Sep 1;11(5):285-91.

Carelli R. Essential Infantile Esotropia with Inferior Oblique hyperfunction and Lateral Recti pseudoparalysis: long term follow-up of 6 muscles approach.

Castro PD, Pedroso A, Hernández L, Naranjo RM, de Jesús Méndez T, Arias A. Results of surgery for congenital esotropia. MEDICC review. 2011 Mar 30;13(1):18-22.

Çelik S (Celik S), Inal A, Ocak OB, Aygıt ED, Akar S, Gökyiğit B. Cyclic strabismus: what measured angle of strabismus should guide surgery?. Strabismus. 2019 Oct 2;27(4):205-10.

Çelik S, Ocak OB, İnal A, Aygıt ED, Gürez C, Hüseyinhan Z, Gökyiğit B. Assessment of Refractive Error Changes and Factors for Decompensation in Patients With Fully Accommodative Esotropia. Journal of Pediatric Ophthalmology & Strabismus. 2020 Jul 1;57(4):217-23.

Cestari DM, Freire MV, Chun BY. Vertical rectus muscle recession versus combined vertical and horizontal rectus muscle recession in patients with thyroid eye disease and hypotropia. Journal of American Association for Pediatric Ophthalmology and Strabismus. 2018 Aug 1;22(4):257-61.

Çetinkaya YF, Önder F. Evaluation of the Main Factors in the Need for Reoperation in Horizontal Strabismus. Kırşehir Ahi Evran Üniversitesi Sağlık Bilimleri Dergisi. 2022 Apr 4;6(1):40-6.

Ceylan OM, Gokce G, Mutlu FM, Uludag HA, Turk A, Altinsoy HI. Consecutive exotropia: risk factor analysis and management outcomes. European Journal of Ophthalmology. 2014 Mar;24(2):153-8.

Chamberlain W. The single medial rectus recession operation. Journal of Pediatric Ophthalmology and 0Strabismus. 1970 Nov 1;7(4):208.

Chan TY, Mao AJ, Piggott JR, Makar I. Factors affecting postoperative stereopsis in acquired nonaccommodative esotropia. Canadian Journal of Ophthalmology. 2012 Dec 1;47(6):479-83.

Chandler PA. Practical Considerations Concerning Choice of Operation in Convergent Squint: Edward Jackson Memorial Lecture. American Journal of Ophthalmology. 1951 Mar 1;34(3):375-81.

Chang YH, Ryu IH, Han SH, Lee SJ, Lee JB. Intraoperative adjustment in strabismus surgery under topical anesthesia. Yonsei Medical Journal. 2006 Oct 31;47(5):667-71.

Chatzistefanou KI, Brouzas D, Droutsas KD, Koutsandrea C, Chimonidou E. Unilateral recession-resection surgery for infantile esotropia: Survival of motor outcomes and postoperative drifts. InSeminars in Ophthalmology 2018 May 19 (Vol. 33, No. 4, pp. 498-505). Taylor & Francis.

Chau VT, Hong NT, Dai NQ. The relationship of age at surgical alignment and the development of stereopsis in infantile esotropia.

Chaudhuri Z, Demer JL. Characteristics and surgical results in patients with age-related divergence insufficiency esotropia. Journal of American Association for Pediatric Ophthalmology and Strabismus {JAAPOS}. 2015 Feb 1;19(1):98-9.

Chaudhuri Z, Demer JL. Long-term surgical outcomes in the sagging eye syndrome. Strabismus. 2018 Jan 2;26(1):6-10.

Chaudhuri Z, Demer JL. Medial rectus recession is as effective as lateral rectus resection in divergence paralysis esotropia. Archives of ophthalmology. 2012 Oct 1;130(10):1280-4.

Chen Y, Ma X. Treatment of younger patients with accommodative esotropia. Researchsquare.com

Chen YY, Wei YH, Liao SL. Postoperative residual vertical deviation affects quality of life in Asian patients with thyroid-associated ophthalmopathy (Graves ophthalmopathy). Japanese Journal of Ophthalmology. 2023 Apr 20:1-9.

Choi HY, Jung JH. Bilateral lateral rectus muscle recession with medial rectus pulley fixation for divergence excess intermittent exotropia with high AC/A ratio. Journal of American Association for Pediatric Ophthalmology and Strabismus. 2013 Jun 1;17(3):266-8.

Chun BY, Freire MV, Cestari DM. Surgical responses and outcomes of bilateral medial rectus recession in esotropia with spinocerebellar ataxia. Journal of Pediatric Ophthalmology & Strabismus. 2019 Jul 1;56(4):266-70.

Chung AK, Rehman SU, Bradbury JA. Comparison of modified anchored “hang-back technique (HBT)” with conventional HBT in bimedial rectus recession. Journal of American Association for Pediatric Ophthalmology and Strabismus. 2005 Jun 1;9(3):234-9.

Cifuentes DL, Pineles SL, Demer JL, Velez FG. Surgical success and lateral incomitance following three-muscle surgery for large-angle horizontal strabismus. Journal of American Association for Pediatric Ophthalmology and Strabismus. 2018 Feb 1;22(1):17-21.

Clark AC, Nelson LB, Simon JW, Wagner RU, Rubin SE. Acute acquired comitant esotropia. British Journal of Ophthalmology. 1989 Aug 1;73(8):636-8.

Clark RA, Ariyasu R, Demer JL. Medial rectus pulley posterior fixation is as effective as scleral posterior fixation for acquired esotropia with a high AC/A ratio. American journal of ophthalmology. 2004 Jun 1;137(6):1026-33.

Clark RA, Choy AE, Demer JL. Lateral rectus sag and recurrent esotropia in children. Journal of American Association for Pediatric Ophthalmology and Strabismus. 2019 Apr 1;23(2):81-e1.

Clark RA, Demer JL. Lateral rectus superior compartment palsy. American journal of ophthalmology. 2014 Feb 1;157(2):479-87.

Clark RA, Demer JL. Magnetic resonance imaging of the globe-tendon interface for extraocular muscles: is there an “arc of contact”?. American journal of ophthalmology. 2018 Oct 1;194:170-81.

Clark RA, Isenberg SJ, Rosenbaum AL, Demer JL. Posterior fixation sutures: a revised mechanical explanation for the fadenoperation based on rectus extraocular muscle pulleys. American journal of ophthalmology. 1999 Dec 1;128(6):702-14.

Clark RA, Rosenbaum AL. Instrument-induced measurement errors during strabismus surgery. Journal of American Association for Pediatric Ophthalmology and Strabismus. 1999 Feb 1;3(1):18-25.

Clark TY, Clark RA. Medial rectus pulley posterior fixation for esotropia with a high AC/A ratio. Journal of American Association for Pediatric Ophthalmology and Strabismus {JAAPOS}. 2017 Feb 1;21(1):63-.

Clorfeine GS, Parker WT. Adjustment sensitivity of horizontal rectus muscles in adjustable strabismus surgery. Archives of Ophthalmology. 1987 Dec 1;105(12):1664-6.;

Grossniklaus HE, Reinhart WJ, Thomas SW. Adjustable Strabismus Surgery," published in the December 1987 issue of the Archives. 1987; 105: 1664.

Coats DK, Avilla CW, Lee AG, Paysse EA. Etiology and surgical management of horizontal pontine gaze palsy with ipsilateral esotropia. Journal of American Association for Pediatric Ophthalmology and Strabismus. 1998 Oct 1;2(5):293-7.

Costenbader FD, Bair DR. Strabismus surgery—monocular or binocular?. AMA Archives of Ophthalmology. 1954 Nov 1;52(5):655-63.

Crawford BA. The surgery of exophthalmic ophthalmoplegia. Medical Journal of Australia. 1969 Dec;2(24):1200-4.

Crouch ER, Dean TW, Holmes JM, Kraker RT, Miller AM, Kraus C, Gunton KB, Repka MX, Marsh JD, Del Monte MA, Luke PA. A prospective observational study of adult divergence insufficiency esotropia. Journal of American Association for Pediatric Ophthalmology and Strabismus {JAAPOS}. 2019 Aug 1;23(4):e7.

Crouch ER, Dean TW, Kraker RT, Miller AM, Kraus CL, Gunton KB, Repka MX, Marsh JD, Del Monte MA, Luke PA, Peragallo JH. A prospective study of treatments for adult-onset divergence insufficiency–type esotropia. Journal of American Association for Pediatric Ophthalmology and Strabismus. 2021 Aug 1;25(4):203-e1.

Dagi LR, Elhusseiny AM. Adjustable graded augmentation of superior rectus transposition for treatment of abducens nerve palsy and Duane syndrome. Journal of American Association for Pediatric Ophthalmology and Strabismus. 2020 Oct 1;24(5):268-e1.

Damanakis A, Ikonomopoulos N, Alatsaki M, Arvanitis P. Effect of posterior fixation sutures on the accommodative element of partially accommodative strabismus. Ophthalmologica. 1994 Apr 1;208(2):71-6.

Daniel SM, Flanders M. Pontine Lesions And Abducens Nerve Palsy: clinical Presentation And Surgical Management. Investigative Ophthalmology & Visual Science. 2011 Apr 22;52(14):6367-.

Dankner SR, Mash AJ, Jampolsky A. Intentional surgical overcorrection of acquired esotropia. Archives of ophthalmology. 1978 Oct 1;96(10):1848-52.

Daphna MZ, Tomer ZB, Ofira Z, Sharon BM, Chaim S. Refractive Changes Induced by Recession of the Medial Rectus and the Infe-rior Oblique Muscles. J Ophthalmic Clin Res. 2019;6:053.

Dass SE, Cheng M, Bahl RS. Surgical outcomes for esotropia in children with high accommodative convergence/accommodation ratio. Indian Journal of Ophthalmology. 2021 Oct;69(10):2766.

de Alba Campomanes AG, Binenbaum G, Eguiarte GC. Comparison of botulinum toxin with surgery as primary treatment for infantile esotropia. Journal of American Association for Pediatric Ophthalmology and Strabismus. 2010 Apr 1;14(2):111-6.

de Castro-Abeger A, Benegas NM, Kushner B, Donahue SP. Horizontal transposition of the vertical rectus muscles to correct a head tilt in 5 patients with idiopathic nystagmus syndrome. American Journal of Ophthalmology. 2020 Sep 1;217:68-73.

de Liaño Sánchez PG, González GO, Sanz PM, Villafruela JE. Age-related distance esotropia: Clinical features and therapeutic outcomes. Archivos de la Sociedad Española de Oftalmología (English Edition). 2016 Dec 1;91(12):561-6.

de Molina AC, Muñoz ML. Ocular alignment under general anesthesia in congenital esotropia. Journal of Pediatric Ophthalmology & Strabismus. 1991 Sep 1;28(5):278-82.

deB Ribeiro G, Brooks SE, Archer SM, Del Monte MA. Vertical shift of the medial rectus muscles in the treatment of A-pattern esotropia: analysis of outcome. Journal of Pediatric Ophthalmology & Strabismus. 1995 May 1;32(3):167-71.

Deguchi M, Yokoyama T, Kamihata K, Matsuzaka Y, Kawanami K, Hosoi S, Tanaka H. A Study of Horizontal Strabismus Surgery: Is Medial Rectus Surgery More Effective on Near Deviation?. In: Strabismus and Ocular Motility Disorders: Proceedings of the Sixth Meeting of the International Strabismological Association Surfers Paradise, Australia, 1990 1990 (pp. 487-492). Macmillan Education UK.

Deller M. Why should surgery for early-onset strabismus be postponed?. British journal of ophthalmology. 1988 Feb 1;72(2):110-5.

Demer JL, Clark RA. Axis for Horizontal Ocular Rotation Is Unchanged by Surgical Correction of Esotropia. Investigative Ophthalmology & Visual Science. 2023 Jun 1;64(8):5320-.

Demer JL, Clark RA. The medial rectus is the bad actor in intermittent esotropia. Journal of American Association for Pediatric Ophthalmology and Strabismus {JAAPOS}. 2019 Aug 1;23(4):e7.

Dickmann A, Petroni S, Salerni A, Parrilla R, Savino G, Battendieri R, Perrotta V, Radini C, Balestrazzi E. Effect of vertical transposition of the medial rectus muscle on primary position alignment in infantile esotropia with A-or V-pattern strabismus. Journal of American Association for Pediatric Ophthalmology and Strabismus. 2011 Feb 1;15(1):14-6.

Dimitrova G, Mihaylova B. Comparison between traditional bilateral and large unilateral medial rectus muscle recession for moderate esotropia. Bulgarian Review of Ophthalmology. 2016 Oct 1;60(4):11-4.

Docherty PT. Paralytic strabismus correction by adjustable suture technique. British journal of ophthalmology. 1984 May 1;68(5):353-9.

Dogan M, Gökyigit B, Akar S, Yilmaz ÖF. Effects of ocular parameters on medial rectus faden operation with recession for esotropia. Journal of Biosciences and Medicines. 2019 Feb 2;7(02):54.

Dohvoma VA, Ebana Mvogo SR, Ndongo JA, Mvilongo CT, Ebana Mvogo C. Outcome of esotropia surgery in 2 tertiary hospitals in Cameroon. Clinical Ophthalmology. 2020 Feb 13:449-54.

Dotan G, Klein A, Ela-Dalman N, Shulman S, Stolovitch C. The efficacy of asymmetric bilateral medial rectus muscle recession surgery in unilateral, esotropic, type 1 Duane syndrome. Journal of American Association for Pediatric Ophthalmology and Strabismus. 2012 Dec 1;16(6):543-7.

Dotan G, Miller M, Strominger MB, Nelson LB. Surgical management of nystagmus. Journal of Pediatric Ophthalmology & Strabismus. 2018 Sep 20;55(5):280-3.

Dubinsky-Pertzov B, Einan-Lifshitz A, Pras E, Hartstein ME, Morad Y. Routine use of non-absorbable sutures in bi-medial rectus recession as a measure to reduce the incidence of consecutive exotropia. Eye. 2022 Sep;36(9):1772-6.

Dunnington JH, Regan EF. Factors influencing the postoperative results in concomitant convergent strabismus. AMA Archives of Ophthalmology. 1950 Dec 1;44(6):813-22.

Dunnington JH, Wheeler MC. Operative results in two hundred and eleven cases of convergent strabismus. Archives of Ophthalmology. 1942 Jul 1;28(1):1-1.

Durrani A, Morrison D, Donahue S. Maximal tolerated near angle surgery for divergence insufficiency type esotropia. Journal of American Association for Pediatric Ophthalmology and Strabismus {JAAPOS}. 2012 Feb 1;16(1):e15.

Duss DN, Guo S, Wang FM, Wagner RS. Management of Divergence Insufficiency Esotropia. Journal of Pediatric Ophthalmology & Strabismus. 2020 Jan 1;57(1):4-6.

Eckstein A, Esser J, Oeverhaus M, Saeed P, Jellema HM. Surgical treatment of diplopia in Graves orbitopathy patients. Ophthalmic Plastic & Reconstructive Surgery. 2018 Jul 1;34(4S):S75-84.

Eckstein A, Esser J. Surgical management of extraocular muscle dysfunction in patients with GO. Graves’ Disease: A Comprehensive Guide for Clinicians. 2015:287-99.

Edelman PM, Brown MH, Murphree AL, Wright KW. Consecutive esodeviation… then what?. American Orthoptic Journal. 1988 Jan 1;38(1):111-6.

Edelman PM, Lingua RW, Azen SP. A Comparison of the Effect of Symmetrical vs. Asymmetrical Esotropia Surgery on the AC/A Ratio and on the Distance-Near Disparity. American Orthoptic Journal. 1986 Jan 1;36(1):58-64.

Edwards WC, Moran CT, Askew W. Statistical analysis of esotropia surgery. Journal of Pediatric Ophthalmology & Strabismus. 1973 Nov 1;10(4):256-66.

Ehrenberg M, Nihalani BR, Cain CE, Melvin P, Dagi LR. Goal-determined, risk-stratified assessment of esotropia surgery outcomes. Journal of American Association for Pediatric Ophthalmology and Strabismus {JAAPOS}. 2012 Feb 1;16(1):e15.

El Zahar WS, Shafik HM, Awara AM, El Desouky MA. Palpebral fissure changes following horizontal recti muscles surgery. Tanta Medical Journal. 2023 Jan 1;51(1):6-12.

Elkamshoushy A, Kassem A. Stepped strabismus surgery. Clinical Ophthalmology. 2021 Apr 28:1783-9.

Elkamshoushy A, Langue MA. Outcomes of bilateral lateral rectus recession in treatment of recurrent exotropia after bilateral medial rectus resection. European Journal of Ophthalmology. 2019 Jul;29(4):402-5.

ElKamshoushy AA. Bilateral medial rectus resection for primary large-angle exotropia. Journal of American Association for Pediatric Ophthalmology and Strabismus. 2017 Apr 1;21(2):112-6.

Ellis Jr GS, Pritchard CH, Baham L, Babiuch A. Medial rectus surgery for convergence excess esotropia with an accommodative component: a comparison of augmented recession, slanted recession, and recession with posterior fixation. American Orthoptic Journal. 2012 Jan 1;62(1):50-60.

Elsaadani IA, Eleiwa T, Khater AA. Anchored absorbable versus conventional nonabsorbable sutures in hang-back medial rectus muscle recession for large-angle esotropia. Delta Journal of Ophthalmology. 2022 Apr 1;23(2):125-30.

Elsas FJ, Mays A. Augmenting surgery for sensory esotropia with near/distance disparity with a medial rectus posterior fixation suture. Journal of Pediatric Ophthalmology & Strabismus. 1996 Jan 1;33(1):28-30.

Elsayed SH, Basyoni OE, El-Marakby MA, Attia RS. Medial rectus recession versus lateral rectus advancement in surgical correction of consecutive esotropia. Delta Journal of Ophthalmology. 2022 Jul 1;23(3):206-12.

El-Zawahry WM. Refractive error changes after bilateral medial rectus muscle recession surgery in congenital esotropia. Journal of the Egyptian Ophthalmological Society. 2017 Jul 1;110(3):100-4.

Engel JM, Guyton DL, Hunter DG. Adjustable sutures in children. Journal of American Association for Pediatric Ophthalmology and Strabismus. 2014 Jun 1;18(3):278-84.

Engel JM, Rousta ST. Adjustable sutures in children using a modified technique. Journal of American Association for Pediatric Ophthalmology and Strabismus. 2004 Jun 1;8(3):243-8.

Eustis HS, Shah P. Accommodative esotropia (AET) treatment utilizing simultaneous strabismus surgery and photorefractive keratoplasty. Journal of American Association for Pediatric Ophthalmology and Strabismus {JAAPOS}. 2015 Aug 1;19(4):e43.

Fard MA, Ghahvehchian H. Lateral rectus advancement versus medial rectus recession for consecutive esotropia. European Journal of Ophthalmology. 2021 Jan;31(1):258-62.

Farid MF, Elbarky AM, Saeed AM. Superior rectus and lateral rectus muscle union surgery in the treatment of myopic strabismus fixus: three sutures versus a single suture. Journal of American Association for Pediatric Ophthalmology and Strabismus. 2016 Apr 1;20(2):100-5.

Farid MF, Elbarky AM, Saeed AM. Superior rectus and lateral rectus muscle union surgery in the treatment of myopic strabismus fixus: three sutures versus a single suture. Journal of American Association for Pediatric Ophthalmology and Strabismus. 2016 Apr 1;20(2):100-5.

Farid MF, Mahmoud MR, Awwad MA. Management of stretched scar–induced secondary strabismus. BMC ophthalmology. 2020 Dec;20(1):1-6.

Farid MF, Saeed AM. Bimedial faden recession versus augmented medial rectus recession in the treatment of high ac/a ratio partially accommodative esotropia with large distant near disparity. Journal of the Egyptian Ophthalmological Society. 2016 Jan 1;109(1):10-5.

Farvardin H, Farvardin M, Koohestani S. Combined Lateral Rectus Myectomy and Maximal Medial Rectus Resection in Complete Third Cranial Nerve Palsy. Medical Hypothesis, Discovery and Innovation in Ophthalmology. 2018;7(2):83.

Farvardin M, Rad AH, Ashrafzadeh A. Results of bilateral medial rectus muscle recession in unilateral esotropic Duane syndrome. Journal of American Association for Pediatric Ophthalmology and Strabismus. 2009 Aug 1;13(4):339-42.

Felius J, Stager Jr DR, Beauchamp GR, Stager DR. Re-recession of the medial rectus muscles in patients with recurrent esotropia. Journal of American Association for Pediatric Ophthalmology and Strabismus. 2001 Dec 1;5(6):381-7.

Felius J, Stager Jr DR, Beauchamp GR, Stager DR. Re-recession of the medial rectus muscles in patients with recurrent esotropia. Journal of American Association for Pediatric Ophthalmology and Strabismus. 2001 Dec 1;5(6):381-7.

Fells P, Kousoulides L, Pappa A, Munro P, Lawson J. Extraocular muscle problems in thyroid eye disease. Eye. 1994 Sep;8(5):497-505.

Fells P. The treatment of non-comitant strabismus. In: Strabismus Symposium Amsterdam, September 3–4, 1981 1982 (pp. 197-207). Dordrecht: Springer Netherlands.

Fells PE. Management of paralytic strabismus. The British journal of ophthalmology. 1974 Mar;58(3):255.

Filipovic T. The accommodative convergence/accommodation (AC/A) and near convergence/distance (NC/D) ratios in esotropia. Journal of Pediatric Ophthalmology & Strabismus. 1998 Mar 1;35(2):91-5.

Fison PN, Chignell AH. Diplopia after retinal detachment surgery. British journal of ophthalmology. 1987 Jul 1;71(7):521-5.

Fletcher MC, Silverman SJ, Abbott W, Girard LJ, Guber D, Tomlinson E. Results of biostatistical study of the management of alternating esotropia with and without orthoptics. American Orthoptic Journal. 1969 Jan 1;19(1):31-9.

Folk ER, Miller MT, Chapman L. Consecutive exotropia following surgery. British Journal of Ophthalmology. 1983 Aug 1;67(8):546-8.

Freedman SF. Strabismus associated with glaucoma and glaucoma surgery. American Orthoptic Journal. 2001 Jan 1;51(1):2-10.

Freeley DA, Nelson LB, Calhoun JH. Recurrent esotropia following early successful surgical correction of congenital esotropia. Journal of Pediatric Ophthalmology & Strabismus. 1983 Mar 1;20(2):68-71.

Fu JJ, Hsieh MW, Lee LC, Chen PL, Wen LY, Chen YH, Chien KH. A novel method ensuring an immediate target angle after horizontal strabismus surgery in children. Frontiers in Medicine. 2022 Feb 24;9:791068.

Gaballah K. Adjustable strabismus surgery versus conventional surgery in esotropia. Journal of the Egyptian Ophthalmological Society. 2018 Oct 1;111(4):153-7.

Gaballah KA, Shawky D. Treatment modalities in Duane’s Retraction Syndrome. International Journal of Ophthalmology. 2020;13(2):278.

Galetta SL, Smith JL. Chronic isolated sixth nerve palsies. Archives of neurology. 1989 Jan 1;46(1):79-82.

Galli M, Lueder GT. Persistently recurrent infantile esotropia. American Orthoptic Journal. 2010 Jan 1;60(1):95-100.

Ganesh A, Pirouznia S, Ganguly SS, Fagerholm P, Lithander J. Consecutive exotropia after surgical treatment of childhood esotropia: a 40-year follow-up study. Acta ophthalmologica. 2011 Nov;89(7):691-5.

Gazit I, Or L, Pras E, Morad Y. The Routine Use of Nonabsorbable Sutures in Bilateral Horizontal Rectus Recession. Ophthalmology. 2024 Jan 1;131(1):124-6.

Ghali MA. Adult divergence insufficiency esotropia: a comparison of lateral rectus resection, medial rectus recession, and miniplication of lateral rectus. Journal of the Egyptian Ophthalmological Society. 2017 Jul 1;110(3):105-8.

Ghali MA. Bimedial rectus muscle elongation versus bimedial rectus muscle recession for the surgical treatment of large-angle infantile esotropia. Clinical Ophthalmology. 2017 Oct 17:1877-81.

Ghali MA. Combined resection–recession versus combined recession–retroequatorial myopexy of medial rectus muscles for treatment of near-distance disparity Esotropia. Clinical Ophthalmology. 2017 Jun 6:1065-8.

Gharabaghi D, Zanjani LK. Comparison of results of medial rectus muscle recession using augmentation, Faden procedure, and slanted recession in the treatment of high accommodative convergence/accommodation ratio esotropia. Journal of Pediatric Ophthalmology & Strabismus. 2006 Mar 1;43(2):91-4.

Ghasia F, Brunstrom-Hernandez J, Tychsen L. Repair of strabismus and binocular fusion in children with cerebral palsy: gross motor function classification scale. Investigative ophthalmology & visual science. 2011 Sep 1;52(10):7664-71.

Gibson GG. Surgical principles of concomitant convergent strabismus. American Journal of Ophthalmology. 1951 Oct 1;34(10):1431-7.

Gilbert AL, Hunter DG. Duane syndrome with prominent oculo-auricular phenomenon. Journal of American Association for Pediatric Ophthalmology and Strabismus. 2017 Apr 1;21(2):165-7.

Gillies WE, Hughes AN. Results in 50 cases of strabismus after graduated surgery designed by A scan ultrasonography. British journal of ophthalmology. 1984 Nov 1;68(11):790-5.

Gillies WE. The results of surgical treatment of esotropia. Australian Journal of Opthalmology. 1974 Feb;2(1):20-3.

Gobin MH. Causes and treatment of consecutive exotropia. In: Strabismus Symposium Amsterdam, September 3–4, 1981 1982 (pp. 123-130). Dordrecht: Springer Netherlands.

Gobin MH. Surgical management of Duane’s syndrome. The British Journal of Ophthalmology. 1974 Mar;58(3):301.

Gogate PM, Rishikeshi N, Taras S, Aghor M, Deshpande MD. Clinical audit of horizontal strabismus surgery in children in Maharashtra, India. Strabismus. 2010 Mar 1;18(1):13-7.

Goncu T, Dilmen F, Ali AK, Adibelli FM, Cakmak S. The factors affecting surgical success rate for the patients with congenital esotropia. The European Research Journal. 2015 Apr 7;1(2):33-8.

Gong Q, Wei H, Zhou X, Li Z, Liu L. Risk factors analysis of consecutive exotropia: Oblique muscle overaction may play an important role. Medicine. 2016 Dec;95(50).

Good WV, Da Sa LC, Lyons CJ, Hoyt CS. Monocular visual outcome in untreated early onset esotropia. British journal of ophthalmology. 1993 Aug 1;77(8):492-4.

Grace S, Cavuoto K, Capo H. Characteristics and Surgical Management of Divergence Insufficiency. Investigative Ophthalmology & Visual Science. 2014 Apr 30;55(13):2592-.

Grace SF, Cavuoto KM, Shi W, Capo H. Surgical treatment of adult-onset esotropia: characteristics and outcomes. Journal of Pediatric Ophthalmology & Strabismus. 2017 Mar 1;54(2):104-11.

Graeber CP, Hunter DG. Changes in lateral comitance after asymmetric horizontal strabismus surgery. JAMA ophthalmology. 2015 Nov 1;133(11):1241-6.

Gräf M, Lorenz B. Recess–resect surgery with myopexy of the lateral rectus muscle to correct esotropia with high myopia. British Journal of Ophthalmology. 2015 Dec 1;99(12):1702-5.

Gray ME, Motley WW, Melson A, Salisbury S. Outcomes of strabismus surgery for esotropia in children with Down syndrome compared with matched controls. Journal of American Association for Pediatric Ophthalmology and Strabismus {JAAPOS}. 2011 Feb 1;15(1):e19.

Greenwald MJ, Eagle JR, Peters C, Haldi BA. Treatment of acquired esotropia: for augmented surgery. American Orthoptic Journal. 1998 Jan 1;48(1):16-20.

Greninger D, Berg P, Steele E. Treatment of esotropia from thyroid eye disease by lateral rectus resection. Investigative Ophthalmology & Visual Science. 2013 Jun 16;54(15):5905-.

Gündüz A (Gunduz A), Öztürk E, Özsoy E, Güntürkün PN. Infantile Esotropia: Clinical Features and Results of Bilateral Medial Rectus Recession. Istanbul Medical Journal. 2023 May 1;24(2).

Gupta AK, Bajracharya K, Yadav RD, Rai SK. Comparison of bilateral medial rectus recession vs recession-resection as surgery for infantile esotropia. Asian Journal of Medical Sciences. 2021 Aug 1;12(8).

Gürez C, Çakmak Tuğcu B, Yiğit U, Özdemir S, Ağaçhan A. The evaluation of the development of binocular functions in infantile esotropia after surgical treatment. https://jag.journalagent.com/z4/vi.asp?pdir=tjo&plng=eng&un=TOG-62207&look4=

Gurez C, Ocak OB, Inal A, Aygit ED, Celik S, Huseyinhan Z, Demirok A, Gokyigit B. Surgical dose–response relationship in patients with down syndrome with esotropia; a comparative study. Journal of American Association for Pediatric Ophthalmology and Strabismus {JAAPOS}. 2018 Aug 1;22(4):e45-6.

Gurland J, Vagge A, Nelson LB. One-muscle strabismus surgery: a review. Journal of Pediatric Ophthalmology & Strabismus. 2018 Sep 20;55(5):288-92.

Habot-Wilner Z, Spierer A, Barequet IS, Wygnanski-Jaffe T. Long-term results of esotropia surgery in children with developmental delay. Journal of American Association for Pediatric Ophthalmology and Strabismus. 2012 Feb 1;16(1):32-5.

Hakim OM, El-Hag YG, Haikal MA. Releasable adjustable suture technique for children. Journal of American Association for Pediatric Ophthalmology and Strabismus. 2005 Aug 1;9(4):386-90.

Hakim OM, El-Hag YG, Haikal MA. Strabismus surgery under augmented topical anesthesia. Journal of American Association for Pediatric Ophthalmology and Strabismus. 2005 Jun 1;9(3):279-84.

Hamed LM, Lingua RW, Fanous MM, Saunders TG, Lusby FW. Synergistic divergence: saccadic velocity analysis and surgical results. Journal of Pediatric Ophthalmology & Strabismus. 1992 Jan 1;29(1):30-7.

Han SY, Han J, Rhiu S, Lee JB, Han SH. Risk factors for consecutive exotropia after esotropia surgery. Japanese journal of ophthalmology. 2016 Jul;60:333-40.

Hao R, Zhao K, Zhang W. Resolution of hypertropia with correction of consecutive horizontal deviation. Journal of the Chinese Medical Association. 2017 Jul 1;80(7):458-61.

Harley RD. Paralytic strabismus in children: etiologic incidence and management of the third, fourth, and sixth nerve palsies. Ophthalmology. 1980 Jan 1;87(1):24-43.

Harrison A, Allen L, O’Connor A. Strabismus surgery for Esotropia, Down syndrome and developmental delay; is an altered surgical dose required? A literature review. The British and Irish orthoptic journal. 2020;16(1):4.

Hasebe S, Morisawa S, Furuse T, Kobashi R. Double-angle Modification of Parks Surgical Table Improves the Outcomes of Unilateral Medial Rectus Recession in Adult Patients with Small-angle Esotropia. Investigative Ophthalmology & Visual Science. 2018 Jul 13;59(9):2936-.

Hassan AA. Clinical Variability and Treatment Outcomes of Pediatric Esotropia at Sohag University Hospital. Sohag Medical Journal. 2020 Jul 1;24(3):138-46.

Hassan MB, Diehl NN, Mohney BG. Immediate postoperative alignment following bimedial rectus recession for esotropia in children compared to adults. Journal of Pediatric Ophthalmology & Strabismus. 2018 Sep 20;55(5):299-305.

HE, Cao, and Luo Xiao-ling. “Surgical outcome of extra-large recession of unilateral medial rectus for small angle esotropia in adults.” International Eye Science (2017): 2370-2372.

Heede S, Salchow D. Comparison of hang-back medial rectus recession with conventional recession for the correction of esotropia in children. Klinische Monatsblatter fur Augenheilkunde. 2014 Oct 21;231(10):988-93.

Heidary G, Mackinnon S, Elliott A, Barry BJ, Engle EC, Hunter DG. Outcomes of strabismus surgery in genetically confirmed congenital fibrosis of the extraocular muscles. Journal of American Association for Pediatric Ophthalmology and Strabismus. 2019 Oct 1;23(5):253-e1.

Helveston EM. ‘Ultra-Early ‘Surgery for Essential Infantile Esotropia. In: Strabismus and Ocular Motility Disorders: Proceedings of the Sixth Meeting of the International Strabismological Association Surfers Paradise, Australia, 1990 1990 (pp. 325-333). Macmillan Education UK.

Helveston EM. Surgery for esotropia. International Ophthalmology. 1983 Jan;6:55-9.

Helveston EM. Surgical Management of Strabismus: An Atlas of Strabismus Surgery, 5th edn. Wayenborgh Publishing: Belgium, 2005, pp 109–110.

Helveston EM. Surgical treatment of cyclic esotropia. American Orthoptic Journal. 1976 Jan 1;26(1):87-8.

Hemmerdinger C, Rowe N, Baker L, Lloyd IC. Bimedial hang-back recession—outcomes and surgical response. Eye. 2005 Nov;19(11):1178-81.

Hendler K, Pineles SL, Demer JL, Yang D, Velez FG. Adjustable augmented rectus muscle transposition surgery with or without ciliary vessel sparing for abduction deficiencies. Strabismus. 2014 Jun 1;22(2):74-80.

Hennein L, Moore A. Surgical treatment of children with cyclic esotropia. Investigative Ophthalmology & Visual Science. 2020 Jun 10;61(7):517-.

Herlihy EP, Phillips JO, Weiss AH. Esotropia greater at distance: children vs adults. JAMA ophthalmology. 2013 Mar 1;131(3):370-5.

Hermann JS, BURIAN HM. Accommodative esotropia: classification and treatment. International ophthalmology clinics. 1971 Dec 1;11(4):23-6.

Hertle RW, Granet DB, Schaffer MA, Wilson MC. Adjustable horizontal rectus recession surgery for disparate distance-near ocular deviations. Strabismus. 1997 Jan 1;5(3):109-15.

Hinterhuber L, Rezar-Dreindl S, Schmidt-Erfurth U, Stifter E. Postoperative outcome and influencing factors of strabismus surgery in infants aged 1–6 years. Graefe’s Archive for Clinical and Experimental Ophthalmology. 2024 Feb 16:1-9.

Hoehn ME, Calderwood J, O’Donnell T, Armstrong GT, Gajjar A. Children with dorsal midbrain syndrome as a result of pineal tumors. Journal of American Association for Pediatric Ophthalmology and Strabismus. 2017 Feb 1;21(1):34-8.

Hollander J, Eldweik L, Ledoux D. Surgical strabismus in Down syndrome patients. Investigative Ophthalmology & Visual Science. 2015 Jun 11;56(7):5225-.

Höllhumer R, Vallabh B, Carmichael T. Retrospective review of ocular alignment after large-angle congenital esotropia surgery. African Vision and Eye Health. 2015 Jul 17;74(1):7.

Holman RE, Merritt JC, Biglan AW. Infantile Esotropia: Results in the Neurologic Impaired and “Normal” Child at NCMH (Six Years)/DISCUSSION. Journal of Pediatric Ophthalmology & Strabismus. 1986 Jan 1;23(1):41-5.

Holmes JM, Leske DA, Christiansen SP. Initial treatment outcomes in chronic sixth nerve palsy. Journal of American Association for Pediatric Ophthalmology and Strabismus. 2001 Dec 1;5(6):370-6.

Holmes JM. Evaluation and Management of Divergence Insufficiency-Type Esotropia. Journal of Binocular Vision and Ocular Motility. 2022 Oct 2;72(4):230-3.

Honglertnapakul W, Cavuoto KM, McKeown CA, Capó H. Surgical treatment of strabismus in thyroid eye disease: Characteristics, dose–response, and outcomes. Journal of American Association for Pediatric Ophthalmology and Strabismus. 2020 Apr 1;24(2):72-e1.

Hopker LM, Weakley DR. Surgical results after one-muscle recession for correction of horizontal sensory strabismus in children. Journal of American Association for Pediatric Ophthalmology and Strabismus. 2013 Apr 1;17(2):174-6.

Hsieh MW, Hsu CK, Kuo PC, Chang HC, Chen YH, Chien KH. Factors Predicting the Success of Combined Orbital Decompression and Strabismus Surgery in Thyroid-Associated Orbitopathy. Journal of Personalized Medicine. 2022 Jan 31;12(2):186.

Huang LJ, Wu YY, Li ND. Y-splitting medial rectus muscle and recession in treatment for convergence excess esotropia. International Journal of Ophthalmology. 2022;15(4):661.

Huang X, Meng Y, Hu X, Zhao Y, Ye M, Yi B, Zhou L. The effect of different treatment methods on acute acquired concomitant esotropia. Computational and Mathematical Methods in Medicine. 2022 Apr 23;2022.

Huddleston SM, Kerr NC, Fleming JC. Proptosis and Orbital Decompression Prior to Strabismus Surgery in Patients with Graves Ophthalmopathy: Influence on Postoperative Shift in Eye Alignment. Investigative Ophthalmology & Visual Science. 2011 Apr 22;52(14):6348-.

Huelin FJ, Del Valle JM, Sales-Sanz M, Ye-Zhu C, Díaz-Montealegre A, Muñoz-Negrete FJ. Postoperative drift and dose-response of strabismus surgery in thyroid eye disease: predicting desired outcomes with intraoperative adjustable sutures. Canadian Journal of Ophthalmology. 2022 Nov 17.

Huston PA, Hoover DL. Surgical outcomes following rectus muscle plication versus resection combined with antagonist muscle recession for basic horizontal strabismus. Journal of American Association for Pediatric Ophthalmology and Strabismus. 2018 Feb 1;22(1):7-11.

Hwang B, Heo H, Lambert SR. Risk factors for reoperation after strabismus surgery among patients with thyroid eye disease. American Journal of Ophthalmology. 2022 Jun 1;238:10-5.

Hwang JM, Min BM, Park SC, Oh SY, Sung NK. A randomized comparison of prism adaptation and augmented surgery in the surgical management of esotropia associated with hypermetropia: one-year surgical outcomes. Journal of American Association for Pediatric Ophthalmology and Strabismus. 2001 Feb 1;5(1):31-4.

Hyndman J. Spasm of the near reflex: literature review and proposed management strategy. Journal of binocular vision and ocular motility. 2018 Jul 3;68(3):78-86.

Icazbalceta RE, Urdapilleta M, Zermeno A, Vargas J, Murillo C. Amblyopia as a risk factor for recurrence in patients with infantile esotropia treated surgically: 10-year follow-up. Investigative Ophthalmology & Visual Science. 2018 Jul 13;59(9):1027-.

Ing MR, Norcia A, Stager Sr D, Black B, Hoffman R, Mazow M, Troia S, Scott W, Lambert S. A prospective study of alternating occlusion before surgical alignment for infantile esotropia: one-year postoperative motor results. Journal of American Association for Pediatric Ophthalmology and Strabismus. 2006 Feb 1;10(1):49-53.

Ing MR. Early surgical alignment for congenital esotropia. Transactions of the American Ophthalmological Society. 1981;79:625.

Ing MR. Outcome Study of Surgical Alignment before Sup Months of Age for Congenital Esotropia. Ophthalmology. 1995 Dec 1;102(12):2041-5.

Ing MR. Progressive increase in the quantity of deviation in congenital esotropia. Ophthalmic Surgery, Lasers and Imaging Retina. 1996 Jul 1;27(7):612-7.

Ing MR. Surgical alignment prior to six months of age for congenital esotropia. Transactions of the American Ophthalmological Society. 1995;93:135.

Isaac CR, Chalita MR. Effect of combining oblique muscle weakening procedures with bimedial rectus recessions on the surgical correction of esotropia. Journal of American Association for Pediatric Ophthalmology and Strabismus. 2015 Feb 1;19(1):54-6.

Ismail AA, Abdelkader MF, Mohamed AA, Abdelaziz ST. Evaluation of Efficacy and Lateral Gaze Incomitance in Symmetrical and Asymmetrical Surgery for Concomitant Esotropia and Exotropia. Clinical Ophthalmology. 2021 Aug 26:3613-21.

Israel MD, Basu R, Choudhury M. Efficacy of hang-back recessions in comparison to conventional recessions. Journal of American Association for Pediatric Ophthalmology and Strabismus {JAAPOS}. 2011 Feb 1;15(1):e21-2.

Israel MD, Deka A. Unilateral medial rectus recession for moderate angle esotropia. Journal of American Association for Pediatric Ophthalmology and Strabismus {JAAPOS}. 2007 Feb 1;11(1):92.

Issaho DC, de Freitas D, Cronemberger MF. Plication versus resection in horizontal strabismus surgery: a systematic review with meta-analysis. Journal of ophthalmology. 2020 Jul 3;2020:1-0.

Issaho DC, Wang SX, De Freitas D, Weakley Jr DR. Variability in response to bilateral medial rectus recessions in infantile Esotropia. Journal of Pediatric Ophthalmology & Strabismus. 2016 Sep 1;53(5):305-10.

Jameson PC. The surgical entity of muscle recession. Archives of Ophthalmology. 1931 Sep 1;6(3):329-61.

Jefferis JM, Raoof N, Burke JP. Prioritising downgaze alignment in the management of vertical strabismus for thyroid eye disease: principles and outcomes. Eye. 2020 May;34(5):906-14.

Jeon H, Choi H. Postoperative esotropia: initial overcorrection or consecutive esotropia?. European Journal of Ophthalmology. 2017 Nov 8;27(6):652-7.

Jeong IY, Park YG, Park SW. Long-term Follow-up Results of Partially Accommodative Esotropia After Near Geared Standard Surgery. Journal of the Korean Ophthalmological Society. 2008 Apr 1;49(4):628-33.

Jiang D, Han D, Zhang J, Pei T, Zhao Q. Clinical study of the influence of preoperative wearing time on postoperative effects in children with partially accommodative esotropia. Medicine. 2018 May;97(19).

Jo SH, Lee JE, Choi HY, Jung JH. Clinical Characteristics and Treatment of Esotropia Following Bare Sclera Pterygium Surgery. Journal of the Korean Ophthalmological Society. 2013 May 15;54(5):771-6.

Johnston SC, Crouch Jr ER, Crouch ER. An innovative approach to transposition surgery is effective in treatment of Duane’s syndrome with esotropia. Investigative Ophthalmology & Visual Science. 2006 May 1;47(13):2475-.

Junejo SA, Lodhi AA, Ansari MA. Surgical outcome of 7-millimeter bilateral medial rectus recession in large angle child esotropia. Pak J Med Sci July-September. 2010;26(3):589-94.

Kalita IR, Veena K, Mouttappa F, Sundaralakshmi P, Singh HV. Clinical profile and management of sixth nerve palsy in pediatric patients (0–15 years) in Southern India–A hospital-based study. Indian Journal of Ophthalmology. 2022 Mar;70(3):952.

Kang BW, Park SW, Yoon KC, Heo H. Comparison of Functional Equator-Considering and Parks Methods in Bilateral Medial Rectus Recession for Esotropia. Journal of the Korean Ophthalmological Society. 2012 Jan 1;53(1):138-44.

Kang W, Kim WJ. Surgical outcomes of medial rectus recession and lateral rectus resection for large-angle deviations of acute acquired concomitant esotropia. Korean Journal of Ophthalmology: KJO. 2021 Apr;35(2):101.

Kapoor S, Gupta N, Grover A. A Rare Complication of COVID-19 Vaccination: Cyclical Esotropia. Delhi Journal of Ophthalmology. 2023 Jul 1;33(3):211-3.

Karabas VL, Elibol O. One-stage vs. two-stage adjustable sutures for the correction of esotropia. Strabismus. 2004 Jan 1;12(1):27-34.

Kaufmann H, Sohlenkamp,, Hartwig H. Ergebnisse der operative behandlungbei strabismus convergens. Klin Mbl Augenheilk 1975; 167: 237-244.

Kault DA, Stark DJ, Stark KP. Further investigation of a strabismus model. Australian and New Zealand Journal of Ophthalmology. 1987 Feb;15(1):43-55.

Kaur S, Korla S, Sukhija J. Long-term outcomes of single monocular resection–recession in adult sensory strabismus and factors affecting the postoperative drift. Indian Journal of Ophthalmology. 2023 Jul 1;71(7):2841-4.

Kaur S, Rastogi A, Gupta P, Goyal G, Dash D, Dadeya S. Expansion of binocular fields in the treatment of lateral rectus paresis. CLEVER Clinical and Experimental Vision and Eye Research. 2018 Jul 1;1(2):11.

Keech RV, Scott WE, Baker JD. The medial rectus muscle insertion site in infantile esotropia. American journal of ophthalmology. 1990 Jan 1;109(1):79-84.

Kekunnaya R, Gupta A, Sachdeva V, Krishnaiah S, Venkateshwar Rao B, Vashist U, Ray D. Duane retraction syndrome: series of 441 cases. Journal of Pediatric Ophthalmology & Strabismus. 2012 May 1;49(3):164-9.

Kekunnaya R, Kraft S, Rao VB, Velez FG, Sachdeva V, Hunter DG. Surgical management of strabismus in Duane retraction syndrome. Journal of American Association for Pediatric Ophthalmology and Strabismus. 2015 Feb 1;19(1):63-9.

Kekunnaya R, Negalur M. Duane retraction syndrome: causes, effects and management strategies. Clinical Ophthalmology. 2017 Oct 30:1917-30.

Kekunnaya R, Pineles SL, Thacker N, Velez FG. Surgical outcomes in partially accommodative esotropic Duane syndrome. Journal of American Association for Pediatric Ophthalmology and Strabismus {JAAPOS}. 2011 Feb 1;15(1):e22.

Kekunnaya R, Velez FG, Pineles SL. Outcomes in patients with esotropic duane retraction syndrome and a partially accommodative component. Indian Journal of Ophthalmology. 2013 Dec;61(12):701.

Kennedy A, Lengwiler F, Dosanjh S, Jolly R, Jain S. Effect of bimedial recession on near-distance disparity in esotropia. Eye. 2023 Aug;37(11):2294-8.

Keshtcar-Jafari AR, TaherZadeh M, Anvari F, Ameri A, Ahadzadegan I, AliMahmoodi A, Rajabi MT. Treatment of A and V Pattern Strabismus by Slanting Muscle Insertions in Farabi Eye Hospital. Iranian Journal of Ophthalmology. 2009 Jan 1;21(1):41-6.

Keskinbora KH, Gonen T, Horozoglu F. Outcomes of Surgery in Long-Standing Infantile Esotropia With Cross Fixation. Journal of Pediatric Ophthalmology & Strabismus. 2011 Mar 1;48(3):77-83.

Keskinbora KH, Pulur NK. Long-term results of bilateral medial rectus recession for congenital esotropia. Journal of Pediatric Ophthalmology & Strabismus. 2004 Nov 1;41(6):351-5.

Khalifa YM. Augmented medial rectus recession, medial rectus recession plus Faden, and slanted medial rectus recession for convergence excess esotropia. European journal of ophthalmology. 2011 Mar;21(2):119-24.

Khanna RK, Pasco J, Santallier M, Pisella PJ, Arsene S. Objective ocular torsion outcomes after unilateral horizontal rectus surgery in infantile esotropia. Graefe’s Archive for Clinical and Experimental Ophthalmology. 2018 Sep;256:1783-8.

Kim DH, Noh HJ. Surgical outcomes of acute acquired comitant esotropia of adulthood. BMC ophthalmology. 2021 Dec;21:1-7.

Kim DH, Park SC, Lee YC. Result of Unilateral Medial Rectus Recession in Moderate Angle Esotropia. Journal of the Korean Ophthalmological Society. 1997:1842-6.

Kim DH, Yang HK, Hwang JM. Long-term surgical outcomes of preoperative prism adaptation in patients with partially accommodative esotropia. Eye. 2021 Apr;35(4):1165-70.

Kim E, Choi DG. Comparison of surgical outcomes between bilateral medial rectus recession and unilateral recess-resect for infantile esotropia. Ophthalmic Epidemiology. 2019 Mar 4;26(2):102-8.

Kim E, Choi DG. Outcomes after the surgery for acquired nonaccommodative esotropia. BMC ophthalmology. 2017 Dec;17:1-5.

Kim HK, Chung HJ, Park SH, Shin SY. Consecutive Exotropia After Bilateral Medial Rectus Recession for Infantile Esotropia. Journal of the Korean Ophthalmological Society. 2009 Nov 15;50(11):1712-6.

Kim MJ, Khwarg SI, Kim SJ, Chang BL. Results of re-operation on the deviated eye in patients with sensory heterotropia. Korean Journal of Ophthalmology. 2008 Mar 1;22(1):32-6.

Kim WJ, Kim MM. Subsequent strabismus surgeries in patients with no prior medical records. Indian Journal of Ophthalmology. 2018 Oct;66(10):1451.

King RA, Calhoun JH, Nelson LB. Reoperations for esotropia. Journal of Pediatric Ophthalmology & Strabismus. 1987 May 1;24(3):136-40.

Kinori M, Miller KE, Cochran M, Patil PA, El Sahn M, Khayali S, Robbins SL, Hertle RW, Granet DB. Plication augmentation of the modified Hummelsheim procedure for treatment of large-angle esotropia due to abducens nerve palsy and type 1 Duane syndrome. Journal of American Association for Pediatric Ophthalmology and Strabismus. 2015 Aug 1;19(4):311-5.

Kitzmann AS, Mohney BG, Diehl NN. Short-term motor and sensory outcomes in acquired nonaccommodative esotropia of childhood. Strabismus. 2005 Jan 1;13(3):109-14.

Kiziltunc PB, Atilla H, Çalış F, Alay C. Comparison of surgical success for infantile esotropia and strabismus associated with neurological impairment. Strabismus. 2016 Jul 2;24(3):97-100.

Knapp P. Vertically incomitant horizontal strabismus: the so-called “A” and “V” syndromes. Transactions of the American Ophthalmological Society. 1959;57:666.

Koc F, Durlu N, Ozal H, Yasar H, Firat E. Single-stage adjustable strabismus surgery under topical anesthesia and propofol. Strabismus. 2005 Jan 1;13(4):157-61.

Kozeis N, Triantafylla M, Adamopoulou A, Veliki S, Kozei A, Tyradellis S. A modified surgical technique to treat strabismus in complete sixth nerve palsy. Ophthalmology and Therapy. 2018 Dec;7:369-76.

Kraft SP, Scott WE. Surgery for congenital esotropia-an age comparison study. Journal of Pediatric Ophthalmology & Strabismus. 1984 Mar 1;21(2):57-68.

Kraft SP. A surgical approach for Duane syndrome. Journal of Pediatric Ophthalmology & Strabismus. 1988 May 1;25(3):119-30.

Kraus DJ, Bullock JD. Treatment of thyroid ocular myopathy with adjustable and nonadjustable suture strabismus surgery. Transactions of the American Ophthalmological Society. 1993;91:67.

Kurup SP, Barto HW, Myung G, Mets MB. Stereoacuity outcomes following surgical correction of the nonaccommodative component in partially accommodative esotropia. Journal of american association for pediatric ophthalmology and strabismus. 2018 Apr 1;22(2):92-6.

Kushner BJ, Fisher MR, Lucchese NJ, Morton GV. Factors influencing response to strabismus surgery. Archives of Ophthalmology. 1993 Jan 1;111(1):75-9.

Kushner BJ, Lucchese NJ, Morton GV. Should recessions of the medial recti be graded from the limbus or the insertion?. Archives of Ophthalmology. 1989 Dec 1;107(12):1755-8.

Kushner BJ, Morton GV. The effect of surgical technique and amount, patient age, abduction quality and deviation magnitude on surgical success rates in infantile esotropia. Binocular Vision. 1987;2(1):25-40.

Kushner BJ, Preslan MW, Morton GV. Treatment of partly accommodative esotropia with a high accommodative convergence-accommodation ratio. Archives of Ophthalmology. 1987 Jun 1;105(6):815-8.

Kushner BJ. Evaluation of the posterior fixation plus recession operation with saccadic velocities. Journal of Pediatric Ophthalmology & Strabismus. 1983 Sep 1;20(5):202-9.

Kushner BJ. Fifteen-year outcome of surgery for the near angle in patients with accommodative esotropia and a high accommodative convergence to accommodation ratio. Archives of Ophthalmology. 2001 Aug 1;119(8):1150-3.

Kushner BJ. Insertion slanting strabismus surgical procedures. Archives of Ophthalmology. 2011 Dec 1;129(12):1620-5.

Kushner BJ. Partly accommodative esotropia: should you overcorrect and cut the plus?. Archives of Ophthalmology. 1995 Dec 1;113(12):1530-4.

Kushner BJ. Surgical treatment of teenagers with high AC/A ratios. American Orthoptic Journal. 2014 Jan 1;64(1):37-42.

Kutlutürk I, Eren Z, Koytak A, Sari ES, Alis A, Özertürk Y. Surgically induced astigmatism following medial rectus recession: short-term and long-term outcomes. Journal of Pediatric Ophthalmology & Strabismus. 2014 May 1;51(3):171-6.

Kutschke PJ, Keech RV. Surgical outcome after prism adaptation for esotropia with a distance-near disparity. Journal of American Association for Pediatric Ophthalmology and Strabismus. 2001 Jun 1;5(3):189-92.

Kutschke PJ, Scott WE, Stewart SA. Prism adaptation for esotropia with a distance-near disparity. Journal of Pediatric Ophthalmology & Strabismus. 1992 Jan 1;29(1):12-5.

Kyeong JW, Choi MY. Clinical Course of Patients with Consecutive Esotropia Angle Larger Than Preoperative Angle After Exotropia Surgery. Journal of the Korean Ophthalmological Society. 2008 Oct 15;49(10):1641-8.

Lai HC, Chen SH, Chen YF, Chiang YS, Yang ML. Early predict the outcomes of refractive accommodative esotropia by initial presentations. Chang Gung Med J. 2004 Dec 1;27(12):887-93.

Lam CP, Yam J, Lau FH, Fan DS, Wong CY, Yu CB, Lau WW. SR and LR union suture for the treatment of myopic strabismus fixus: is scleral fixation necessary?. BioMed Research International. 2015 Apr 12;2015.

Latka S. Clinical evaluation and management of’A’or’V’pattern tropias in squint (Doctoral dissertation, Rajiv Gandhi University of Health Sciences (India)). 2009.

Lau FH, Fan DS, Yip WW, Yu CB, Lam DS. Surgical outcome of single-staged three horizontal muscles squint surgery for extra-large angle exotropia. Eye. 2010 Jul;24(7):1171-6.

Lee D, Kim WJ, Kim MM. Surgical outcomes and occurrence of associated vertical strabismus during a 10-year follow-up in patients with infantile esotropia. Indian Journal of Ophthalmology. 2021 Jan;69(1):130.

Lee EK, Hwang JM. Prismatic correction of consecutive esotropia in children after a unilateral recession and resection procedure. Ophthalmology. 2013 Mar 1;120(3):504-11.

Lee HJ, Kim JA, Kim SJ, Yu YS. Relation between preoperative hyperopia and surgical outcome in infantile esotropia. International Journal of Ophthalmology. 2018;11(12):1963.

Lee HS, Park SW, Heo H. Acute acquired comitant esotropia related to excessive Smartphone use. BMC ophthalmology. 2016 Dec;16:1-7.

Lee KW, Paik HJ. Risk Factors for Consecutive Exotropia and Hyperopic Changes after Bilateral Medial Rectus Recession. Journal of the Korean Ophthalmological Society. 2018 Mar 15;59(3):276-81.

Lee TE, Kim SH. Accommodative and tonic convergence and anatomical contracture in partially accommodative and non-accommodative esotropia. Ophthalmic and Physiological Optics. 2012 Nov;32(6):535-8.

Legrand A, Quoc EB, Vacher SW, Ribot J, Lebas N, Milleret C, Bucci MP. Postural control in children with strabismus: effect of eye surgery. Neuroscience letters. 2011 Aug 26;501(2):96-101.

Leitch RJ, Burke JP, Strachan IM. Convergence excess esotropia treated surgically with fadenoperation and medical rectus muscle recessions. British journal of ophthalmology. 1990 May 1;74(5):278-9.

Lekskul A, Chotkajornkiat N, Wuthisiri W, Tangtammaruk P. Acute acquired comitant esotropia: etiology, clinical course, and management. Clinical Ophthalmology. 2021 Apr 15:1567-72.

Lekskul A, Tangtammaruk P, Wuthisiri W. The outcome of one-to-Four muscle surgery by intraoperative relaxed muscle positioning with adjustable suture technique in thyroid eye disease. Clinical Ophthalmology. 2021 Sep 11:3833-9.

Lim CW, Lee J, Kim WJ. Changes in the number and characteristics of patients with acute acquired concomitant esotropia over time: An 8-year retrospective study. Medicine. 2023 Jun 6;102(22).

Lim L, Rosenbaum AL, Demer JL. Saccadic velocity analysis in patients with divergence paralysis. Journal of Pediatric Ophthalmology & Strabismus. 1995 Mar 1;32(2):76-81.

Lim SH, Lee YG, Kim US. Non-adjustable surgery for acute acquired comitant esotropia under general anesthesia. BMC ophthalmology. 2022 Dec;22(1):1-6.

Lingua RW, Feuer W. Intraoperative succinylcholine and the postoperative eye alignment. Journal of Pediatric Ophthalmology & Strabismus. 1992 May 1;29(3):167-70.

Lipton JR, Willshaw HE. Prospective multicentre study of the accuracy of surgery for horizontal strabismus. British journal of ophthalmology. 1995 Jan 1;79(1):10-1.

Liu GT, Hertle RW, Quinn GE, Schaffer DB. Comitant esodeviation resulting from neurologic insult in children. Journal of American Association for Pediatric Ophthalmology and Strabismus. 1997 Sep 1;1(3):143-6.

Liu H, Cao Y, Li R, Wu J. Development and Validation of a Nomogram for Predicting Second Surgery in Patients with Concomitant Esotropia. Ophthalmology and Therapy. 2022 Dec;11(6):2169-82.

Lucas E, Bentley CR, Aclimandos WA. The effect of surgery on the AC/A ratio. Eye. 1994 Jan;8(1):109-14.

LYLE TK. Management of Ocular Palsies of Traumatic Origin Excluding Reconstructive Surgery in Cases of Fracture of the Orbit. International Ophthalmology Clinics. 1971 Dec 1;11(4):146-76.

Lyu IJ, Lee JY, Kong M, Park KA, Oh SY. Surgical responses of medial rectus muscle recession in thyroid eye disease-related esotropia. Plos one. 2016 Jan 21;11(1):e0146779.

Ma DJ, Yang HK, Hwang JM. Surgical outcomes of medial rectus recession in esotropia with cerebral palsy. Ophthalmology. 2013 Apr 1;120(4):663-7.

MacLeod JA, Rhatigan MC, Luff AJ, Morris RJ. Bimedial rectus recession using the anchored hang-back technique. Ophthalmic Surgery, Lasers and Imaging Retina. 1997 Apr 1;28(4):343-6.

Magli A, Carelli R, Matarazzo F, Bruzzese D. Essential infantile esotropia: postoperative motor outcomes and inferential analysis of strabismus surgery. BMC ophthalmology. 2014 Dec;14(1):1-7.

Martin-Doyle C. Cosmetic cure of alternating squint. American journal of ophthalmology. 1950 Aug;33(8):1290-1.

Masoomian B, Akbari MR, Mirmohamadsadeghi A, Aghsaei Fard M, Khorrami-Nejad M, Hamad N, Heirani M. Clinical characteristics and surgical approach in Duane retraction syndrome: a study of 691 patients. Japanese Journal of Ophthalmology. 2022 Sep;66(5):474-80.

Matlach J, Döllinger VK, Eha J, Elflein HM, Weyer-Elberich V, Mildenberger P, Pitz S. Ocular ductions after rectus muscle recession and resection in thyroid eye disease. Strabismus. 2019 Jul 3;27(3):143-8.

Maudgil A, Raoof N, Burke J. The value of fusional convergence amplitudes in esodeviation surgery without adjustable sutures. Journal of Pediatric Ophthalmology & Strabismus. 2018 Nov 19;55(6):375-81.

McKeown CA, Lambert HM, Shore JW. Preservation of the anterior ciliary vessels during extraocular muscle surgery. Ophthalmology. 1989 Apr 1;96(4):498-507.

Merriam SW, Kushner BJ. An investigation into the mechanisms causing antipodean strabismus. Journal of American Association for Pediatric Ophthalmology and Strabismus. 2007 Jun 1;11(3):249-53.

Merrill KS, Areaux Jr RG. Unilateral Horizontal Rectus Muscle Recessions for Pediatric Comitant Strabismus. Journal of Binocular Vision and Ocular Motility. 2022 Jul 3;72(3):147-50.

Mets MB, Beauchamp C, Haldi BA. Binocularity following surgical correction of strabismus in adults. Journal of American Association for Pediatric Ophthalmology and Strabismus. 2004 Oct 1;8(5):435-8.

Metwally H. Comparative study between medial rectus slanting versus medial rectus Faden with or without recession for management of esotropia with distance/near disparity: a retrospective study. Journal of Medicine in Scientific Research. 2022 Apr 1;5(2):194.

Metwally H. Comparative Study of the Faden Technique versus the Y split recession in management of Esotropia with Near-Distance disparity. Minia Journal of Medical Research. 2019 Mar 1;30(1):267-74.

Metwally H. Rate of reoperation after strabismus surgery. Journal of Medicine in Scientific Research. 2022 Jan 1;5(1):59.

Mikhail M, Flanders M. Clinical profiles and surgical outcomes of adult esotropia. Canadian Journal of Ophthalmology. 2017 Aug 1;52(4):403-8.

Miles DR, Burian HM. “Computer statistical analysis of symmetrical and asymmetrical surgery in esoptropia.” Trans Am Acad Ophthalmol & Otolaryngol 71 (1967): 290-300.

Millicent M, Peterseim W, Buckley EG. Medial rectus fadenoperation for esotropia only at near fixation. Journal of American Association for Pediatric Ophthalmology and Strabismus. 1997 Sep 1;1(3):129-33.

Mims 3rd JL, Wood RC. Outcome of a surgical treatment protocol for late consecutive exotropia following bilateral medial rectus recession for esotropia. Binocular Vision & Strabismus Quarterly. 2004 Mar 1;19(4):201-6.

Mims JL, Treff G, Kincaid M, Schaffer B, Wood R. Quantitative surgical guidelines for bimedial recession for infantile esotropia. Binocular Vision. 1985;1:7-22.

Mims JL, Treff G, Wood RC. Variability of strabismus surgery for acquired esotropia. Archives of Ophthalmology. 1986 Dec 1;104(12):1780-2.

Mims JL. Factors influencing response to strabismus surgery. Archives of Ophthalmology. 1993 Oct 1;111(10):1312-.

MIN EJ, LEE MK, PARK BI. A clinical study on strabismus in children. Journal of the Korean Ophthalmological Society. 1991:319-28.

Mirza UT, Siddiqui ZK. The outcome of Hang-Back Sutures Technique in Horizontal Strabismus Surgery. Annals of King Edward Medical University. 2021 Mar 17;27(1):43-7.

Mitchell L, Kowal L. Medial rectus muscle pulley posterior fixation sutures in accommodative and partially accommodative esotropia with convergence excess. Journal of American Association for Pediatric Ophthalmology and Strabismus. 2012 Apr 1;16(2):125-30.

Miyata M, Suda K, Uji A, Hata M, Oishi A, Nakano E, Yamamoto A, Nakao S, Ohtsuki H, Tsujikawa A. One-year outcome predictors of strabismus surgery from anterior segment optical coherence tomography with multiple B-scan averaging. Scientific Reports. 2019 Feb 21;9(1):2523.

Mohan K, Sharma A, Pandav SS. Unilateral lateral rectus muscle recession and medial rectus muscle resection with or without advancement for postoperative consecutive exotropia. Journal of American Association for Pediatric Ophthalmology and Strabismus. 2006 Jun 1;10(3):220-4.

Mohan K, Sharma SK. Long-term Motor and Sensory Outcomes After Unilateral Medial Rectus Recession-Lateral Rectus Resection for Infantile Esotropia. Journal of Pediatric Ophthalmology & Strabismus. 2023 Aug 1:1-8.

Mojon DS. Comparison of a new, minimally invasive strabismus surgery technique with the usual limbal approach for rectus muscle recession and plication. British journal of ophthalmology. 2007 Jan 1;91(1):76-82.

Mokhtar NH, Chew FL. Unilateral four-recti muscle surgery in large angle sensory esotropia. Eye South East Asia Vol.16 Issue 1 2021.

Moon HJ, Park SW, Park YG. Bilateral Medial Rectus Recession Posterior to the Functional Equator in Esotropia Over 40 Prism Diopters. Journal of the Korean Ophthalmological Society. 2009 Mar 15;50(3):429-34.

Morad Y, Pras E, Goldich Y, Barkana Y, Zadok D, Hartstein M. Surgical treatment of esotropia associated with high myopia: unilateral versus bilateral surgery. European Journal of Ophthalmology. 2010 Jul;20(4):653-8.

Morad Y, Pras E, Nemet A. Superior and lateral rectus myopexy for acquired adult distance esotropia: a “one size fits all” surgery. Strabismus. 2017 Jul 3;25(3):140-4.

Morrison DG, Emanuel M, Donahue SP. Surgical management of residual or recurrent esotropia following maximal bilateral medial rectus recession. Archives of Ophthalmology. 2011 Feb 14;129(2):173-5.

Mostafa AM, Kassem RR. A comparative study of medial rectus slanting recession versus recession with downward transposition for correction of V-pattern esotropia. Journal of American Association for Pediatric Ophthalmology and Strabismus. 2010 Apr 1;14(2):127-31.

Motley III WW, Melson AT, Gray ME, Salisbury SR. Outcomes of strabismus surgery for esotropia in children with Down syndrome compared with matched controls. Journal of Pediatric Ophthalmology & Strabismus. 2012 Jul 1;49(4):211-4.

Mourits MP, Koorneef L, van Mourik-Noordenbos AM, Van Der Meulen-Schot HM, Prummel MF, Wiersinga WM, Berghout A. Extraocular muscle surgery for Graves’ ophthalmopathy: does prior treatment influence surgical outcome?. British Journal of Ophthalmology. 1990 Aug 1;74(8):481-3.

Muchnick RS, Sanfilippo S, Dunlap EA. Cyclic esotropia developing after strabismus surgery. Archives of Ophthalmology. 1976 Mar 1;94(3):459-60.

Mullaney P, Vajsar J, Smith R, Buncic JR. The natural history and ophthalmic involvement in childhood myasthenia gravis at the hospital for sick children. Ophthalmology. 2000 Mar 1;107(3):504-10.

Muste JC, Wang K, Hwang CJ, Perry JD, Traboulsi EI. Outcomes of the intraoperative relaxed muscle positioning technique in strabismus surgery for thyroid eye disease. Journal of American Association for Pediatric Ophthalmology and Strabismus. 2023 Dec 1;27(6):340-e1.

Muz OE, Sanac AS. Effects of surgical timing on surgical success and long-term motor and sensory outcomes of infantile esotropia. Journal of Pediatric Ophthalmology & Strabismus. 2020 Sep 1;57(5):319-25.

Na KH, Cho YA, Kim SH. Time and factors affecting the direction of re-drift in essential infantile esotropia. Journal of Pediatric Ophthalmology & Strabismus. 2018 Mar 1;55(2):93-9.

Nabie R, Manouchehri V, Meydan SR. Outcomes of Bilateral Lateral Rectus Resection in Residual Esotropia following Bilateral Medial Rectus Recession. Journal of Current Ophthalmology. 2022 Apr;34(2):247.

Nadeem S, Naeem BA, Khan F. Adjustable strabismus surgery: an early glance. Pak J Ophthalmol 2018;34(2): 89-97.

Nadeem S. Pain Score in Adjustable Strabismus Surgery. Pakistan Journal of Ophthalmology. 2020 Mar 17;36(2).

Nadeem S. Types of Pattern Strabismus and Their Surgical Outcomes after Adjustable Strabismus Surgery. Pakistan Journal of Ophthalmology. 2021;37(1).

Natan K, Traboulsi EI. Unilateral rectus muscle recession in the treatment of Duane syndrome. Journal of American Association for Pediatric Ophthalmology and Strabismus. 2012 Apr 1;16(2):145-9.

Neely DE, Helveston EM, Thuente DD, Plager DA. Relationship of dissociated vertical deviation and the timing of initial surgery for congenital esotropia. Ophthalmology. 2001 Mar 1;108(3):487-90.

Neena R, Remya S, Anantharaman G. Acute acquired comitant esotropia precipitated by excessive near work during the COVID-19-induced home confinement. Indian Journal of Ophthalmology. 2022 Apr;70(4):1359.

Nelson LB, Calhoun JH, Simon JW, Wilson TH, Harley RD. Surgical management of large angle congenital esotropia. British journal of ophthalmology. 1987 May 1;71(5):380-3.

Nishikawa N, Kawaguchi Y, Fushitsu R. Prism adaptation response and surgical outcomes of acquired nonaccommodative comitant esotropia. Strabismus. 2023 Jan 2;31(1):9-16.

Nucci P, Serafino M, Trivedi RH, Saunders RA. One-muscle surgery in small-angle residual esotropia. Journal of American Association for Pediatric Ophthalmology and Strabismus. 2007 Jun 1;11(3):269-72.

O’Brien JC, Melson AT, Bryant JC, Ding K, Farris BK, Siatkowski RM. Surgical outcomes following strabismus surgery for abducens nerve palsy. Journal of American Association for Pediatric Ophthalmology and Strabismus. 2023 May 11.

Ocak OB, Inal A, Aygit ED, Ocak SY, Ozturker C, Demirok A, Gokyigit B. Surgical Dose-Response Relationship in Patients with Down Syndrome with Esotropia: A Comparative Study. Age (years). 2017;7(4.81):7-65.

O’Connor AR, Fawcett SI, Stager DR, Birch EE. Factors influencing sensory outcome following surgical correction of infantile esotropia. American Orthoptic Journal. 2002 Jan 1;52(1):69-74.

Oeverhaus M, Fischer M, Hirche H, Schlüter A, Esser J, Eckstein AK. Tendon Elongation with Bovine Pericardium in Patients with Severe Esotropia after Decompression in Graves’ Orbitopathy—efficacy and Long-term Stability. Strabismus. 2018 Apr 3;26(2):62-70.

Oguz V, Yolar M, Pazarli H. Extraocular muscle surgery in dysthyroid orbitomyopathy: influence of previous conditions on surgical results. Journal of Pediatric Ophthalmology & Strabismus. 2002 Mar 1;39(2):77-80.

Oh SY, Lee JY, Park KA, Oh SY. Long-term changes in refractive error and clinical evaluation in partially accommodative esotropia after surgery. PLoS One. 2016 Dec 9;11(12):e0166695.

Oh SY, Park KA, Oh SY. Comparison of recurrent esotropia and consecutive exotropia with horizontal muscle reoperation in infantile esotropia. Japanese Journal of Ophthalmology. 2018 Nov;62:693-8.

Ohmi G, Hosohata J, Okada AA, Fujikado T, Tanahashi N, Uchida I. Strabismus surgery using the intraoperative adjustable suture method under anesthesia with propofol. Japanese journal of ophthalmology. 1999 Nov 1;43(6):522-5.

Olitsky SE, Kelley CJ, Lee H, Nelson LB. Unilateral rectus resection in the treatment of undercorrected or recurrent strabismus. Journal of Pediatric Ophthalmology & Strabismus. 2001 Nov 1;38(6):349-53.

Oliveira JS, Silva RS, Freitas-Costa P, Falcão-Reis F, Breda J, Magalhães A. Strabismus Surgery for Abnormal Head Position: 7-Year Experience at a Tertiary Care Center. Revista Sociedade Portuguesa de Oftalmologia. 2022 Jun 30;46(2):114-9.

Ortube MC, Velez FG, Park HJ, Demer JL. Strabismus due to brain lesions and central fusion disruption: long-term follow-up in a cohort of 50 patients. Journal of American Association for Pediatric Ophthalmology and Strabismus {JAAPOS}. 2011 Feb 1;15(1):e6.

Owens PL, Folk ER, Chen F. Previous Strabismus Surgery and Eye Position Under Anesthesia. Journal of Pediatric Ophthalmology and Strabismus. 1979 Sep 1;16(5):313.

Özkan SB. Restrictive problems related to strabismus surgery. Taiwan Journal of Ophthalmology. 2016 Sep 1;6(3):102-7.

Pamela P, Cotesta PA, Schofield MJ, Stephens D, Kraft S, Mireskandari K. Immediate postoperative angle is the best predictor of long-term strabismus surgery success in children. Journal of American Association for Pediatric Ophthalmology and Strabismus {JAAPOS}. 2013 Feb 1;17(1):e11.

Pamukçu K, Köse S, Cengiz H, Akkin C. The results of surgery in a series of partially accommodative esotropic patients. Strabismus. 1996 Jan 1;4(2):77-82.

Paraskevopoulos K, Karakosta C, Kokolaki A, Droutsas K, Georgalas I, Papakonstantinou D. Long-term astigmatism changes following horizontal muscle recession: a prospective cohort study. Strabismus. 2022 Apr 3;30(2):90-8.

Paraskevopoulos K, Karakosta C, Liaskou M, Feretzakis G, Papakonstantinou D, Droutsas K, Georgalas I. Evaluation of Macular Thickness Changes Following Large Horizontal Rectus Muscle Recession: A Prospective Cohort Study. Cureus. 2023 Aug 8;15(8).

Park KA, Lee GI, Oh SY. Comparison of surgical dose response between divergence insufficiency esotropia and non-accommodative esotropia without divergence insufficiency. Plos one. 2019 Jul 24;14(7):e0220201.

Park KA, Oh SY. Esotropia with an accommodative component after surgery for infantile esotropia compared to primary accommodative esotropia. Journal of American Association for Pediatric Ophthalmology and Strabismus. 2017 Feb 1;21(1):9-14.

Park KA, Oh SY. Extraocular muscle injury during endoscopic sinus surgery: an ophthalmologic perspective. Eye. 2016 May;30(5):680-7.

Park KA, Oh SY. Long-term surgical outcomes of infantile-onset esotropia in preterm patients compared with full-term patients. British Journal of Ophthalmology. 2015 May 1;99(5):685-90.

Park SB, Chung SA, Lee JB. Surgical Results in Duane Retraction Syndrome. Journal of the Korean Ophthalmological Society. 2009 Jun 15;50(6):893-7.

Park SH, Kim HK, Jung YH, Shin SY. Unilateral lateral rectus advancement with medial rectus recession vs bilateral medial rectus recession for consecutive esotropia. Graefe’s archive for clinical and experimental ophthalmology. 2013 May;251:1399-403.

Parks MM, Wheeler MB. Concomitant esodeviation. In: Tasman W, Jaeger EA, eds. Duane’s Clinical Ophthalmology. Vol. 1. Philadelphia, PA: JB Lippincott; 1986: 10.

Parks MM. Management of acquired esotropia. The British Journal of Ophthalmology. 1974 Mar;58(3):240.

Patel HI, Dawson E, Lee J. Surgery for residual convergence excess esotropia. Strabismus. 2011 Dec 1;19(4):153-6.

Patti M. Two brothers with horizontal gaze palsy with progressive scoliosis. Australian Orthoptic Journal. 2017 Jan;49:13-7.

Pellanda N, Mojon DS. Minimally invasive strabismus surgery technique in horizontal rectus muscle surgery for esotropia. Ophthalmologica. 2010 Feb 1;224(2):67-71.

Penne RB, Flanagan JC, Stefanyszyn MA, Nowinski T. Ocular motility disorders secondary to sinus surgery. Ophthalmic Plastic & Reconstructive Surgery. 1993 Mar 1;9(1):53-61.

Penne RB, Flanagan JC, Stefanyszyn MA, Nowinski T. Ocular motility disorders secondary to sinus surgery. Ophthalmic Plastic & Reconstructive Surgery. 1993 Mar 1;9(1):53-61.

Peragallo JH, Bruce BB, Hutchinson AK, Lenhart PD, Biousse V, Newman NJ, Lambert SR. Functional and motor outcomes of strabismus surgery for chronic isolated adult sixth nerve palsy. Neuro-Ophthalmology. 2014 Dec 1;38(6):320-5.

Perez CI, Zuazo F, Zanolli MT, Guerra JP, Acuña O, Iturriaga H. Esotropia surgery in children with Down syndrome. Journal of American Association for Pediatric Ophthalmology and Strabismus. 2013 Oct 1;17(5):477-9.

Petavy–Blanc AS, Fayol N, Ott AC, Feumi–Jantou C, Jounda G, Bousquet A, Barreau E. Bilateral Medial Rectus Recession Combined With Retroequatorial Myopexy as a Surgical Treatment of Large Angle Esotropia at Distance Fixation. Investigative Ophthalmology & Visual Science. 2006 May 1;47(13):2473-.

Phanphruk W, Hennein L, Hunter DG. Strabismus Surgery in Orthophoric Patients With Symptomatic, Asymmetric Vertical or Horizontal Incomitance. American Journal of Ophthalmology. 2023 May 1;249:29-38.

Pichler U, Schmidbauer E, Hermann P, Wagner H, Bolz M, Mursch-Edlmayr AS. A comparative study of various prism adaptation forms in the surgical management of esophoria. Acta Ophthalmologica. 2022 Jun;100(4):e1010-4.

Pickering JD, Simon JW, Ratliff CD, Melsopp KB, Lininger LL. Alignment success following medial rectus recessions in normal and delayed children. Journal of Pediatric Ophthalmology & Strabismus. 1995 Jul 1;32(4):225-7.

Pineles SL, Rosenbaum AL, Kekunnaya R, Velez FG. Medial rectus recession after vertical rectus transposition in patients with esotropic Duane syndrome. Archives of Ophthalmology. 2011 Sep 12;129(9):1195-8.

Polling JR, Eijkemans MJ, Esser J, Gilles U, Kolling GH, Schulz E, Lorenz B, Roggenkämper P, Herzau V, Zubcov A, ten Tusscher MP. A randomised comparison of bilateral recession versus unilateral recession–resection as surgery for infantile esotropia. British journal of ophthalmology. 2009 Jul 1;93(7):954-7.

Pollock DA, Biglan AW. Surgery for Esotropia in Young Adults. American Orthoptic Journal. 1996 Jan 1;46(1):36-43.

Post LT. Suggestions for the Surgical Management of Strabismus: The Fourth Sanford R. Gifford Memorial Lecture. American Journal of Ophthalmology. 1949 Mar 1;32(3):345-50.

Prangen AD. Some Observations on the Surgical Treatment of the Extra-ocular Muscles. Transactions of the American Ophthalmological Society. 1946;44:251-272.

Prat D, Katowitz WR, Strong A, Katowitz JA. Ocular manifestations and surgical interventions in pediatric patients with Koolen-de-Vries syndrome. Ophthalmic Genetics. 2021 Mar 4;42(2):186-8.

Pratt-Johnson JA, Tillson G. Sensory outcome with nonsurgical management of esotropia with convergence excess (a high accommodative convergence/accommodation ratio). Canadian journal of ophthalmology. Journal canadien d’ophtalmologie. 1984 Aug 1;19(5):220-3.

Preslan MW, Cioffi G, Min YI. Refractive error changes following strabismus surgery. Journal of Pediatric Ophthalmology & Strabismus. 1992 Sep 1;29(5):300-4.

Prieto-Diaz J. Large bilateral medial rectus recession in early esotropia with bilateral limitation of abduction. Journal of Pediatric Ophthalmology & Strabismus. 1980 Mar 1;17(2):101-5.

Pruna VI, Cioplean D, Voinea LM. Therapeutic results in sixth nerve palsy. Romanian Neurosurgery. 2015 Mar 15:51-8.

Puligadda RK. Surgical planning for Duane retraction syndrome. J Clin Res Ophthalmol 2 (2): 019-025. DOI: 10.17352/2455. 2015;1414:019.

Qi D, Gao L, Xie J, Yu T. Clinical Research on the Efficacy of Modified Surgery for Esotropia Fixus With High Myopia. Journal of Pediatric Ophthalmology & Strabismus. 2018 Jul 1;55(4):219-24.

Quaranta-Leoni FM, Quaranta-Leoni C, Di Marino M. Marcus-Gunn jaw winking syndrome: case series study and management algorithm. Orbit. 2023 Mar 2:1-8.

Quéré MA, Péchereau A. Surgical Results in Functional Squints: Analysis and Pathogenetic Meaning. Ophthalmologica. 1981 Mar 30;182(2):104-12.

Quigley C, Cairns M, McElnea E, Doyle F, McCance J, Mullaney P. A retrospective evaluation of bilateral medial rectus recession for management of accommodative esotropia according to prism-adapted motor response preoperatively. Journal of American Association for Pediatric Ophthalmology and Strabismus. 2017 Apr 1;21(2):157-9.

Rabinovich R, Demer JL. Abnormal rectus muscle length in horizontal strabismus. Journal of American Association for Pediatric Ophthalmology and Strabismus {JAAPOS}. 2013 Feb 1;17(1):e8.

Raiyawa T, Jariyakosol S, Praneeprachachon P, Pukrushpan P. Outcomes of 3 or 4 horizontal muscles surgery in large-angle exotropia. The Asia-Pacific Journal of Ophthalmology. 2015 Jul 1;4(4):208-11.

Rajavi Z, Feizi M, Nabavi SA, Sabbaghi H, Behradfar N, Yaseri M, Faghihi M, Abdi S. Slanted versus augmented recession for horizontal strabismus. Journal of Ophthalmic & Vision Research. 2019 Oct;14(4):465.

Rajavi Z, Gozin M, Sabbaghi H, Behradfar N, Kheiri B, Faghihi M. Reoperation in horizontal strabismus and its related risk factors. Medical Hypothesis, Discovery and Innovation in Ophthalmology. 2018;7(2):73.

Rajavi Z, Mohammad Rabei H, Ramezani A, Heidari A, Daneshvar F. Refractive effect of the horizontal rectus muscle recession. International Ophthalmology. 2008 Apr;28:83-8.

Ramadan AE, Abd El GZ, Rajab R, El-Saadani AE, Marey HM. Management of duane retraction syndrome. Journal of the Egyptian Ophthalmological Society. 2019 Oct 1;112(4):130-6.

Ramawat A, Singh A, Agrawal A, Panyala R, Verma R, Mittal SK, Kumar B. Determinants of postoperative alignment in patients of infantile esotropia. Himalayan Journal Of Ophthalmology. 2023 Jan 1;17(1):11-7.

Ravindran M, Pawar N, Karkera RP, Kunnatur R. Clinical analysis, management and outcome of Duane’s retraction syndrome at a Tertiary Eye Care Centre in South India. Delhi Ophthalmol Soc. 2013 Sep 20;24:97-101.

Reader AL. Age-related distance esotropia. Journal of American Association for Pediatric Ophthalmology and Strabismus {JAAPOS}. 2007 Feb 1;11(1):69-70.

Repka MX, Connett JE, Baker JD, Rosenbaum AL. Surgery in the prism adaptation study: accuracy and dose response. Journal of Pediatric Ophthalmology & Strabismus. 1992 May 1;29(3):150-6.

Repka MX, Downing E. Characteristics and surgical results in patients with age-related divergence insufficiency esotropia. Journal of American Association for Pediatric Ophthalmology and Strabismus. 2014 Aug 1;18(4):370-3.

Repka MX, Fishman PJ, Guyton DL. The site of reattachment of the extraocular muscle following hang-back recession. Journal of Pediatric Ophthalmology & Strabismus. 1990 Nov 1;27(6):286-90.

Rhiu S, Michalak S, Phanphruk W, Hunter DG. Anomalous vertical deviations in attempted abduction occur in the majority of patients with esotropic Duane syndrome. American Journal of Ophthalmology. 2018 Nov 1;195:171-5.

Richards AB, Iqbal O, Walsh A. Surgical techniques for sagging eye syndrome: avoiding lateral rectus surgery. Journal of American Association for Pediatric Ophthalmology and Strabismus {JAAPOS}. 2022 Aug 1;26(4):e29.

Ridley-Lane M, Lane E, Yeager LB, Brooks SE. Adult-onset chronic divergence insufficiency esotropia: clinical features and response to surgery. Journal of American Association for Pediatric Ophthalmology and Strabismus. 2016 Apr 1;20(2):117-20.

Robb RM, Rodier DW. The broad clinical spectrum of early infantile esotropia. Transactions of the American Ophthalmological Society. 1986;84:103.

Robb RM, Rodier DW. The variable clinical characteristics and course of early infantile esotropia. Journal of Pediatric Ophthalmology & Strabismus. 1987 Nov 1;24(6):276-81.

Roberts CJ, Murphy MF, Adams GG, Lund VJ. Strabismus following endoscopic orbital decompression for thyroid eye disease. Strabismus. 2003 Jan 1;11(3):163-71.

Roda M, Pellegrini M, Rosti A, Fresina M, Schiavi C. Augmented bimedial rectus muscles recession in acute acquired concomitant esotropia associated with myopia. Canadian Journal of Ophthalmology. 2021 Jun 1;56(3):166-70.

Romano P, Gabriel L. Intraoperative adjustment of eye muscle surgery: correction based on eye position during general anesthesia. Archives of Ophthalmology. 1985 Mar 1;103(3):351-3.

Romano PE, Gabriel L, Bennett WL, Snyder BM. Stage I intraoperative adjustment of eye muscle surgery under general anesthesia: consideration of graduated adjustment. Graefe’s archive for clinical and experimental ophthalmology. 1988 May;226:235-40.

Rosenbaum AL, Egbert JE, Keogan T, Wheeler N, Wang C, Buzard K. Length-tension properties of extraocular muscles in patients with esotropia and intermittent exotropia. American journal of ophthalmology. 1994 Jun 1;117(6):791-9.

Rosenbaum AL, Jampolsky A, Scott AB. Bimedial recession in high AC/A esotropia: a long-term follow-up. Archives of Ophthalmology. 1974 Apr 1;91(4):251-3.

Rosenbaum AL, Santiago AP. Clinical strabismus management principles and surgical techniques. Philadelphia: W. B. Saunders Company; 1999: 552-555.

Rosenbaum AL, Weiss SJ, Bateman JB, Liu PY. Quantitative analysis of spring forces in esotropia and exotropia during surgery. Journal of Pediatric Ophthalmology & Strabismus. 1982 Jan 1;19(1):7-9.

Roshani D. Study of the clinical course of abducens nerve palsy and outcome of its surgical intervention (Doctoral dissertation, Madras Medical College, Chennai).

Roth A. Which Angle for which Surgical Strategy in Comitant Strabismus?. American Orthoptic Journal. 2003 Jan 1;53(1):75-87.

Rowe FJ. Long-term postoperative stability in infantile esotropia. Strabismus. 2000 Jan 1;8(1):3-13.

Roy M, Maurya RP, Singh VP, Singh MK, Loha S, Singh TB. Comparison of incidence of oculocardiac reflex between hang-back and conventional rectus recession in horizontal strabismus surgery. Indian Journal of Clinical and Experimental Ophthalmology. 2020;6(4):518-21.

Ruatta C, Schiavi C. Acute acquired concomitant esotropia associated with myopia: is the condition related to any binocular function failure?. Graefe’s Archive for Clinical and Experimental Ophthalmology. 2020 Nov;258:2509-15.

Ruiz MF, Álvarez MT, Sánchez-Garfido CM, Hernáez JM, Rodríguez JM. Surgery and botulinum toxin in congenital esotropia. Canadian journal of ophthalmology. 2004 Oct 1;39(6):639-49.

Saeed H. Evaluation of adjustable muscle surgery under general anesthesia with rapid recovery profile in children. Adv Ophthalmol Vis Syst. 2015;2(6):212-6.

Saeed M, Mahmood MA, Aslam S, Khan KR, Shaheen A, Qamar AR, Aasi NA. Augmented Recession of Horizontal Recti for the treatment of large angle Horizontal Tropias: A Safe & Effective Approach. AN OFFICIAL JOURNAL OF PESHAWAR MEDICAL COLLEGE. 2011 Oct;9(4):66.

Saleh MA, Abdelhalim NE. Outcome of management o finferior oblique inclusion syndrome associated with recurrent strabismus. International Journal of Current Research. 2018; 10(2): 65939-65942.

Sanjari MS, Shahraki K, Davoudi F, Abriaghdam K, Nekoozadeh S, Shahraki K. The outcome of bilateral medial rectus muscle recession in esotropia. Journal of Current Ophthalmology. 2014;26(1):33.

Saunders RA, Wilson ME, Bluestein EC, Sinatra RB, Kraft SP. Surgery on the Normal Eye in Duane Retraction Syndrome/Surgery on the Normal Eye in Duane Retraction Syndrome: Discussion. Journal of Pediatric Ophthalmology & Strabismus. 1994 May 1;31(3):162-71.

Savino G, Mattei R, Salerni A, Fossataro C, Pafundi PC. Long-term follow-up of surgical treatment of thyroid-associated orbitopathy restrictive strabismus. Frontiers in Endocrinology. 2022 Nov 10;13:1030422.

Schäfer WD, Kellermann A. Results of surgical treatment in strabismus convergens. Albrecht von Graefes Archiv für klinische und experimentelle Ophthalmologie. 1976 Jan;198:207-15.

Schliesser JA, Sprunger DT, Helveston EM. Type 4 Duane syndrome. Journal of American Association for Pediatric Ophthalmology and Strabismus. 2016 Aug 1;20(4):301-4.

Schlossman A, Shier JM. Criteria for the management of alternating strabismus. American Journal of Ophthalmology. 1955 Mar 1;39(3):351-8.

Scobee RG. Degrees of correction per millimeter of surgery. American Journal of Ophthalmology. 1949 Oct 1;32(10):1376-80.

Scobee RG. Intermittent exotropia. Transactions-American Academy of Ophthalmology and Otolaryngology. American Academy of Ophthalmology and Otolaryngology. 1949;53:658-73.

Scott WE, Drummond GT, Keech RV. Vertical offsets of horizontal recti muscles in the management of A and V pattern strabismus. Australian and New Zealand Journal of Ophthalmology. 1989 Aug;17(3):281-8.

Scott WE, Reese PD, Hirsh CR, Flabetich CA. Surgery for large-angle congenital esotropia: two vs three and four horizontal muscles. Archives of Ophthalmology. 1986 Mar 1;104(3):374-7.

Scott WE, Thalacker JA. Diagnosis and treatment of thyroid myopathy. Ophthalmology. 1981 Jun 1;88(6):493-8.

Seher GV, Thaler A, Schneider B, Scheiber V. Combined surgery of convergent squint: A proposal for the amount of recession/resection in unilateral procedures. Spektrum der Augenheilkunde. 1997;4(11):156-62.

Semmlow J, Putteman A, Vercher JL, Gauther G, Berard PV. Surgical modification of the AC/A ratio and the binocular alignment (” Phoria”) at distance; its influence on accommodative esotropia: a study of 21 cases. Binocular vision & strabismus quarterly. 2000 Jan 1;15(2):121-30.

Seo JW, Paik HJ. Decision for Proper Surgical Amount in Consecutive Esotropia Following Bilateral Lateral Rectus Recession. Journal of the Korean Ophthalmological Society. 2018 Jan 1;59(1):67-72.

Sharma M, Ganjoo S, Ganesh S. Occurrence and subsequent development of vertical deviations in patients treated surgically for infantile esotropia. Indian Journal of Ophthalmology. 2023 Jul 1;71(7):2835-40.

Sharma R, Tibrewal S, Majumdar A, Rath S, Ganesh S. Acquired comitant esotropias–comparison of surgical outcomes of accommodative vs non-accommodative types. Strabismus. 2023 Oct 2;31(4):293-305.

Shauly Y, Miller B, Meyer E. Clinical characteristics and long-term postoperative results of infantile esotropia and myopia. Journal of Pediatric Ophthalmology & Strabismus. 1997 Nov 1;34(6):357-64.

Sheth J, Goyal A, Natarajan D, Warkad VU, Sachdeva V, Kekunnaya R. Clinical Profile, Neuroimaging Characteristics, and Surgical Outcomes of Patients With Acute Acquired Non-accommodative Comitant Esotropia. Journal of Pediatric Ophthalmology & Strabismus. 2023 May 1;60(3):218-25.

Shi M, Zhou Y, Qin A, Cheng J, Ren H. Treatment of acute acquired concomitant esotropia. BMC ophthalmology. 2021 Dec;21(1):1-7.

Shin DH, Choi CY, Han SY. Risk factors for spontaneous consecutive exotropia in children with refractive and nonrefractive accommodative esotropia. Japanese Journal of Ophthalmology. 2020 May;64:292-7.

Shin H, Ha SG, Suh YW, Kim SH. Long-term Clinical Outcomes of Patients with Infantile and Refractive Accommodative Esotropia. Journal of the Korean Ophthalmological Society. 2023 Sep 15;64(9):838-44.

Shoshi M, Shoshi A, Bakalli A. Alternating esotropia and surgical correction in both eyes. Medical Archives. 2009 Jul 1;63(4):220.

Singh A, Parihar JK, Mishra SK, Maggon R, Badhani A. Outcome of early surgery in infantile esotropia: Our experience in tertiary care hospital. Medical Journal Armed Forces India. 2017 Apr 1;73(2):129-33.

Singh DH, Singh GU, Aggarwal LP, Chandra PR. “ A” and” V” phenomena. The British Journal of Ophthalmology. 1966 Dec;50(12):718.

Sivakumar K. Analysis of surgical management of AV pattern deviations (Doctoral dissertation, Madras Medical College, Chennai).

Somer D, Cinar FG, Oral B, Ornek F. Combined recession and resection surgery in the management of convergence excess esotropia with different levels of AC/A ratio. Journal of American Association for Pediatric Ophthalmology and Strabismus. 2017 Feb 1;21(1):7-e1.

Song D, Qian J, Chen Z. Efficacy of botulinum toxin injection versus bilateral medial rectus recession for comitant esotropia: a meta-analysis. Graefe’s Archive for Clinical and Experimental Ophthalmology. 2023 May;261(5):1247-56.

Speeg-Schatz C, Roth A. Surgical management in infantile esotropia. Expert Review of Ophthalmology. 2008 Apr 1;3(2):155-64.

Spierer O, Spierer A. Comparison of hang-back and conventional bimedial rectus recession in infantile esotropia. Graefe’s Archive for Clinical and Experimental Ophthalmology. 2010 Jun;248:901-5.

Sridharan NS. To report results of unilateral medial rectus recession in improvement of ocular alignment in patients with unilateral or bilateral esotropic duane syndrome. University Journal of Surgery and Surgical Specialities. 2021 Dec 5;7(4).

Stager DR, Black T, Felius J. Unilateral lateral rectus resection for horizontal diplopia in adults with divergence insufficiency. Graefe’s Archive for Clinical and Experimental Ophthalmology. 2013 Jun;251:1641-4.

Stager DR, Weakley DR, Everett M, Birch EE. Delayed consecutive exotropia following 7-millimeter bilateral medial rectus recession for congenital esotropia. Journal of Pediatric Ophthalmology & Strabismus. 1994 May 1;31(3):147-50.

Stallworth JY, Rasool N, Indaram M. Recurrent divergence-insufficiency esotropia in Machado-Joseph disease (spinocerebellar ataxia type 3). American Journal of Ophthalmology Case Reports. 2022 Dec 1;28:101754.

Stine GT. The surgical treatment of esophoria. American Journal of Ophthalmology. 1951 Sep 1;34(9):1307-13.

Sugar HS. Intermittent exotropia; treatment. The American orthoptic journal. 1952 Sep;2:18-21.

Sugar HS. Management of eye movement restriction (particularly vertical) in dysthyroid myopathy. Annals of Ophthalmology. 1979 Sep 1;11(9):1305-8.

Suk Yoon J, Samuel Kim U. Surgical treatment of cyclic esotropia. Korean J Ophthalmol; 2019;33(6): 571-572.

Sultan E, Kasem MA, Mostafa A, Mohsen TA, El-Zeini R. Surgical Management of Convergence Excess Esotropia. Egyptian Journal of Ophthalmology,(Mansoura Ophthalmic Center). 2023 Sep 1;3(3):137-42.

Suwanrat C, Tengtrisorn S, Singha P. Results of an Initial Strabismus Surgery for Infantile Eso-tropia before and after 2 tropia before and after 2 Years of ears of ears of Age. Available from rcopt.org

Swaminathan M, Shah SV, Mittal S, Gunasekaran A. Results of bilateral medial rectus recession for comitant esotropia in patients with developmental delay. Strabismus. 2014 Sep 1;22(3):138-42.

Tae T, Devould C, Skilbeck L, Hoehn ME, Kerr N. Persistent Binocular Diplopia in Childhood Following Successful Treatment for Esotropia: A Case Series. American Orthoptic Journal. 2014 Jan 1;64(1):117-22.

Talebnejad MR, Johari MK, Khalili MR, Zare M. Supramaximal recession and resection surgery in large-angle strabismus: Outcomes of large interventional case series exotropia and esotropia. Journal of Current Ophthalmology. 2020 Jan 1;32(1):82-7.

Tawfik AM, Ahmed MM, El Sayed SM. Single eye surgery by hang-back recession vs muscle transplantation in large-angle esotropia. The Scientific Journal of Al-Azhar Medical Faculty, Girls. 2020 Jul 1;4(3):507-12.

Taylor HR, Billson FA. Bimedial Rectus Recession for Alternating Esotropia. Australian Journal of Opthalmology. 1977 Feb;5(1):21-3.

Tejedor J, Rodríguez JM. Early retreatment of infantile esotropia: comparison of reoperation and botulinum toxin. British Journal of Ophthalmology. 1999 Jul 1;83(7):783-7.

Thomas SM, Cruz OA. Comparison of two different surgical techniques for the treatment of strabismus in dysthyroid ophthalmopathy. Journal of American Association for Pediatric Ophthalmology and Strabismus. 2007 Jun 1;11(3):258-61.

Thouvenin D, Norbert O. Intraoperative assessment of medial rectus pulley location in strabismus. European Journal of Ophthalmology. 2013 Jan;23(1):13-8.

Thuma TB, Nelson LB. Indications and Outcomes for Extraocular Muscle Disinsertion in Strabismus Surgery. Journal of Pediatric Ophthalmology & Strabismus. 2023 Jul 1;60(4):253-6.

Tibrewal S, Sachdeva V, Ali MH, Kekunnaya R. Comparison of augmented superior rectus transposition with medial rectus recession for surgical management of esotropic Duane retraction syndrome. Journal of American Association for Pediatric Ophthalmology and Strabismus. 2015 Jun 1;19(3):199-205.

Tolkovsky A, Bachar Zipori A, Maharshak I, Aviel Gadot E, Spierer O. Surgical success of lateral rectus resection for residual and recurrent esotropia. International Ophthalmology. 2021 Oct;41(10):3497-503.

Tolun H, Dikici K, Ozkiris A. Long-term results of bimedial rectus recessions in infantile esotropia. Journal of Pediatric Ophthalmology & Strabismus. 1999 Jul 1;36(4):201-5.

Topcu Yilmaz P, Ural Fatihoglu Ö, Sener EC. Acquired comitant esotropia in children and young adults: clinical characteristics, surgical outcomes, and association with presumed intensive near work with digital displays. Journal of Pediatric Ophthalmology & Strabismus. 2020 Jul 1;57(4):251-6.

Torrefranca Jr AB, Santiago AP. Large bilateral medial rectus recession versus three-to-four horizontal muscle surgery for large-angle esodeviations. European Journal of Ophthalmology. 2022 Nov;32(6):3250-7.

Torres CH, Goldchmit M. Bilateral medial rectus recession for Möbius sequence esotropia. Journal of American Association for Pediatric Ophthalmology and Strabismus {JAAPOS}. 2018 Aug 1;22(4):e46.

TOUR R, Asbury T. Overcorrection of esotropia following bilateral 5mm medial rectus recession. InTransactions of the Pacific Coast Oto-Ophthalmological Society annual meeting 1957 (Vol. 38, pp. 145-166).

Tour RL, Asbury T. Overcorrection of Esotropia: Following Bilateral Five-Mm. Medial Rectus Recession. American Journal of Ophthalmology. 1958 May 1;45(5):644-53.

Tour RL. Surgical overcorrection in convergent strabismus. American Orthoptic Journal. 1958 Jan 1;8(1):59-65.

Trigler L, Siatkowski RM. Factors associated with horizontal reoperation in infantile esotropia. Journal of American Association for Pediatric Ophthalmology and Strabismus. 2002 Feb 1;6(1):15-20.

Tsai CB. Adjustable suture strabismus surgery in pediatric patients using pull-string technique. Taiwan Journal of Ophthalmology. 2017 Jan;7(1):38.

Üretmen Ö (Uretmen O), Pamukçu K, KÖse S, Uçak E. Binocular visual function in congenital esotropia after bilateral medial rectus recession with loop suture. Strabismus. 2002 Jan 1;10(3):215-24.

Urist MJ. Atypical accommodative esotropia. American Journal of Ophthalmology. 1956 Jun 1;41(6):955-64.

URIST MJ. The effect of asymmetrical horizontal muscle surgery. AMA Archives of Ophthalmology. 1958 Feb 1;59(2):247-59.

Uzun A, Sahin AK. Factors influencing surgical success in concomitant horizontal strabismus. Acta Medica. 2021 Nov 29;52(4):341-7.

Van Rijn LJ, Langenhorst AE, Krijnen JS, Bakels AJ, Jansen SM. Predictability of strabismus surgery in children with developmental disorders and/or psychomotor retardation. Strabismus. 2009 Sep 1;17(3):117-27.

Velez FG, Pineles SL. The management of vertical deviations in sagging eye syndrome. Acta estrabológica: publicación oficial de la Sociedad Española de Estrabología, Pleóptica, Ortóptica, Visión Binocular, Reeducación y Rehabilitación Visual. 2023 Jan;52(1):20-4.

Velez FG, Rosenbaum AL. Preoperative prism adaptation for acquired esotropia: long-term results. Journal of American Association for Pediatric Ophthalmology and Strabismus. 2002 Jun 1;6(3):168-73.

Villaseca A. Surgical classification of squints with a vertical deviation. The British Journal of Ophthalmology. 1955 Mar;39(3):129.

von Noorden GK, Avilla CW. Nonaccommodative convergence excess. American journal of ophthalmology. 1986 Jan 1;101(1):70-3.

von Noorden GK, Munoz M, Raab EL. Recurrent Esotropia/Discussion. Journal of Pediatric Ophthalmology & Strabismus. 1988 Nov 1;25(6):275-80.

von Noorden GK, Tredici TD, Ruttum M. Pseudo-internuclear ophthalmoplegia after surgical paresis of the medial rectus muscle. American journal of ophthalmology. 1984 Nov 1;98(5):602-8.

Von Noorden GK. Surgical treatment of congenital esotropia. Trans Am Acad Opth Optolaryn-gol. 1972;76:1465.

Wabulembo G, Demer JL. Long-term outcome of medial rectus recession and pulley posterior fixation in esotropia with high AC/A ratio. Strabismus. 2012 Sep 1;20(3):115-20.

Wan MJ, Chiu H, Shah AS, Hunter DG. Long-term surgical outcomes for large-angle infantile esotropia. American Journal of Ophthalmology. 2018 May 1;189:155-9.

Wan MJ, Mantagos IS, Shah AS, Kazlas M, Hunter DG. Comparison of botulinum toxin with surgery for the treatment of acute-onset comitant esotropia in children. American Journal of Ophthalmology. 2017 Apr 1;176:33-9.

Wang Jia-lu., Dong Ling-yan, Cen Jie, & Kang Xiao-li. (2021). The Surgical Effects of Medial Rectus Recession on Divergence Insufficiency Esotropia. JOURNAL OF SICHUAN UNIVERSITY (MEDICAL SCIENCES), 52(6), 1011-1015.

Wang L, Nelson LB. Outcome study of graded unilateral medial rectus recession for small to moderate angle esotropia. Journal of Pediatric Ophthalmology & Strabismus. 2011;48(1):20-4.

Wang L, Wang X. Comparison between graded unilateral and bilateral medial rectus recession for esotropia. British Journal of Ophthalmology. 2011 Jan 1.

Wang L, Wang X. Comparison between graded unilateral and bilateral medial rectus recession for esotropia. British journal of ophthalmology. 2012 Apr 1;96(4):540-3.

Wang T, Wang LH. Surgical treatment for residual or recurrent strabismus. International Journal of Ophthalmology. 2014;7(6):1056.

Wang Z, Fu L, Shen T, Qiu X, Yu X, Shen H, Yan J. Supramaximal Horizontal Rectus Recession–Resection Surgery for Complete Unilateral Abducens Nerve Palsy. Frontiers in Medicine. 2022 Feb 22;8:795665.

Weakley DR, Holland DR. Effect of ongoing treatment of amblyopia on surgical outcome in esotropia. Journal of Pediatric Ophthalmology & Strabismus. 1997 Sep 1;34(5):275-8.

Weeks CL, Hamed LM. Treatment of acute comitant esotropia in Chiari I malformation. Ophthalmology. 1999 Dec 1;106(12):2368-71.

Weisberg OL, Sprunger DT, Plager DA, Neely DE, Sondhi N. Strabismus in pediatric pseudophakia. Ophthalmology. 2005 Sep 1;112(9):1625-8.

Wiggins Jr RE, Baumgartner S. Diagnosis and management of divergence weakness in adults. Ophthalmology. 1999 Jul 1;106(7):1353-6.

Willshaw HE, Mashhoudi N, Powell S. Augmented medial rectus recession in the management of esotropia. British journal of ophthalmology. 1986 Nov 1;70(11):840-3.

Wilson WA. Overcorrections In Strabismus Surgery. Journal of Pediatric Ophthalmology & Strabismus. 1965 Apr 1;2(2):23-6.

Wipf M, Priglinger S, Palmowski-Wolfe A. Y-split recession of the medial rectus muscle as a secondary and/or unilateral procedure in the treatment of esotropia with distance/near disparity. Journal of Ophthalmology. 2017 Jul 19;2017.

Wortham EV, Greenwald MJ. Expanded binocular peripheral visual fields following surgery for esotropia. Journal of Pediatric Ophthalmology & Strabismus. 1989 May 1;26(3):109-12.

Wright KW, Arow M, Zein M, Strube YN. Wright hang-back recession with fibrin glue compared with standard fixed suture recession for the treatment of horizontal strabismus. Canadian Journal of Ophthalmology. 2021 Aug 1;56(4):244-9.

Wright KW, Bruce-Lyle L. Augmented surgery for esotropia associated with high hypermetropia. Journal of Pediatric Ophthalmology & Strabismus. 1993 May 1;30(3):167-70.

Wright KW, Corradetti G. Lateral Rectus Central Plication Versus Medial Rectus Recession in Age-Related Distance Esotropia. Investigative Ophthalmology & Visual Science. 2016 Sep 26;57(12):2446-.

Wright KW, Corradetti G. Wright central plication of lateral rectus versus standard medial rectus recession in adult divergence insufficiency esotropia. Journal of American Association for Pediatric Ophthalmology and Strabismus. 2017 Apr 1;21(2):94-6.

Wygnanski-Jaffe T, Habot-Wilner Z, Glovinsky J, Spierer A. Bilateral Medial Rectus Muscle Recession Results in Children with Developmental Delay Compared to Normally Developed Children. Journal of American Association for Pediatric Ophthalmology and Strabismus {JAAPOS}. 2006 Feb 1;10(1):95.

Wygnanski-Jaffe T, Trotter J, Watts P, Kraft SP, Abdolell M. Preoperative prism adaptation in acquired esotropia with convergence excess. Journal of American Association for Pediatric Ophthalmology and Strabismus. 2003 Feb 1;7(1):28-33.

Xia Q, Wang Z, Yan J. Surgical management of strabismus in patients with orbital fracture. Journal of Craniofacial Surgery. 2018 Oct 1;29(7):1865-9.

Xia Y, Cao L, Peng X, Wang L. Young patients with divergence insufficiency related to excessive near work. Strabismus. 2020 Jul 2;28(3):136-41.

Yabas Kiziloglu O, Ziylan S, Simsek I. Long term motor and sensory outcome after surgery for infantile esotropia and risk factors for residual and consecutive deviations. InSeminars in Ophthalmology 2020 Jan 2 (Vol. 35, No. 1, pp. 27-32). Taylor & Francis.

Yadav I, Singh VP, Bhushan P, Maurya RP, Singh MK. Comparison of surgical success rates and amount of corneal astigmatism induced by hag-back and conventional muscle recession surgery in horizontal strabismus. Indian J Clin Exp Ophthalmol. 2015 Jan;1(1):7-13.

Yagasaki T, Yokoyama Y, Yagasaki A, Eboshita R, Tagami K, Haga Y, Touya A. Surgical Outcomes with and without Prism Adaptation of Cases with Acute Acquired Comitant Esotropia Related to Prolonged Digital Device Use. Clinical Ophthalmology. 2023 Dec 31:807-16.

Yahalom C, Mechoulam H, Cohen E, Anteby I. Strabismus surgery outcome among children and young adults with Down syndrome. Journal of American Association for Pediatric Ophthalmology and Strabismus. 2010 Apr 1;14(2):117-9.

Yahalom C, MScOptom NS. Chiari 1 malformation presenting as strabismus: a case series of 12 patients and review of the literature.

Yang S, MacKinnon S, Dagi LR, Hunter DG. Superior rectus transposition vs medial rectus recession for treatment of esotropic Duane syndrome. JAMA ophthalmology. 2014 Jun 1;132(6):669-75.

Yang Y, Theodorou M, Roberts C, Acheson J, Adams G. Audit of inferior oblique weakening surgery to determine impact on horizontal alignment. Journal of American Association for Pediatric Ophthalmology and Strabismus {JAAPOS}. 2018 Aug 1;22(4):e46.

Yao J, Xia W, Wang X, Zhu W, Jiang C, Ling L, Wu L, Zhao C. Three-muscle surgery for large-angle esotropia in chronic sixth nerve palsy: comparison of two approaches. British Journal of Ophthalmology. 2023 Sep 1;107(9):1377-82.

Yazdani A, Traboulsi EI. Classification and surgical management of patients with familial and sporadic forms of congenital fibrosis of the extraocular muscles. Ophthalmology. 2004 May 1;111(5):1035-42.

Yel G, Acar Z, Vural ET, Mumcu UY, Unculu RB, Nohutçu A. Comparison of the Results of Plication and Resection Techniques in Horizontal Strabismus Surgery. HAYDARPAŞA NUMUNE MEDICAL JOURNAL. 2023 Jan 1;63(1):21-6.

Yu X, Pan W, Tang X, Zhang Y, Lou L, Zheng S, Yao K, Sun Z. Efficacy of augmented-dosed surgery versus botulinum toxin A injection for acute acquired concomitant esotropia: a 2-year follow-up. British Journal of Ophthalmology. 2023 Aug 24.

Yurdakul NS, Bodur S, Koç F. Surgical results of symmetric and asymmetric surgeries and dose-response in patients with infantile esotropia. Turkish Journal of Ophthalmology. 2015 Oct;45(5):197.

Yurdakul NS, Ugurlu S, Maden A. Surgical management of chronic complete sixth nerve palsy. Ophthalmic Surgery, Lasers and Imaging Retina. 2011 Jan 1;42(1):72-7.

Yurdakul NS, Ugurlu S. Analysis of risk factors for consecutive exotropia and review of the literature. Journal of Pediatric Ophthalmology & Strabismus. 2013 Sep 1;50(5):268-73.

Zehavi-Dorin T, Ben-Zion I, Mezer E, Wygnanski-Jaffe T. Long-term results of bilateral medial rectus muscle recession in children with developmental delay. Strabismus. 2016 Jan 2;24(1):7-11.

Zhang C, Belous D, Reynolds AL. Surgical dosing of partial sixth nerve palsy with novel incomitance protocol for improvement of area of single binocular vision. Journal of American Association for Pediatric Ophthalmology and Strabismus {JAAPOS}. 2022 Aug 1;26(4):e36.

Zhang P, Zhang Y, Gao L, Yang J. Comparison of the therapeutic effects of surgery following prism adaptation test versus surgery alone in acute acquired comitant esotropia. BMC ophthalmology. 2020 Dec;20(1):1-6.

Zhou W, Wu H, Zheng Y, Zhang L. The changes of refractive status and anterior segment parameters after strabismus surgery.

Zhou Y, Ling L, Wang X, Jiang C, Wen W, Zhao C. Augmented-Dose Unilateral Recession-Resection Procedure in Acute Acquired Comitant Esotropia. Ophthalmology. 2023 May 1;130(5):525-32.

Zloto O, Simon GB, Fabian ID, Sagiv O, Huna-Baron R, Zion IB, Wygnanski-Jaffe T. Association of orbital decompression and the characteristics of subsequent strabismus surgery in thyroid eye disease. Canadian Journal of Ophthalmology. 2017 Jun 1;52(3):264-8.

## Notes

The authors have no conflicts of interest to disclose.

### Competing Interest Statement

The authors have declared no competing interest.

### Funding Statement

No funding.

### Author Declarations

Office of Research Subjects Protection of the Virginia Commonwealth University approved the study.

### Summary of Updates

Extra paragraph added to results section to clarify that limited data at the smallest deviations.

